# The genetic architecture of fibromyalgia across 2.5 million individuals

**DOI:** 10.1101/2025.09.18.25335914

**Authors:** Isabel Kerrebijn, Gyda Bjornsdottir, Keon Arbabi, Lea Urpa, Hele Haapaniemi, Gudmar Thorleifsson, Lilja Stefansdottir, Stephan Frangakis, Jesse Valliere, Lovemore Kunorozva, Erik Abner, Caleb Ji, Bitten Aagaard, Henning Bliddal, Søren Brunak, Mie T Bruun, Maria Didriksen, Christian Erikstrup, Arni J Geirsson, Daniel F Gudbjartsson, Thomas F Hansen, Ingileif Jonsdottir, Stacey Knight, Kirk U Knowlton, Christina Mikkelsen, Lincoln D Nadauld, Thorunn A Olafsdottir, Sisse R Ostrowski, Ole BV Pedersen, Saedis Saevarsdottir, Astros T Skuladottir, Erik Sørensen, Hreinn Stefansson, Patrick Sulem, Olafur A Sveinsson, Gudny E Thorlacius, Unnur Thorsteinsdottir, Henrik Ullum, Arnor Vikingsson, Thomas M Werge, Chronic Pain Genomics Consortium, FinnGen, DBDS Genomic Consortium, Estonian Biobank Research Team, Genes & Health Research Team, Richa Saxena, Kari Stefansson, Chad M Brummett, Bente Glintborg, Daniel J Clauw, Thorgeir E Thorgeirsson, Frances MK Williams, Nasa Sinnott-Armstrong, Hanna M Ollila, Michael Wainberg

## Abstract

Fibromyalgia is a common and debilitating chronic pain syndrome of poorly understood etiology. Here, we conduct a multi-ancestry genome-wide association study meta-analysis across 2,563,755 individuals (54,629 cases and 2,509,126 controls) from 11 cohorts, identifying the first 26 risk loci for fibromyalgia. The strongest association was with a coding variant in *HTT*, the causal gene for Huntington’s disease. Gene prioritization implicated the *HTT* regulator *GPR52*, as well as diverse genes with neural roles, including *CAMKV*, *DCC*, *DRD2*/*NCAM1*, *MDGA2*, and *CELF4*. Fibromyalgia heritability was exclusively enriched within brain tissues and neural cell types. Fibromyalgia showed strong, positive genetic correlation with a wide range of chronic pain, psychiatric, and somatic disorders, including genetic correlations above 0.7 with low back pain, post-traumatic stress disorder and irritable bowel syndrome. Despite large sex differences in fibromyalgia prevalence, the genetic architecture of fibromyalgia was nearly identical between males and females. This work provides the first robust genetic evidence defining fibromyalgia as a central nervous system disorder, thereby establishing a biological framework for its complex pathophysiology and extensive clinical comorbidities.

## Introduction

Fibromyalgia is a multifaceted syndrome encompassing chronic widespread musculoskeletal pain, fatigue, unrefreshing sleep, cognitive impairment, and somatic symptoms. Prevalence is estimated at ∼2-4% of the adult population, with substantial underdiagnosis^1–3^. It is ∼2-3 times more prevalent in women than in men^3,4^. It is notable for its substantial disease burden and associated health care system utilization^5,6^, with fibromyalgia patients averaging twice as many medical visits as the general population^3,5,6^.

Fibromyalgia is frequently comorbid with variety of disorders including pain conditions, irritable bowel syndrome, myalgic encephalomyelitis/chronic fatigue syndrome (ME/CFS), autoimmune disorders such as rheumatoid arthritis and systemic lupus erythematosus, neuropsychiatric traits such as depression and anxiety, and metabolic syndrome^7,8^. Whether fibromyalgia itself is an autoimmune disease is a matter of longstanding debate among clinical specialists^9^.

Fibromyalgia is considered the prototypical central sensitization or nociplastic pain syndrome^10^. Central sensitization is a neuroplastic process by which the central nervous system becomes excessively sensitive to nociceptive input from the peripheral nervous system, leading to multisite hyperalgesia (an increased pain response to painful stimuli) and allodynia (a pain response to typically non-painful stimuli)^11–13^. Central sensitization is a transdiagnostic contributor to chronic pain, and also occurs in conditions like rheumatoid arthritis and osteoarthritis that involve damage to peripheral tissues^14,15^, although peripheral injury is not necessary to trigger it^12^. Central sensitization has been shown to reflect specific underlying biological processes, including neuroinflammation and glial activation (particularly in the dorsal horn of the spinal cord)^12,16–18^, altered functional connectivity according to magnetic resonance imaging^19,20^, altered neurotransmitter levels^21,22^, and altered neuronal electrophysiology^12^. Other biological processes that may play a role in at least a subset of fibromyalgia patients include (demyelinating) polyneuropathy^23–25^, endocrine dysregulation^26,27^, oxidative stress^28,29^, and dysfunctional mitochondrial energy metabolism^29,30^.

The diagnostic criteria for fibromyalgia have changed over time. The most widely used criteria, from the American College of Rheumatology (ACR)^31^, require widespread pain and tenderness across multiple body regions, alongside some combination of fatigue, difficulty with thinking or remembering, and waking up tired (unrefreshed), all present at a similar level of severity for at least three months. Crucially, this definition removed the prior requirement for patients not to have another explanatory disorder, stating instead that “a diagnosis of fibromyalgia is valid irrespective of other diagnoses”. To date, there are no widely accepted biomarkers for fibromyalgia^3,32^. Treatment often focuses on non-pharmacological treatments like exercise and behavioral interventions, since established pharmacotherapies such as tricyclic antidepressants, cyclobenzaprine, serotonin-norepinephrine reuptake inhibitors, and gabapentinoids are usually only modestly effective (though emerging therapies like low-dose naltrexone, cannabinoids, ketamine and neurostimulation show promise)^4,33^. There is a significant unmet need for effective fibromyalgia therapeutics^34^.

Like most complex traits, fibromyalgia has a partly genetic basis. An early family-based study found that first-degree relatives (parents, full siblings and children) of fibromyalgia patients were 8.5 times more likely to have fibromyalgia themselves, relative to first-degree relatives of rheumatoid arthritis patients^35^. However, attempts to find specific genetic loci associated with fibromyalgia have proven elusive: the GWAS Catalog lists no prior associations with fibromyalgia. A linkage study of 116 families^36^, a genome-wide association study (GWAS) and copy number variant analysis of 313 female fibromyalgia cases and 220 controls^37^, and a GWAS of a quantitative fibromyalgia score across 26,749 individuals^38^ all failed to find loci reaching genome-wide significance. Studies examining pain-related conditions in much larger samples have had greater success. A GWAS of multisite chronic pain – an ordinal scale ranging from 0 to 7 representing the number of body sites (head, face, neck/shoulder, back, stomach/abdomen, hip, knee) at which chronic pain of at least 3 months’ duration was recorded – in 380,000 participants from the UK Biobank^39^ found 76 independent genome-wide significant variants at 39 risk loci^40^. However, this GWAS deliberately excluded participants with chronic pain ‘all over the body’, i.e. chronic widespread pain. Meanwhile, a GWAS of 1308 female cases and 5791 controls specifically focused on chronic widespread pain found one genome-wide significant locus (near *CCT5*/*FAM173B*)^41^, and a more recent chronic widespread pain GWAS of 6914 cases and 242,929 controls from the UK Biobank found three (near *RNF123*, *ATP2C1*, and *COMT*)^42^. Recent GWAS with less specific phenotype definitions have found further risk loci: a joint analysis of 17 pain-related traits in the UK Biobank identified 99 risk loci^43^, while a GWAS of pain intensity in 598,339 Million Veteran Program participants identified 125 loci^44^. However, it is important to note that the chronic widespread pain definitions used in these studies do not fully capture fibromyalgia’s myriad non-pain symptoms.

This study aimed to identify fibromyalgia genetic risk factors through a combined genetic analysis of over 2.5 million individuals, using diagnosis of fibromyalgia by ICD-10 code M79.7. We explored the genetic architecture by identifying novel risk loci, prioritizing causal genes to illuminate underlying biology, defining tissues and cell types with enriched heritability, quantifying genetic overlap with comorbid disorders, and assessing genetic consistency between the sexes.

## Results

### Associations with 26 variants establish genetic underpinnings of fibromyalgia

We performed a multi-ancestry GWAS meta-analysis of fibromyalgia (ICD-10 code M79.7 in inpatient and/or primary care records) across 11 cohorts (Figure 1). Briefly, after performing quality control and GWAS for each cohort-ancestry pair, we harmonized alleles and genome builds across cohorts, corrected for inflation by adjusting p-values by the linkage disequilibrium (LD) score regression intercept, then performed a fixed-effects inverse variance-weighted meta-analysis across cohorts and ancestries.

**Figure 1:**
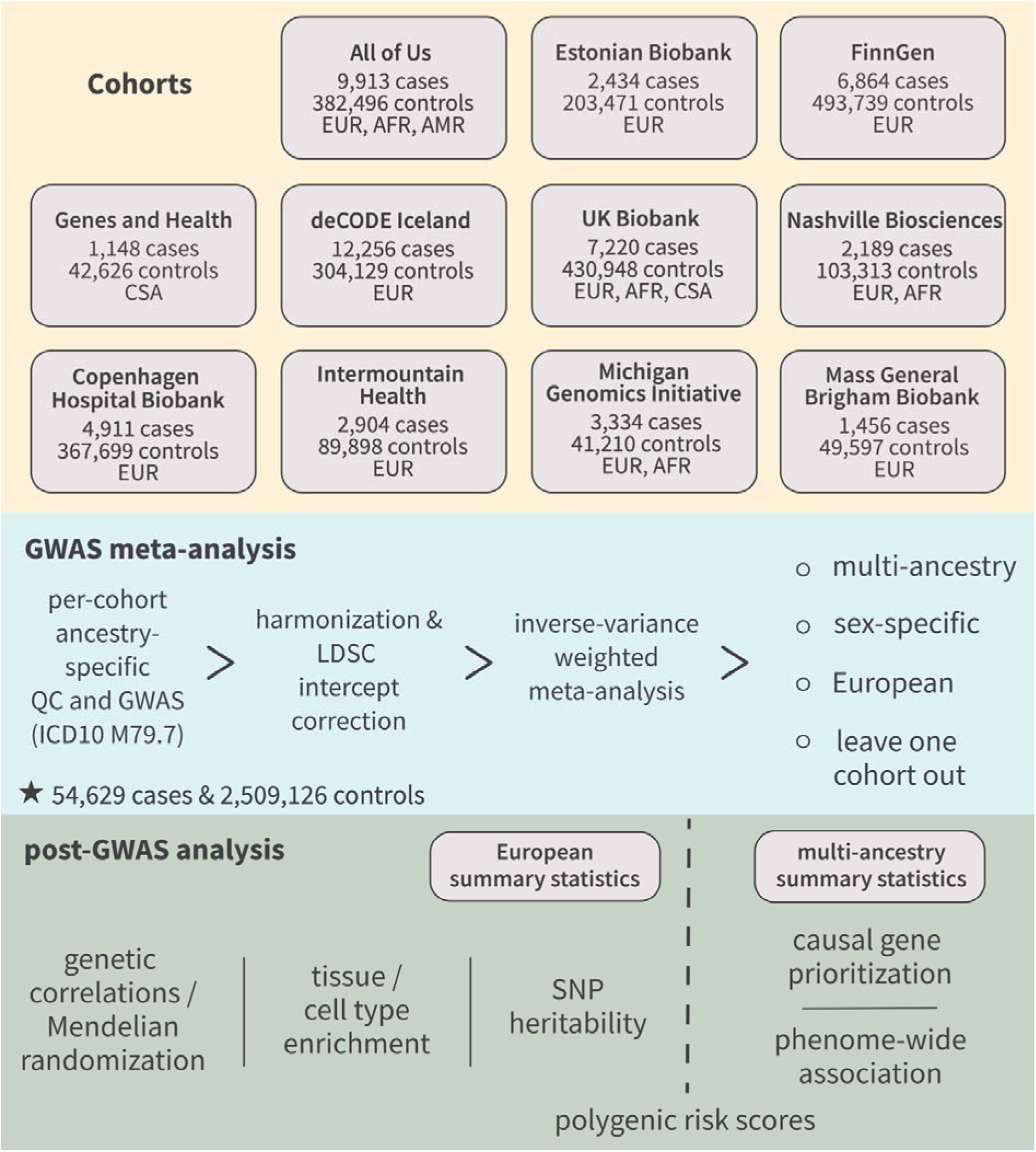
Study overview. “Copenhagen Hospital Biobank” is short for “Copenhagen Hospital Biobank and Danish Blood Donor Study”.

Our primary meta-analysis encompassed 54,629 fibromyalgia cases and 2,509,126 controls, for a total of 2,563,755 individuals (Table S1). This represents a hundred-fold increase in sample size over the previous largest fibromyalgia GWAS^38^. We found 26 independent genome-wide-significant (p < 5 × 10^-8^) risk loci for fibromyalgia (Table 1, Figure 2), with largely consistent effect sizes across cohorts (Figure S1).

**Table 1:**
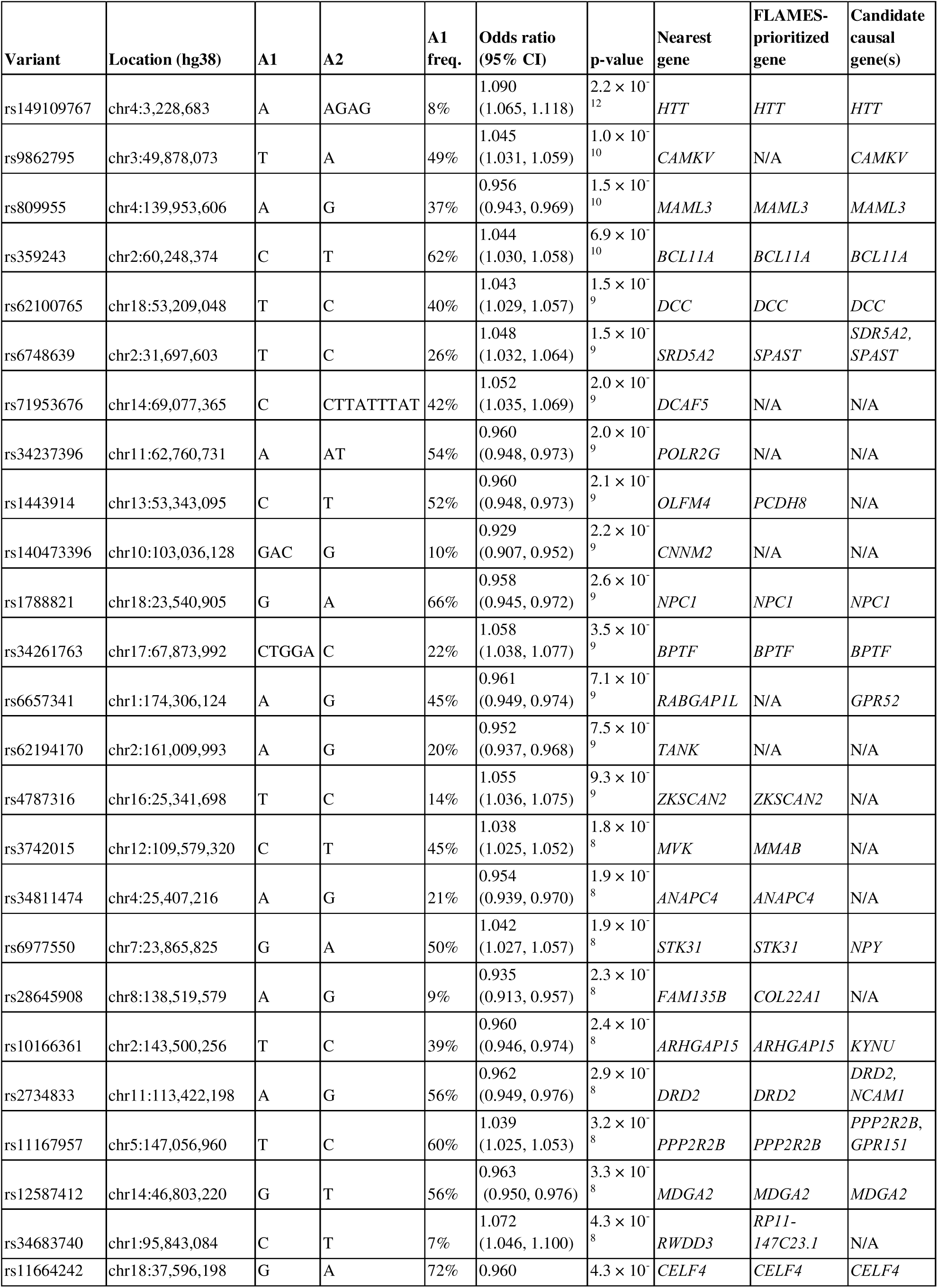
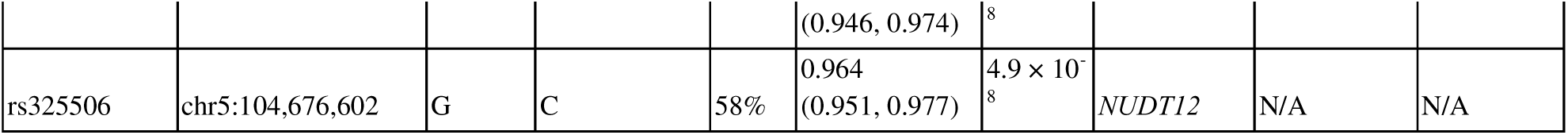
Lead variants and candidate causal genes from the primary meta-analysis. freq. = frequency.

**Figure 2:**
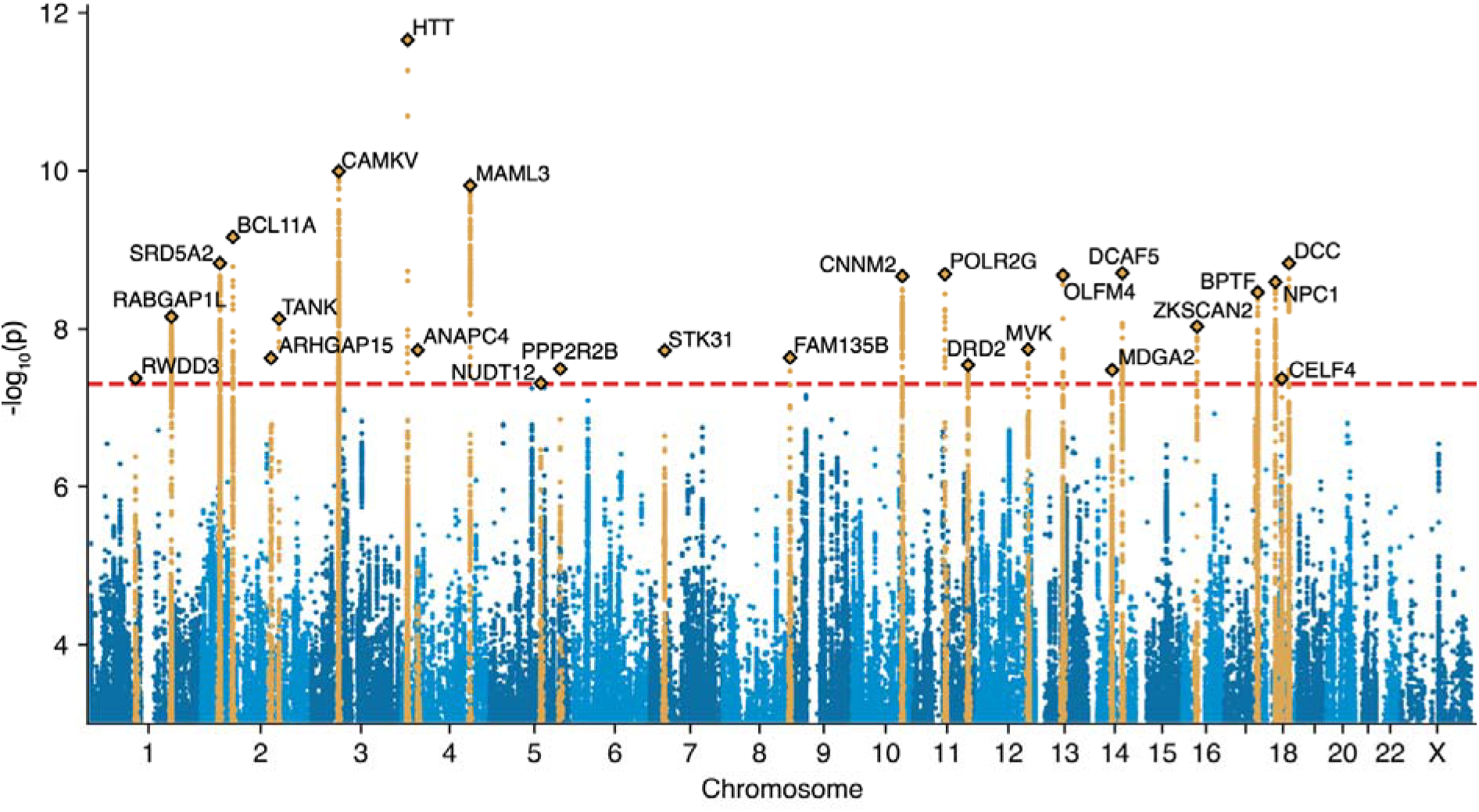
Manhattan plot of the primary meta-analysis. Gold-highlighted variants have linkage disequilibrium r^2^ > 0.001 and are within 5 megabases of the lead variants (diamonds). The nearest gene to each lead variant is labeled.

We also performed multiple secondary meta-analyses (Figure 1). First, to enable genetic risk prediction in our 11 cohorts without sample overlap bias, we performed leave-one-cohort-out meta-analyses for each cohort. Second, we noted that 49,000 cases (89.7%) were of European ancestry. To reduce bias in analyses that rely on European LD reference panels – like heritability, genetic correlation, and tissue/cell-type enrichment – we performed European-only meta-analyses (Figure S2).

Consistent with known sex differences in fibromyalgia prevalence^45,46^, the vast majority of our 54,629 cases (47,895 cases, 87.7%) were female. We therefore conducted sex-stratified meta-analyses to determine if the genetic architecture of the disorder differs between sexes (Figure S3, Figure S4, Table S2). While the lower number of male cases yielded less statistical power (2 genome-wide significant associations in males versus 21 in females), multiple lines of evidence point to a shared genetic architecture. The inter-sex genetic correlation in European-ancestry individuals was 1.03 (statistically indistinguishable from perfect correlation), and the odds ratios for the 26 lead variants from the primary meta-analysis did not differ significantly between males and females (Figure S5). These results suggest the genetic architecture of fibromyalgia is largely consistent between sexes.

We prioritized causal genes at the 26 significant loci from the primary meta-analysis. We performed manual literature review of the nearest protein-coding gene and other notable genes at the locus, augmented by an annotation pipeline from deCODE genetics that considers quantitative trait loci and coding variants in strong LD with our lead variants (Table S3, Methods) and the machine-learning-based GWAS causal gene prioritization method FLAMES^47^. We discuss particularly notable prioritizations here. Many, though not all, have strong links to brain function or have previously been associated with brain disease.

### The top fibromyalgia association is with a coding variant in the Huntingtin gene

The most significant association in the combined ancestry analysis was with rs149109767-A (OR = 1.09, p = 2.2 × 10^-12^), an inframe glutamic acid deletion in the Huntingtin (*HTT*) gene. *HTT*, the causal gene for Huntington’s disease (HD), is ubiquitously expressed and involved in diverse cellular processes including signaling, transcriptional regulation, cell division, endocytosis, autophagy, and axonal transport^48–50^, and is required for healthy neurodevelopment^51^. *HTT* contains a trinucleotide CAG repeat region in exon 1, where expansions of ≥36 repeats cause HD^52^. Our fibromyalgia variant is located in exon 58 of the large *HTT* gene (which spans 67 exons and 180 kb) and does not lie in this repeat region, but is part of the Huntington’s disease-associated “A1” haplotype found in Europeans^53^. This variant has the largest effect size of all 26 lead variants: carriers have about 9% increased odds of fibromyalgia.

We next investigated whether our fibromyalgia variant is associated with HD using a 6 European-cohort HD GWAS meta-analysis available at deCODE (N_case/ctrl_ = 184/1,198,665). The top HD association (rs71180116-CCAGCAGCAGCAGCAGCAGCAGCAGCAGCAGCAGCAG, OR = 44.26, p = 2.62 × 10^-30^) was not in strong linkage disequilibrium with our lead fibromyalgia variant (r^2^ = 0.0055, D’ = 0.258) and showed no association with fibromyalgia (OR = 1.027, p = 0.41). While our fibromyalgia variant rs149109767-A did associate with HD risk (OR = 2.118, p = 0.00354), this association disappeared in the UK Biobank after conditioning on rs71180116 (OR_adj_ = 1.328, p = 0.297), an effect replicated in the smaller Icelandic GWAS.

We found a second association with a possible HD link: rs6657341-A (OR = 0.961, p = 7.1 × 10^-9^), an intronic variant in *RABGAP1L*, ∼150 kilobases upstream of its second-nearest gene *GPR52*. *RABGAP1L* regulates *RAB* proteins, which direct intracellular trafficking^54,55^, but has no clear link to fibromyalgia. Conversely, *GPR52* is a brain-specific orphan G-protein coupled receptor being investigated as a drug target for HD as it regulates HTT levels^56–59^.

### Additional associations implicate pathways in pain processing

Several associations implicated pathways relevant to pain processing. One notable finding involved rs11664242, an intergenic variant ∼30 kilobases upstream of its nearest coding gene *CELF4*. *CELF4* is an RNA-binding protein and translational regulator predominantly expressed in excitatory neurons, regulating synaptogenesis during neurodevelopment by exerting broad-spectrum effects on the translation of mRNAs with synaptic roles^60,61^. CELF4 reduces pain and sensory sensitivity by negatively regulating nociceptor excitability^62^, and *CELF4* gene therapy is being explored as a therapeutic strategy for chronic pain.

Another association was found with rs6977550, an intergenic variant ∼30 kilobases downstream of its nearest coding gene *STK31*. *STK31* encodes a testis-specific protein kinase with unclear relevance to fibromyalgia; however, the variant is ∼400 kilobases upstream of *NPY* (its sixth-nearest gene), an abundant neuropeptide that modulates appetite, circadian rhythm, anxiety, cardiovascular function, and immune responses^63^ and has both pro-and anti-nociceptive effects^64^.

We also identified rs10166361, an intronic variant in *ARHGAP15* which encodes a Rho GTPase-activating protein that regulates neutrophils^65^ and cellular functions such as cytoskeletal organization^66^. In mouse models, *ARHGAP15* knockout impairs cortical interneuron excitability (causing subclinical seizures) and hippocampal function (causing memory impairment)^66–68^. The variant is also ∼450 kilobases downstream of *KYNU* (the third-nearest gene), which encodes kynureninase, an enzyme that cleaves kynurenine into anthranilic acid^69^. Kynurenine and its metabolites are involved in pain processing, and the kynurenine pathway has been proposed as a therapeutic target for pain^70,71^. Gene-based tests have linked *KYNU* to depressive symptoms, which are often comorbid with chronic pain^72^.

### Other significant associations point to brain-related mechanisms

We found additional brain related associations. One particularly notable hit was rs2734833, an intronic variant in *DRD2*, a locus with pervasive psychiatric and sleep associations. *DRD2* encodes the dopamine D2 receptor, which influences motivation and cognition^73^ and is the primary target of most antipsychotic drugs. In addition, *DRD2* was one of the first loci discovered to modulate sleep duration^74,75^. The variant lies ∼150 kilobases downstream of *NCAM1,* the fourth-nearest gene, which encodes a cell surface receptor (also known as CD56) with both nervous and immune roles. In the nervous system, it regulates neural cell adhesion, neuronal migration, neurite outgrowth, axon guidance, and synaptic plasticity^76–78^, and plays a role in memory^79^. In the immune system, CD56 is the primary marker for natural killer (NK) cells and distinguishes their two main subtypes. It helps NK cells attach to target cells and trigger the release of cytotoxic granules that kill target cells^80^.

Several other associations also highlight neurodevelopmental pathways. The second-most significant association was with rs9862795, an intergenic variant located ∼8 kilobases upstream from *CAMKV*. *CAMKV* encodes a pseudokinase from the Ca²L/calmodulin-dependent protein kinase family that is predominantly brain-expressed and critical for dendritic spine maintenance, synaptic transmission and synaptic plasticity^81,82^. The lead variant is also in strong LD (r^2^ = 0.905; Table S3) with two variants in a pleiotropic tyrosine kinase receptor, *MST1R*.

Two other lead variants were rs809955, an intronic variant in *MAML3*, and rs12587412, an intergenic variant ∼40 kilobases downstream of its nearest coding gene *MDGA2*. *MAML3* encodes a transcriptional coactivator in the Notch signaling pathway^83^. Notch signaling is vital for development and cell-fate decisions: in development, inhibition promotes neural differentiation, while in adulthood, activation influences neural stem cell regulation, neuronal survival, migration, and synaptic plasticity^84,85^. *MDGA2* is predominantly expressed in the brain and regulates synaptic organization^86^, axonal migration^87^, BDNF/TrkB signaling^88^, and excitatory neurotransmission^86^. *MDGA2* haploinsufficiency is associated with autism spectrum disorder^88,89^.

Another neurodevelopmental association was with rs62100765, an intronic variant in *DCC* which was the sole coding gene within 1 megabase. *DCC* encodes a transmembrane receptor that guides midline-crossing axons during neurodevelopment^90^ and regulates myelin structure and axon domain organization^91^.

We also found an association with rs6748639, an intergenic variant located ∼100 kilobases upstream of the nearest coding gene, *SRD5A2*, and ∼350 kilobases upstream of *SPAST*, the fifth-nearest gene and the one prioritized by FLAMES. *SRD5A2* is one of three enzymes that converts testosterone to the more potent androgen dihydrotestosterone, and is critical to normal male sexual development: biallelic loss causes 5α-reductase deficiency, with female-like external genitalia at birth^92^. Although the role of testosterone in pain remains controversial, some studies have reported inverse associations between testosterone levels and pain perception^93–95^. An alternative causal gene candidate at this locus, *SPAST,* encodes the neuronally expressed protein spastin, which regulates microtubule length and disassembly^96^; haploinsufficiency is the most common cause of hereditary spastic paraplegia^97^.

Several associations implicated genes linked to intellectual disability and developmental disorders. One was with rs359243, an intergenic variant located in a gene dense region ∼200 kilobases downstream from its nearest coding gene, the transcription factor *BCL11A*. Besides its canonical role in hematopoiesis and repression of fetal hemoglobin in adults, *BCL11A* has neurodevelopmental roles, and haploinsufficiency causes intellectual disability and autism^98–100^.

Another developmental disorder association was with rs34261763, an intronic variant in *BPTF*, which encodes a subunit of the developmentally essential NURF chromatin remodeling complex^101^. *BPTF* haploinsufficiency causes a neurodevelopmental disorder with intellectual disability, developmental and speech delay, microcephaly, and facial and limb dysmorphia^102^. The lead variant at this locus is in strong LD (r^2^ = 0.831; Table S3) with the poorly characterized anti-apoptotic gene^103^ *C17orf58*.

We also found an association with rs11167957, an intronic variant in *PPP2R2B*, a brain-enriched subunit of the serine/threonine phosphatase PP2A^104^. PP2A regulates cell proliferation and apoptosis, DNA replication, transcription, translation, and signal transduction^105^. CAG repeat expansion in *PPP2R2B* causes spinocerebellar ataxia 12, an autosomal dominant speech and motor disorder^106^, and *PPP2R2B* missense variants are linked to intellectual disability with developmental delay^104^. The variant is also ∼550 kilobases upstream of the orphan brain-specific G protein-coupled receptor *GPR151* (its 5th-nearest gene), which mediates neuropathic pain via effects on microglial activation and neuroinflammation^107,108^.

Lastly we found an association with rs1788821, an intronic variant in *NPC1*. *NPC1* encodes a transmembrane protein vital for intracellular cholesterol trafficking^109^, and biallelic loss of NPC1 function causes Niemann-Pick disease type C1^110^, characterized by lysosomal lipid accumulation, neuroinflammation, abnormal dendrite growth, neurofibrillary tangles, and neuroaxonal dystrophy^111^. The lead variant is in strong LD with missense variants in *NPC1* and an autophagy-related gene^112^, *RMC1* (r^2^ = 0.998 and 0.990, respectively; Table S3).

### Phenome-wide associations of lead variants point to substantial pleiotropy

To identify previously reported associations for our 26 lead variants, we cross-referenced them and their high-LD proxies (r² > 0.8) with the GWAS Catalog. This analysis revealed 20 traits associated with at least four of the 26 lead variants (Figure 3 top, Table S4). The most frequent association was with educational attainment (10 variants), followed by body mass index (8 variants), type 2 diabetes (7 variants), pain intensity (6 variants), and insomnia (6 variants). Among the notable psychiatric associations were 2 with depression, 4 with depressive symptoms (with one association overlapping with depression), 3 with schizophrenia, and 1 with suicide attempt. The remaining associations encompassed a broad range of categories, including additional pain, immune and behavioral traits (e.g. multisite chronic pain, white blood cell count, neuroticism), and anthropometric and metabolic traits (e.g. weight, visceral adipose tissue, metabolic syndrome).

**Figure 3:**
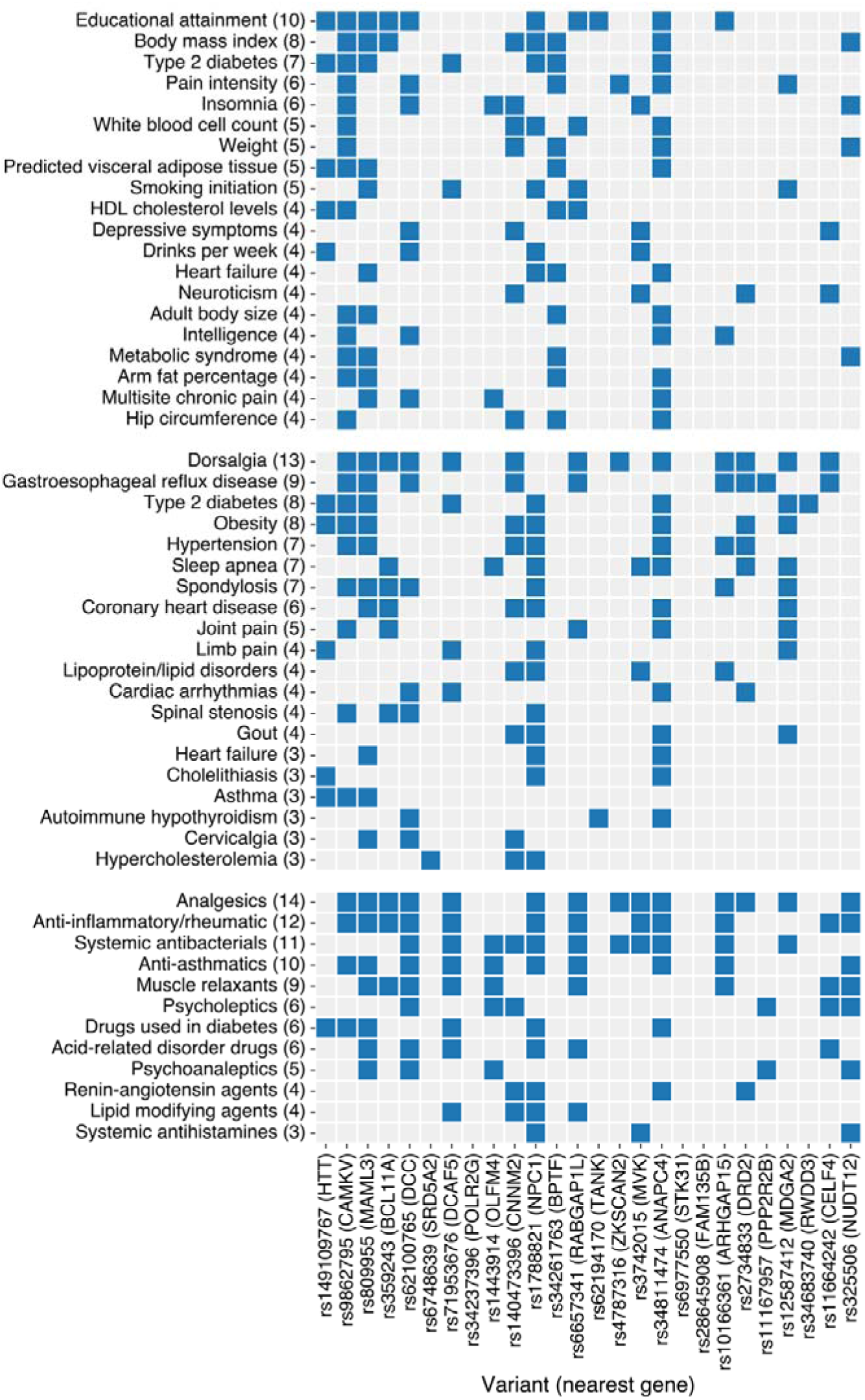
Phenome-wide associations of lead variants. Each of the 26 lead variants from the primary meta-analysis was tested for overlap with genome-wide significant variants from the GWAS Catalog, either with the lead variant itself or variants with r^2^ > 0.8 (top). The 26 lead variants were also tested for association with 330 diseases in a meta-analysis of the Million Veteran Program, FinnGen and the UK Biobank (middle), and 124 drug classes in FinnGen (bottom), applying Bonferroni correction across the number of variants and diseases/drug classes tested. The number of variants (out of 26) significantly associated with each disease or drug is listed in brackets; for brevity, only diseases or drugs associated with at least 10% of the lead variants (i.e. ≥3 of 26) are shown, or 15% (i.e. ≥4 of 26) for the GWAS Catalog analysis, with full results listed in Table S4-S6. Only lead variants with at least one significant phenome-wide association are shown.

We also performed a phenome-wide association study (PheWAS) in which the 26 lead variants were tested for association with 330 diseases, leveraging pre-existing GWAS meta-analyses of the Million Veteran Program, FinnGen and the UK Biobank (“MVP-Finngen-UKBB”). After applying Bonferroni correction across the 330 diseases and 26 variants tested, 20 diseases were associated with three or more lead variants (Figure 3 middle, Table S5). These diseases largely fell into three categories: pain and pain-associated traits (dorsalgia, spondylosis, joint pain, limb pain, spinal stenosis, cervicalgia), metabolic syndrome and its comorbidities (type 2 diabetes, obesity, sleep apnea, hypertension, coronary heart disease, gout, lipoprotein/lipid disorders, hypercholesterolemia, cholelithiasis), and immune disease (autoimmune hypothyroidism, asthma). The most common association was dorsalgia (also known as back pain, 13 of 26 variants associated); the second-most common, strikingly, was gastroesophageal reflux disease (9 variants).

Applying the same approach to 124 drug prescription GWAS from FinnGen (Figure 3 bottom, Table S6), we found the most frequent associations were with analgesics (14 variants) and anti-inflammatory/rheumatic drugs (12 variants). Notably, 11 variants were associated with both categories, likely reflecting the prescription of non-steroidal anti-inflammatory drugs (included in both categories) for pain. A further 10 drug categories were associated with at least three lead variants, including psychoactive drugs (muscle relaxants, psycholeptics, psychoanaleptics), treatments for metabolic syndrome and its comorbidities (diabetes drugs, lipid modifying agents, renin-angiotensin agents), and immune modulators (anti-asthmatics, anti-histamines).

Overall, these results demonstrate that the top genetic risk variants for fibromyalgia are highly pleiotropic, broadly influencing risk for many of the disorder’s most common comorbidities and likelihood of prescription of the drugs used to treat them.

### Fibromyalgia heritability is exclusively enriched in neural tissues and cell types

To identify the cellular context of fibromyalgia genetic risk, we used LD score regression applied to specifically expressed genes (LDSC-SEG)^113^ to quantify fibromyalgia heritability enrichment near genes with enriched expression in each of 53 tissues in the Genotype-Tissue Expression (GTEx) project, and each of 119 cell types in the ∼20 million whole-mouse PanSci single-cell atlas (Figure 4, Table S7, Table S8). The analysis revealed a strong and exclusive enrichment within the nervous system: all 5 GTEx tissues with Bonferroni-significant enrichments were brain regions (cortex, caudate, frontal cortex, putamen, and anterior cingulate cortex) and 12 of 13 enriched cell types were neuronal.

**Figure 4:**
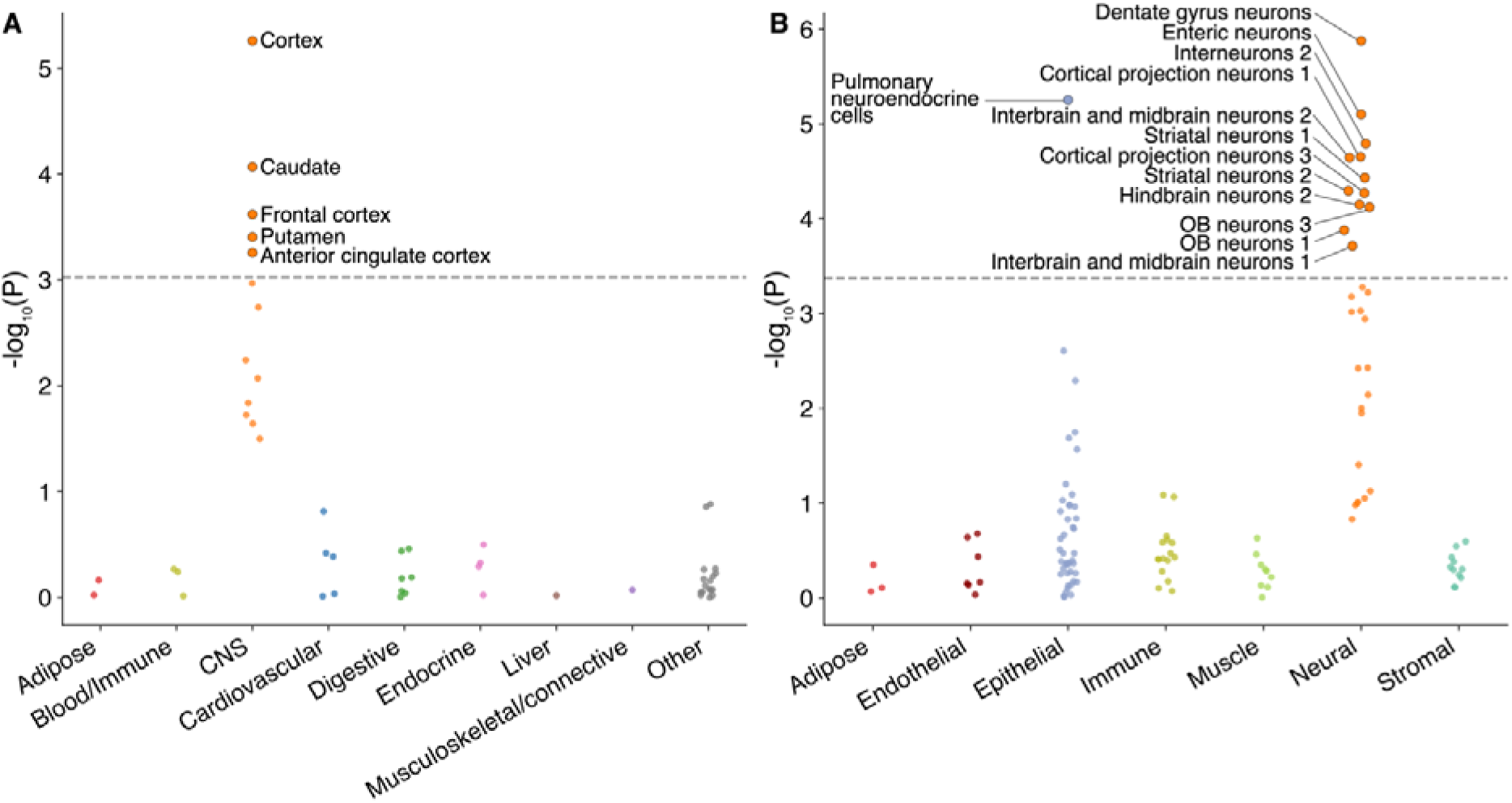
Tissue and cell-type enrichments. Fibromyalgia heritability enrichments among variants inside or within 100 kilobases of genes with enriched expression in a) tissues in the Genotype-Tissue Expression (GTEx) project, and b) cell types in PanSci, a ∼20 million whole-mouse single-cell atlas. Results are shown as negative log p-values for enrichment, grouped and colored by tissue group (GTEx) or cell lineage (PanSci), with random side-to-side jitter for visibility. Bonferroni-significant tissues and cell types are labeled. Full results are listed in Table S7-S9.

The strongest cell-type association was with neurons from the dentate gyrus (p = 1.3 × 10^-6^), a hippocampal region critical for contextualizing sensory experiences, including pain. Significant enrichments were also found in enteric neurons (p = 7.9 × 10^-6^) and diverse interneurons, cortical projection neurons, and striatal neurons. The sole enriched non-neuronal cell type, pulmonary neuroendocrine cells (p = 5.6 × 10^-6^), is a specialized sensory cell type with well-established neuron-like properties. Even when grouping cell types by lineage with the goal of increasing power, the only significant lineage was neural (p = 2.3 × 10^-4^; p > 0.05 for all other cell types; Table S9).

Collectively, these results implicate a broad network of neurons across the central and peripheral nervous systems, including sensory-processing neurons, in the genetic etiology of fibromyalgia. There was no evidence of enrichment in non-neural tissues or cell types, including immune ones.

### Pervasive genetic correlations with comorbid disorders

We computed genetic correlations between our European-only meta-analysis results and each of 855 diseases in FinnGen, resulting in 337 Bonferroni-significant genetic correlations (Figure 5, Table S10).

**Figure 5:**
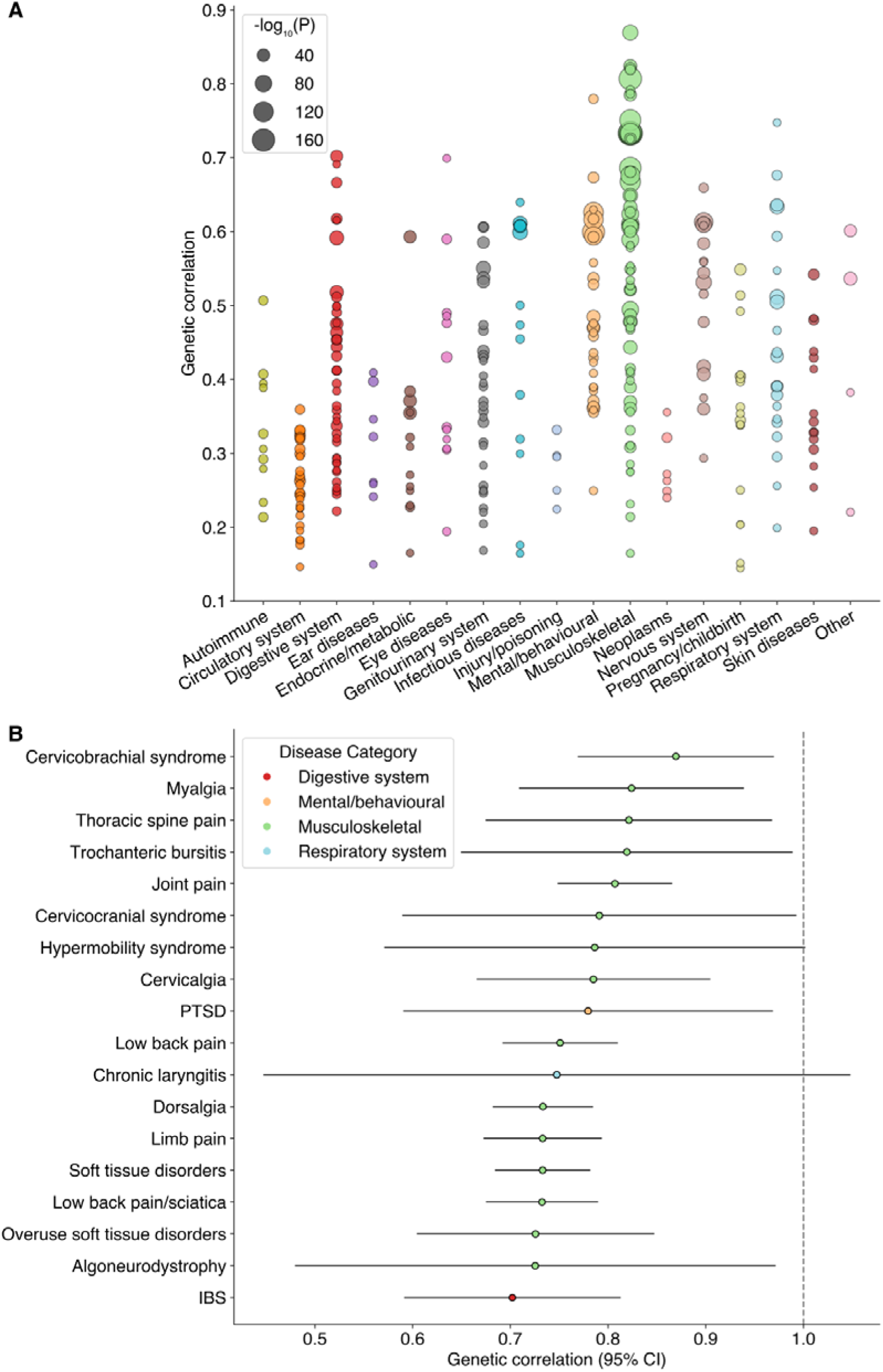
Genetic correlations between fibromyalgia and FinnGen disease endpoints. a) Genetic correlations of the 337 disease endpoints with Bonferroni-significant genetic correlations with fibromyalgia, grouped by disease area. The area of each circle is proportional to the logarithm of its genetic correlation p-value. b) Genetic correlations of the 18 diseases with genetic correlation greater than 0.7; error bars denote 95% confidence intervals. Full results are listed in Table S10.

As anticipated, the most extensive and significant genetic overlap was with musculoskeletal and pain disorders. The strongest signals arose from conditions characterized by widespread or syndromic pain, including cervicobrachial syndrome (r_g_ = 0.87, p = 9.2 × 10^-65^), myalgia (r_g_ = 0.82, p = 9.4 × 10^-45^), and soft tissue disorders (r_g_ = 0.73, p = 5.0 × 10^-190^). We also observed substantial correlations for a range of common localized pain presentations, from joint pain (r_g_ = 0.81, p = 1.7 × 10^-160^) and low back pain (r_g_ = 0.75, p = 1.1 × 10^-137^) to migraine (r_g_ = 0.61, p = 2.2 × 10^-73^). Furthermore, this genetic overlap extended to related musculoskeletal conditions like trochanteric bursitis (r_g_ = 0.82, p = 3.1 × 10^-21^) and hypermobility syndrome (r_g_ = 0.79, p = 8.4 × 10^-13^).

Fibromyalgia also displayed substantial genetic correlations with a spectrum of mental and behavioral disorders, with a magnitude for some traits that rivaled those of pain conditions. The most notable of these was with post-traumatic stress disorder (r_g_ = 0.78, p = 6.6 × 10^-16^). Strong correlations were also evident for dissociative disorders (r_g_ = 0.63, p = 5.0 × 10^-6^), somatoform disorder (r_g_ = 0.67, p = 8.8 × 10^-26^), and depression (r_g_ = 0.63, p = 4.1 × 10^-118^), alongside more modest correlations with generalized anxiety disorder (r_g_ = 0.46, p = 3.8 × 10^-20^) and insomnia (r_g_ = 0.47, p = 2.0 × 10^-43^). Beyond specific diagnoses, we found a strong correlation with the core clinical symptom of malaise and fatigue (r_g_ = 0.60, p = 8.0 × 10^-36^).

Significant genetic overlap extended to other commonly co-occurring conditions, including disorders of the digestive, genitourinary, and respiratory systems. Among digestive disorders, we observed strong signals for irritable bowel syndrome (r_g_ = 0.70, p = 2.1 × 10^-35^) and functional dyspepsia (r_g_ = 0.67, p = 9.1 × 10^-24^). We also identified correlation with genitourinary conditions, such as polycystic ovarian syndrome (r_g_ = 0.59, p = 8.3 × 10^-39^), as well as with the respiratory condition chronic laryngitis (r_g_ = 0.75, p = 1.1 × 10^-6^).

Genetic correlations with autoimmune disorders were relatively modest, especially when compared to the strong signals observed for many of the conditions above. The highest correlations in this category were with psoriatic arthropathies (r_g_ = 0.41, p = 5.4 × 10^-17^) and Sjögren’s syndrome (r_g_ = 0.39, p = 1.7 × 10^-6^). We also observed significant correlations for other rheumatological conditions such as rheumatoid arthritis (r_g_ = 0.33, p = 3.1 × 10^-16^), as well as for psoriasis (r_g_ = 0.29, p = 6.0 × 10^-15^) and autoimmune hypothyroidism (r_g_ = 0.21, p = 4.2 × 10^-17^).

Given the genetic correlations observed with asthma (r_g_ = 0.51, p = 2.5 × 10^-57^) and rheumatoid arthritis (RA) and the longstanding debate on whether fibromyalgia is an inflammatory or autoimmune disease^9^, we explored genetic correlations with defined subtypes of these diseases. Asthma can be broadly classified as T2-high or T2-low, defined by high or low levels of eosinophils and other T2 molecules, with approximately half of individuals with asthma falling into each category^114,115^. While the mechanisms driving T2-high (typical) asthma are increasingly understood, much less is known about the underlying mechanisms that drive T2-low asthma^114,115^. Using GWAS results from T2-high and T2-low asthma at deCODE^116^, we found that the genetic correlation with fibromyalgia is stronger with T2-low asthma (r_g_ = 0.44, p = 3.5 × 10^-24^) than T2-high asthma (r_g_ = 0.27, p = 7.4 × 10^-14^), consistent with the lack of genetic correlation between eosinophil levels and fibromyalgia (r_g_ = 0.04, p = 0.1).

Similarly, RA is classified as seronegative or seropositive based on the absence or presence of anticitrullinated protein antibodies (ACPA) and rheumatoid factor (RF). While seronegative RA is considered less severe than seropositive RA, recent studies show that both types can have severe consequences and should be actively treated^117^. We found that fibromyalgia is more genetically correlated with the seronegative form of RA (r_g_ = 0.41, p = 2.7 × 10^-15^), compared to seropositive RA (r_g_ = 0.24, p = 3.0 × 10^-10^), an association that has also been observed clinically^118^. As is the case for T2-low asthma, less is known about the genetic and environmental risk factors for seronegative RA compared to the seropositive subset of this criteria-based syndrome^119^ and the seronegative subset is likely to have a more heterogeneous background. These subtype analyses of asthma and RA suggest that fibromyalgia is genetically correlated with the more heterogeneous and less well-defined pathobiological forms of these diseases, further distinguishing the genetic architecture of fibromyalgia from that of well-characterized, peripherally driven inflammatory and autoimmune diseases.

Collectively, this landscape of genetic correlations further highlights the profound pleiotropy of fibromyalgia genetic risk factors. This apparent paradox – widespread somatic correlations despite a genetic architecture strongly enriched in neuronal cell types – is resolved when considering that many of the above-mentioned conditions are themselves rooted in the central nervous system. This could point toward a partially shared, centrally-mediated genetic etiology as the key driver for their frequent co-occurrence with fibromyalgia.

### Genetic risk prediction is modestly effective at patient stratification

To enable fibromyalgia genetic risk prediction, we constructed polygenic risk scores (PRSs) for the European and multi-ancestry leave-UK Biobank-out meta-analyses. PRS weights will be made available via the PGS Catalog (see Data Availability).

We evaluated the performance of the multi-ancestry leave-UK Biobank-out PRS within the independent UK Biobank cohort. The PRS showed modest predictive ability, commensurate with its modest observed-scale single nucleotide polymorphism-based heritability of 10.4% (95% confidence interval 9.8% to 11.0%). Performance was highest among European-ancestry UK Biobank participants (area under the receiver-operating curve [AUC] = 0.59), with attenuated South Asian (AUC = 0.55) and African (AUC = 0.55) accuracies (Figure S6A), reflecting the mostly European composition of our samples.

Despite modest AUCs, the PRS effectively stratified individuals by risk, particularly in Europeans where power and predictive accuracy were greatest. Compared to the middle quintile, European-ancestry participants in the highest PRS quintile had a fibromyalgia odds ratio of 1.5 (95% confidence interval 1.4 to 1.7), while those in the lowest quintile had an odds ratio of 0.63 (95% confidence interval 0.57 to 0.68); Figure S6B). This risk stratification translated to marked differences in disease prevalence between the lowest and highest PRS quintiles (Figure S6C): among European-ancestry participants, prevalence increased from approximately 1.0% in the lowest quintile to 2.4% in the highest. The PRS weights derived from the European-only summary statistics performed similarly on the European UK Biobank cohort.

## Discussion

In this GWAS of over 2.5 million individuals, we identified 26 genetic risk loci for fibromyalgia, providing the first insights into its genetic architecture. These findings establish a firm biological basis for a condition long defined solely by its clinical symptoms, and whose validity remains debated in some circles.

Our tissue and cell-type analyses revealed that fibromyalgia heritability is enriched in the brain and in neuronal cell types. This is consistent with the central sensitization model of fibromyalgia, in which the central nervous system develops heightened responsiveness to pain and other sensory stimuli. The specific genes implicated at our risk loci further refine this model. We identified multiple causal genes candidates like *HTT*, *CAMKV*, *DCC*, *MDGA2*, and *CELF4* that participate in central nervous system processes like neurodevelopment and neuronal maintenance. This neural basis is reflected in the extensive overlap of our lead variants (or high-LD proxies) with those of other brain-related traits in our GWAS Catalog and MVP-Finngen-UKBB PheWAS analyses. These included pain traits like multisite chronic pain (*MAML3*, *DCC*, *OLFM4*, *ANAPC4*) and dorsalgia (*CAMKV*, *MAML3*, *BCL11A*, *DCC*, *DCAF5*, *CNNM2*, *RABGAP1L*, *ZKSCAN2*, *ANAPC4*, *ARHGAP15*, *DRD2*, *MDGA2*, *CELF4*), sleep traits like insomnia (*CAMKV*, *DCC*, *OLFM4*, *CNNM2*, *MVK*, *NUDT12*), and canonical psychiatric disorders like depression (*DCC*, *NUDT12)* and schizophrenia (*CNNM2*, *STK31, PPP2R2B*). That said, some brain-related traits have much stronger genetic correlations with fibromyalgia than others: for instance, pain traits and post-traumatic stress disorder have higher correlations than psychiatric disorders like depression and schizophrenia, which in turn have higher genetic correlations than sleep traits like insomnia.

The genetic underpinnings of fibromyalgia are shared with a remarkably wide variety of disease types. Besides brain-related traits, genetic correlations implicate the digestive, genitourinary, and respiratory systems. Additionally, certain risk loci overlapped with long COVID^120^ (*BPTF*) and ME/CFS^121^ (*OLFM4*, *RABGAP1L/GPR52*), two poorly characterized disorders, albeit with different lead variants. These findings could point towards genetic factors associated with comorbidities and/or clinically misdiagnosed conditions. However, another plausible explanation of this widespread pleiotropy is that fibromyalgia genetics capture a core, transdiagnostic vulnerability. Such shared genetic architecture, rooted in central nervous system function, could predispose individuals to a spectrum of conditions characterized by sensory and/or affective dysregulation, which may manifest clinically as fibromyalgia, irritable bowel syndrome, post-traumatic stress disorder, or a constellation of other disorders, depending on other genetic and environmental factors.

There is uncertainty about whether fibromyalgia is an autoimmune disorder^9^. We did not find a significant GWAS signal in the MHC region or heritability enrichment in immune cell types, but did observe significant, albeit weak, genetic correlations between fibromyalgia and autoimmune disorders including Sjögren’s syndrome and RA. These associations should be interpreted with caution in light of the potential for diagnostic misclassification of fibromyalgia as Sjögren’s syndrome or RA^122,123^. For example, studies in the US have suggested that only about 12 to 33 percent of individuals with self-reported RA actually meet diagnostic criteria for RA^124^. Further supporting a divergence between fibromyalgia and classical autoimmune disorders, we found that fibromyalgia had stronger genetic correlation with the more atypical seronegative subtype of RA than with its canonical seropositive subtype. However, we did discover GWAS Catalog associations with other immune traits, including 5 loci that associate with white blood cell count. Overall, our results suggest that fibromyalgia is not primarily an autoimmune disorder, though it may have an immune component.

Our study directly addresses the sex bias in fibromyalgia prevalence. Despite several-fold higher prevalence in females, we found no sex difference in the genetic architecture of fibromyalgia. The observed difference in prevalence is therefore unlikely to be driven by sex-specific risk variants, and is more likely attributable to other biological factors (e.g. hormonal influence), environmental exposure, or their interaction with a shared genetic predisposition.

This work not only defines a genetic basis for fibromyalgia but also identifies potential therapeutic targets. The linked associations of *HTT* and its regulator *GPR52* highlight a clear repurposing opportunity, as *GPR52* is already an investigational drug target for Huntington’s disease. Furthermore, our identification of *CELF4* as a fibromyalgia risk locus provides a direct genetic rationale for exploring *CELF4*-based gene therapies – already under investigation for chronic pain – for fibromyalgia in particular. These loci represent the first genetically-supported molecular targets for fibromyalgia, providing tangible starting points for developing mechanism-based treatments.

This study has important limitations. A primary limitation is its reliance on ICD codes from electronic health records for case ascertainment. This may result in the inclusion of individuals not meeting full syndromic criteria for fibromyalgia, or the exclusion of undiagnosed cases, but is necessary to achieve the massive sample size required for genetic discovery. We also avoided excluding controls with conditions like depression or back pain, since this would create an artificially “super-healthy” control group, biasing our results toward the genetics of those comorbidities rather than the core fibromyalgia phenotype. Lastly, our cohort was of mostly European ancestry, which limits the generalizability of our findings and transferability of our polygenic risk scores to non-European ancestries.

In summary, our large-scale multi-ancestry GWAS provides a map of the genetic architecture of fibromyalgia, identifying 26 risk loci and providing robust genetic validation of the notion that fibromyalgia is primarily a central nervous system disorder. The identification of specific risk loci provides the field with concrete molecular starting points, enabling hypothesis-driven research to dissect the biological basis of its pathophysiology and its shared etiology with comorbid conditions.

## Data availability

GWAS summary statistics will be distributed via the GWAS Catalog (https://www.ebi.ac.uk/gwas) upon publication, and polygenic risk scores via the Polygenic Score (PGS) Catalog (https://www.pgscatalog.org).

This study used data from the *All of Us* Research Program’s Controlled Tier Dataset Curated Data Repository version 8, available to authorized users on the Researcher Workbench.

## Acknowledgments

We gratefully thank the All of Us, Estonian Biobank, FinnGen, Genes and Health, deCODE Iceland, UK Biobank, Nashville Biosciences, Copenhagen Hospital and Danish Blood Donor Study, Intermountain Health, Michigan Genomics Initiative and Mass General Brigham Biobank participants.

We acknowledge funding and institutional support from multiple sources. Canadian contributions were supported by the Canadian Institutes of Health Research (CIHR FBD-199459; CIHR MHP-192163) and by the Natural Sciences and Engineering Research Council of Canada (NSERC) Undergraduate Student Research Award (URSA). In the United States, support was provided by the Michigan Genomics Initiative and AI & Digital Innovation at the University of Michigan, Ann Arbor, MI. S.F. is funded by an NIH K08 Clinical Investigator Award (K08AR082454). Further support from the United States included NIH funding to R.S. and NIH RM1HG010461 and a generous donation to N.S.A..

In Denmark, the Copenhagen Hospital Biobank (CHB) was supported by the Department of Clinical Immunology, Rigshospitalet, Copenhagen University Hospital; by grants from the Novo Nordisk Foundation (NNF23OC0082015, NNF17OC0027594); and by the Rigshospitalet Research Council (Framework grant). The Danish Blood Donor Study (DBDS) is funded by an annual grant from Bio-and Genome Bank Denmark, with additional support from the Danish Administrative Regions (02/2611) and the Danish Council for Independent Research (09–069412), as well as the Novo Nordisk Foundation (NNF23OC0082015, NNF17OC0027864, NNF17OC0027594). The Parker Institute is supported by a core grant from the Oak Foundation (OFIL-24-074). Further support in Denmark was received from the Novo Nordisk Foundation (NNF17OC0027594, NNF14CC0001). In the United Kingdom, support was provided to F.M.K.W. by Versus Arthritis. In Iceland support from the European Commission to the painFACT project T.E.T. (H2020-2020-848099) is acknowledged. In Estonia, the work of E.A. was funded by the European Union through Horizon Europe research and innovation programs (grants 894987, 101137201, 101137154) and by Estonian Research Council Grant PRG1291.

## Competing interests

CMB is a consultant for Vertex Pharmaceuticals and Merck Pharmaceuticals providing expert medicolegal testimony. SF is a consultant for Vertex Pharmaceuticals. BG has received research grants (paid to institution) from Sandoz, AbbVie, AlfaSigma, and Eli Lilly. SB has ownership interests in Hoba Therapeutics Aps, Novo Nordisk A/S, Lundbeck A/S, and Eli Lilly and Co. CE has received unrestricted research grants from Novo Nordisk and Abbott Diagnostics (administered by Aarhus University Hospital, no personal fees received). GB, TET, GET, TAO, SS, HS, IJ, ATS, GT, LS, UT, PS, and DFG are employees of Amgen deCODE Genetics.

## Author Contributions

IK performed data curation, formal analysis, funding acquisition, investigation, methodology, project administration, visualization, and original draft preparation. GB contributed to phenotype definition, data acquisition, writing, critical review, and final approval. KA contributed to formal analysis and original draft preparation. LU contributed to data analysis. HH contributed to data analysis. GT and LS, contributed to data acquisition, writing, critical review, and final approval. SF contributed to data acquisition, critical review, and final approval. JV contributed to data analysis. LK contributed to data acquisition, critical review, and final approval. EA contributed to data curation, formal analysis, funding acquisition, investigation, methodology, and project administration. CJ contributed to formal analysis and original draft preparation. BA, HB, SB, MTB, MD, CE, AJG, DFG, TFH, IJ, SK, KUK, CM, LDN, TAO, SRO, OBVP, SS, ATS, ES, HS, PS, OAS, GET, UT, HU, AV and TMW contributed to data acquisition, phenotype definitions, critical review, and final approval. RS contributed to conceptualization and supervision. KS provided overall supervision. CMB contributed to data acquisition, critical review, and final approval. BG contributed to critical review and final approval. DJC contributed to conceptualization, critical review, and final approval. TET contributed to data acquisition, writing, critical review, and final approval. FMKW contributed to conceptualization, writing, and final approval. NSA contributed to conceptualization, data curation, formal analysis, funding acquisition, investigation, methodology, project administration, supervision, and both original draft writing and review and editing. HMO contributed to conceptualization, data curation, formal analysis, funding acquisition, investigation, methodology, project administration, supervision, and both original draft writing and review and editing. MW contributed to conceptualization, data curation, formal analysis, funding acquisition, investigation, methodology, project administration, supervision, and both original draft writing and review and editing.

## Methods

### Phenotype definition

Cases were individuals with ≥1 inpatient or primary-care record containing ICD-10 code M79.7 (fibromyalgia). No exclusion criteria were applied: controls were all genotyped participants without code M79.7. Phenotype ascertainment was performed independently within each cohort prior to genome-wide association analysis.

### Genotyping, imputation and quality control

We included autosomal and chromosome X data for all cohorts.

#### All of Us

We obtained whole-genome sequencing (WGS, GRCh38) data from All of Us’s Curated Data Repository (CDR) version 8 release, which involved variant calling, initial quality control, and genetic ancestry assignment. We used the ACAF threshold callset, which contains variants with minor allele count > 100 or minor allele frequency > 1% in any genetic ancestry group, as the basis for a second round of ancestry-specific quality control with version 2.0.0 of the plink genetic analysis toolkit^125^, in which variants with minor allele frequency (MAF) < 0.1%, missingness > 10%, or Hardy-Weinberg equilibrium (HWE) p < 1 × 10^-15^ (with mid-P correction) were removed. Participants flagged by All of Us in the first round of quality control, those with sex-chromosome aneuploidy, or sex at birth not equal to male or female were excluded. We performed association analysis on each of the 3 largest genetic ancestry groups – European (EUR), African (AFR), and Admixed American (AMR) – with version 3.3 of the regenie GWAS toolkit^126^, using as covariates age, sex, age^2^, age × sex, age^2^ × sex, and the first 10 genotype principal components. For step 1 of regenie, we used an LD-pruned subset of the full genotypes, calculated with plink version 2.0.0 via the option “--indep-pairwise 500kb 1 0.2”.

We gratefully acknowledge All of Us participants for their contributions, without whom this research would not have been possible. We also thank the National Institutes of Health’s All of Us Research Program for making available the participant data examined in this study.

#### UK Biobank

We obtained imputed genotypes from the UK Biobank’s <u>Data-Field 22828</u>. The UK Biobank imputed genotypes using a combination of two GRCh37 reference panels, the Haplotype Reference Consortium^127^ and a combined UK10K^128^ and 1000 Genomes Phase 3^129^ panel, using the Haplotype Reference Consortium imputation if a variant was present in both panels^39^. We excluded samples with sex-chromosome aneuploidy (<u>Data-Field 22019</u>), that were outliers for heterozygosity or missingness rate (<u>Data-Field 22027</u>), or that had discordant genetic sex (<u>Data-Field 22001</u>) versus self-reported sex (<u>Data-Field 31</u>). We excluded variants with minor allele frequency (MAF) < 0.1%, imputation INFO score < 0.8, missingness > 10 %, or HWE p < 1 × 10^-15^ (with mid-P correction) with plink version 2.0.0. We performed associations on the 3 largest genetic ancestry groups as defined by <u>Pan-UKBB</u> (<u>Return 2442</u>) – European (EUR), Central/South Asian (CSA), and African (AFR) – with version 3.4.1 of regenie, using as covariates age, sex, age^2^, age × sex, age^2^ × sex, and the first 10 genotype principal components. For step 1 of regenie, we used an LD-pruned subset of the full genotypes, calculated with the “--indep-pairwise 500kb 1 0.2” option from plink version 2.0.0.

We thank all participants of the UK Biobank. This research has been conducted using the UK Biobank Resource under application number 116200.

#### FinnGen

Genotyping in the FinnGen cohort was performed by using Illumina (Illumina Inc., San Diego, CA, USA) and Affymetrix arrays (Thermo Fisher Scientific, Santa Clara, CA, USA) and lifted over to Genome Reference Consortium Human Build version 38 (GRCh38/hg38). Individuals with high genotype absence (>5%), inexplicit sex or excess heterozygosity (±4 standard deviations) were excluded from the data. Additionally, variants that had high absence (>2%), low minor allele count (<3) or low Hardy-Weinberg Equilibrium (HWE) (P < 1 × 10^-6^) were removed. More detailed explanations of the genotyping, quality control and the genotype imputation are provided elsewhere^130^. All individuals in the cohort were Finns and matched against the SiSu v4 reference panel .

For the FinnGen cohort (Data Freeze 12), GWAS was conducted using the REGENIE pipeline (https://github.com/FINNGEN/regenie-pipelines). Analysis was adjusted for age at death or end of follow up, sex, genotyping batches and the first 10 genetic principal components. Firth approximation was applied for variants with association p-value <0.01.

Study subjects in FinnGen provided informed consent for biobank research, based on the Finnish Biobank Act. Alternatively, separate research cohorts, collected prior the Finnish Biobank Act came into effect (in September 2013) and start of FinnGen (August 2017), were collected based on study-specific consents and later transferred to the Finnish biobanks after approval by Fimea (Finnish Medicines Agency), the National Supervisory Authority for Welfare and Health. Recruitment protocols followed the biobank protocols approved by Fimea. The Coordinating Ethics Committee of the Hospital District of Helsinki and Uusimaa (HUS) statement number for the FinnGen study is Nr HUS/990/2017.

The FinnGen study is approved by Finnish Institute for Health and Welfare (permit numbers: THL/2031/6.02.00/2017, THL/1101/5.05.00/2017, THL/341/6.02.00/2018, THL/2222/6.02.00/2018, THL/283/6.02.00/2019, THL/1721/5.05.00/2019 and THL/1524/5.05.00/2020), Digital and population data service agency (permit numbers: VRK43431/2017-3, VRK/6909/2018-3, VRK/4415/2019-3), the Social Insurance Institution (permit numbers: KELA 58/522/2017, KELA 131/522/2018, KELA 70/522/2019, KELA 98/522/2019, KELA 134/522/2019, KELA 138/522/2019, KELA 2/522/2020, KELA 16/522/2020), Findata permit numbers THL/2364/14.02/2020, THL/4055/14.06.00/2020, THL/3433/14.06.00/2020, THL/4432/14.06/2020, THL/5189/14.06/2020, THL/5894/14.06.00/2020, THL/6619/14.06.00/2020, THL/209/14.06.00/2021, THL/688/14.06.00/2021, THL/1284/14.06.00/2021, THL/1965/14.06.00/2021, THL/5546/14.02.00/2020, THL/2658/14.06.00/2021, THL/4235/14.06.00/2021, Statistics Finland (permit numbers: TK-53-1041-17 and TK/143/07.03.00/2020 (earlier TK-53-90-20) TK/1735/07.03.00/2021, TK/3112/07.03.00/2021) and Finnish Registry for Kidney Diseases permission/extract from the meeting minutes on 4^th^ July 2019.

The Biobank Access Decisions for FinnGen samples and data utilized in FinnGen Data Freeze 12 include: THL Biobank BB2017_55, BB2017_111, BB2018_19, BB_2018_34, BB_2018_67, BB2018_71, BB2019_7, BB2019_8, BB2019_26, BB2020_1, BB2021_65, Finnish Red Cross Blood Service Biobank 7.12.2017, Helsinki Biobank HUS/359/2017, HUS/248/2020, HUS/430/2021 §28, §29, HUS/150/2022 §12, §13, §14, §15, §16, §17, §18, §23, §58, §59, HUS/128/2023 §18, Auria Biobank AB17-5154 and amendment #1 (August 17 2020) and amendments BB_2021-0140, BB_2021-0156 (August 26 2021, Feb 2 2022), BB_2021-0169, BB_2021-0179, BB_2021-0161, AB20-5926 and amendment #1 (April 23 2020) and it’s modifications (Sep 22 2021), BB_2022-0262, BB_2022-0256, Biobank Borealis of Northern Finland_2017_1013, 2021_5010, 2021_5010 Amendment, 2021_5018, 2021_5018 Amendment, 2021_5015, 2021_5015 Amendment, 2021_5015 Amendment_2, 2021_5023, 2021_5023 Amendment, 2021_5023 Amendment_2, 2021_5017, 2021_5017 Amendment, 2022_6001, 2022_6001 Amendment, 2022_6006 Amendment, 2022_6006 Amendment, 2022_6006 Amendment_2, BB22-0067, 2022_0262, 2022_0262 Amendment, Biobank of Eastern Finland 1186/2018 and amendment 22§/2020, 53§/2021, 13§/2022, 14§/2022, 15§/2022, 27§/2022, 28§/2022, 29§/2022, 33§/2022, 35§/2022, 36§/2022, 37§/2022, 39§/2022, 7§/2023, 32§/2023, 33§/2023, 34§/2023, 35§/2023, 36§/2023, 37§/2023, 38§/2023, 39§/2023, 40§/2023, 41§/2023, Finnish Clinical Biobank Tampere MH0004 and amendments (21.02.2020 & 06.10.2020), BB2021-0140 8§/2021, 9§/2021, §9/2022, §10/2022, §12/2022, 13§/2022, §20/2022, §21/2022, §22/2022, §23/2022, 28§/2022, 29§/2022, 30§/2022, 31§/2022, 32§/2022, 38§/2022, 40§/2022, 42§/2022, 1§/2023, Central Finland Biobank 1-2017, BB_2021-0161, BB_2021-0169, BB_2021-0179, BB_2021-0170, BB_2022-0256, BB_2022-0262, BB22-0067, Decision allowing to continue data processing until 31^st^ Aug 2024 for projects: BB_2021-0179, BB22-0067,BB_2022-0262, BB_2021-0170, BB_2021-0164, BB_2021-0161, and BB_2021-0169, and Terveystalo Biobank STB 2018001 and amendment 25^th^ Aug 2020, Finnish Hematological Registry and Clinical Biobank decision 18^th^ June 2021, Arctic biobank P0844: ARC_2021_1001.

#### Estonian Biobank

All the Estonian Biobank participants have been genotyped at the Core Genotyping Lab of the Institute of Genomics, University of Tartu, using Illumina Global Screening Array v3.0_EST. Samples were genotyped and PLINK format files were created using Illumina GenomeStudio v2.0.4. Individuals were excluded from the analysis if their call-rate was <95%, if they were outliers of the absolute value of heterozygosity (>3SD from the mean) or if sex defined based on heterozygosity of X chromosome did not match sex in phenotype data. Before imputation, variants were filtered by call-rate <95%, HWE p-value < 1 × 10−4 (autosomal variants only), and minor allele frequency <1%. Genotyped variant positions were in build 37 and were lifted over to build 38 using Picard. Phasing was performed using the Beagle v5.4 software. Imputation was performed with Beagle v5.4 software (beagle.22Jul22.46e.jar) and default settings. The dataset was split into batches of 5,000. A population-specific reference panel consisting of 2,695 WGS samples was utilized for imputation and standard Beagle hg38 recombination maps were used. Based on the principal component analysis, samples who were not of European ancestry were removed. Duplicate and monozygous twin detection was performed with KING 2.2.7, and one sample was removed from the pair of duplicates. Analyses were restricted to individuals with European ancestry.

Association analysis in the Estonian Biobank was carried out for all variants with an INFO score >0.4 using the additive model as implemented in REGENIE v3.0.3 with standard binary trait settings. Logistic regression was carried out with adjustment for current age, age2, sex and 10 first genetic principal components as covariates, analyzing only variants with a minimum minor allele count of 2.

The activities of the Estonian Biobank are regulated by the Human Genes Research Act, which was adopted in 2000 specifically for the operations of the Estonian Biobank. Individual level data analysis in the Estonian Biobank was carried out under ethical approval 1.1-12/624 from the Estonian Committee on Bioethics and Human Research (Estonian Ministry of Social Affairs), using data according to release application 6-7/GI/33543 from the Estonian Biobank.

We thank participants of the Estonian Biobank for their contributions. Estonian Biobank analyses were partially carried out at the High Performance Computing Center, University of Tartu, under ethical approval 1.1-12/624 from the Estonian Committee on Bioethics and Human Research (Estonian Ministry of Social Affairs), using data according to release application 6-7/GI/2015 from the Estonian Biobank. The Estonian Biobank Research Team was responsible for data collection, genotyping, quality control, and imputation, and consisted of Andres Metspalu, Mait Metspalu, Lili Milani, Reedik Mägi, Mari Nelis, Georgi Hudjashov, and Tõnu Esko.

#### Genes and Health

Genotyping was performed on Illumina Infinium Global Screening Array v3 with additional multi-disease variants. Quality control was performed following a standardized approach^131^. In brief, variants with call rates less than 0.99 and/or minor allele frequency (MAF)L<L1% were excluded. We excluded individuals unlikely to have genetically inferred Pakistani or Bangladeshi ancestry. Imputation was performed using the TOPMed-r2 panel. We excluded SNPs with low imputation scores (INFOL<L0.3) or MAFL<L0.1%. In the Genes and Health cohort, sex was defined on the basis of XX (female) and XY (male) chromosomal presence in genotype data.

Diagnoses were curated from routine UK NHS EHR data from primary care (Systematized Nomenclature of Medicine (SNOMED) coded) and secondary care (International Classification of Diseases, Tenth Revision (ICD-10) coded) sources. Data were combined without mapping between coding formats. For each clinical code, the earliest ever measure recorded in a participant’s medical records was used, excluding erroneous code dates preceding the participant’s recorded date of birth.

Genes & Health is/has recently been core-funded by Wellcome (WT102627, WT210561), the Medical Research Council (UK) (M009017, MR/X009777/1, MR/X009920/1), Higher Education Funding Council for England Catalyst, Barts Charity (845/1796), Health Data Research UK (for London substantive site), and research delivery support from the NHS National Institute for Health Research Clinical Research Network (North Thames). We acknowledge the support of the National Institute for Health and Care Research Barts Biomedical Research Centre (NIHR203330); a delivery partnership of Barts Health NHS Trust, Queen Mary University of London, St George’s University Hospitals NHS Foundation Trust and St George’s University of London.

Genes & Health is/has recently been funded by Alnylam Pharmaceuticals, Genomics PLC; and a Life Sciences Industry Consortium of AstraZeneca PLC, Bristol-Myers Squibb Company, GlaxoSmithKline Research and Development Limited, Maze Therapeutics Inc, Merck Sharp & Dohme LLC, Novo Nordisk A/S, Pfizer Inc, Takeda Development Centre Americas Inc.

We thank Social Action for Health, Centre of The Cell, members of our Community Advisory Group, and staff who have recruited and collected data from volunteers. We thank the NIHR National Biosample Centre (UK Biocentre), the Social Genetic & Developmental Psychiatry Centre (King’s College London), Wellcome Sanger Institute, and Broad Institute for sample processing, genotyping, sequencing and variant annotation. This work uses data provided by patients and collected by the NHS as part of their care and support. This research utilised Queen Mary University of London’s Apocrita HPC facility, supported by QMUL Research-IT, http://doi.org/10.5281/zenodo.438045.

We thank: Barts Health NHS Trust, NHS Clinical Commissioning Groups (City and Hackney, Waltham Forest, Tower Hamlets, Newham, Redbridge, Havering, Barking and Dagenham), East London NHS Foundation Trust, Bradford Teaching Hospitals NHS Foundation Trust, Public Health England (especially David Wyllie), Discovery Data Service/Endeavour Health Charitable Trust (especially David Stables), Voror Health Technologies Ltd (especially Sophie Don), NHS England (for what was NHS Digital) - for GDPR-compliant data sharing backed by individual written informed consent.

Most of all we thank all of the volunteers participating in Genes & Health.

A favourable ethical opinion for the main Genes & Health research study was granted by NRES Committee London - South East (reference 14/LO/1240) on 16 Sept 2014. Queen Mary University of London is the Sponsor, and Data Controller.

#### Mass General Brigham Biobank

The MGB Biobank genotyped 53,297 participants on the Illumina Global Screening Array (’GSA’) and 11,864 on Illumina Multi-Ethnic Global Array (“MEG”). The GSA arrays captured approximately 652K SNPs and short indels, while the MEG arrays captured approximately 1.38M SNPs and short indels. These genotypes were filtered for high missingness (>2%) and variants out of HWE (p < 1e-12), as well as variants with an AF discordant (p < 1e-150) from a synthesized AF calculated from GnomAD subpopulation frequencies and a genomewide GnomAD model fit of the entire cohort. This resulted in approximately 620K variants for GSA and 1.15M for MEG. The two sets of genotypes were then separately phased and imputed on the TOPMed imputation server (Minimac4 algorithm) using the TOPMed r2 reference panel. The resultant imputation sets were both filtered at an R^2^ > 0.4 and a MAF > .001, and then the two sets were merged/intersected resulting in approximately 19.5M GRCh38 autosomal variants. The sample set for analysis was then restricted to just those classified as EUR according to a random-forest classifier trained with the Human Genome Diversity Project (HGDP)^132^ as the reference panel, with the minimum probability for assignment to an ancestral group of 0.5, in 19/20 iterations of the model. To correct for population stratification, PCs were computed in genetically European participants. Association analysis for the full-cohort (N = 51053) was performed with variants using REGENIE (v3.2.8)^126^ with adjustment for age, sex, genotype-chip, tranche, and PC 1-10. The summary statistics were filtered with a minimum minor allele count of 50. These summary statistics were then lifted over to GRCh37.

In the MGB Biobank, the sex-specific (N of females = 27,682 & N of males = 23,371) association analyses were performed in the GRCh38 build and carried out using REGENIE v3.2.8 with covariates of age, tranche, genotype-chip and the first ten principal components of ancestry calculated by performing the within-EUR PCA analysis. The summary statistics were filtered with a minimum minor allele count of 25.

#### Michigan Genomics Initiative

We included autosomal and chromosome X data from the Michigan Genomics Initiative (MGI), a health system-based biobank of patients recruited primarily during surgical encounters at Michigan Medicine. As of Freeze 6, MGI comprised 80,529 participants with linked genotype and electronic health record data. Detailed descriptions of the MGI cohort, recruitment protocols, and overall design are available in Zawistowski et al.^133^.

DNA samples were genotyped at the University of Michigan Advanced Genomics Core on customized versions of the Illumina Infinium CoreExome-24 (v1.0, v1.1, v1.3; ∼570K markers) or Illumina Infinium Global Screening Array (GSA v1.3; ∼682K markers). Array content included standard backbones with additional custom content to capture GWAS candidate variants, predicted loss-of-function alleles, ancestry-informative markers, and pharmacogenomic variants. Genotype calling was performed in GenomeStudio, supplemented by zCall for recovery of rare variants.

Sample-level QC excluded individuals for: consent withdrawal, genotype-inferred sex mismatch, sex chromosome aneuploidy, call rate <99%, contamination >2.5%, unresolved technical duplicates, or batch-level DNA extraction issues. Relatedness was estimated with KING v2.1.3, and contamination with VICES. Variant-level QC removed probes that did not uniquely map to GRCh38, sites with call rate <98%, Hardy– Weinberg equilibrium (HWE) p < 1 × 10^-4^ in unrelated European ancestry samples, or poor clustering metrics (GenTrain <0.15, Cluster Separation <0.3). Additional harmonization excluded variants with large allele frequency deviations compared to 1000 Genomes reference populations.

Phasing was performed with Eagle v2.4 using the TOPMed reference panel, followed by imputation against TOPMed haplotypes via the Michigan Imputation Server. Post-imputation, variants with Rsq <0.3 or MAF <0.01% were removed, yielding ∼52 million high-quality variants. Principal components (PCs) were calculated using FlashPCA2 after pruning rare and correlated variants.

For GWAS, we analyzed participants of genetically inferred European ancestry as defined by PCA projection and ADMIXTURE (K=7 reference populations from the Human Genome Diversity Project). Association testing was performed with SAIGE v0.35 (https://github.com/weizhouUMICH/SAIGE), a generalized mixed-model framework that accounts for relatedness and case-control imbalance. Covariates included age, sex, genotyping array, and the first ten genetic PCs. Variants with MAF ≥0.01% and imputation Rsq ≥0.3 were included in association testing.

The authors acknowledge the Michigan Genomics Initiative participants, AI & Digital Health Innovation at the University of Michigan, the University of Michigan Medical School Central Biorepository, and the University of Michigan Advanced Genomics Core for providing data and specimen storage, management, processing, and distribution services, and the Center for Statistical Genetics in the Department of Biostatistics at the School of Public Health for genotype data curation, imputation, and management in support of the research reported in this publication.

#### deCODE Genetics/Amgen Iceland

At the time of analysis, 63,118 samples from Icelandic participants were whole genome sequenced at deCODE using Illumina standard TruSeq methods to a mean depth of 38×^134,135^. Only samples with a genome-wide average coverage of 20× and higher were included. Genotypes of single nucleotide polymorphisms (SNPs) and insertions/deletions (indels) were identified and called jointly by Graphtyper(v.2.7.1)^136^. In all, 173,025 samples from Icelandic participants had been chip-genotyped using various Illumina SNP arrays^134,135^. The chip-typed individuals were long-range phased^137^, and the variants identified in the whole-genome sequences of Icelanders imputed into the chip-typed individuals. Using extensive and encrypted Icelandic genealogy data, familial imputation of genotypes in first- and second-degree relatives was used to increase sample size^134,137^. The final dataset used included 19,657,761 variants with imputation information over 0.8 and minor allele frequency (MAF) over 0.1%. The Icelandic samples and diagnostic data were obtained from Icelandic medical record data repositories, were analyzed under approval from the National Bioethics Committee (NBC #17-035-V11-S2, previously 12-162) following review by the Icelandic Data Protection Authority. Data was anonymized and encrypted by a third-party system, approved and monitored by the Icelandic Data Protection Authority^138^.

#### Copenhagen Hospital Biobank and the Danish Blood Donor Study

We obtained genotypes from the Copenhagen Hospital Biobank (CHB)^139^ study on pain and degenerative musculoskeletal diseases (CHB-PDS, approval: NVK-1803812, P-2019-51) and the Danish Blood Donor Study (DBDS, approval: NVK-1700407, P-2019-99)^140^. All samples were genotyped on Illumina’s Infinium Global Screening Array (versions 1.0 and 3.0) and imputed to build hg38. Genotyping and imputation were performed by deCODE genetics. Prior to imputation, duplicate samples and those with genotype call rate < 98% were removed. Phasing was carried out with SHAPEIT4^141^, and imputation was performed using deCODE’s in-house workflow, with a joint Graphtyper-based reference panel comprising ∼50,000 individuals, including ∼10,800 Danes, including danmac5.dk^142^. Initial quality control of genotypes included removal of samples with sex discrepancies, missingness > 5%, and variants with missingness > 10% or Hardy-Weinberg equilibrium p < 1 × 10LL. For genome-wide association analyses, we removed variants with MAF < 0.1%, imputation INFO < 0.8, missingness > 10%, or HWE p < 1 × 10L¹L (with mid-P correction), using plink version 2.0.0. Association analyses were performed separately in DBDS and CHB-PDS with Regenie v3.4.1, including as covariates age, sex, age², age × sex, age² × sex, and the first 10 principal components.

#### Intermountain Health

Under research collaboration between Intermountain Health, Utah, USA and deCODE Genetics/Amgen, samples were obtained from consenting participants of European descent in two ongoing studies; The Intermountain Inspire Registry and The HerediGene Population study^143^. The Intermountain Healthcare Institutional Review Board approved both studies, and all participants provided written informed consent prior to enrollment. Samples were genotyped at deCODE Genetics/Amgen using Illumina Global Screening Array chips. In all, 138,006 individuals of European origin were chip-typed. Intermountain and Danish imputation was based on a multi-ethnic reference panel of 50,179 whole genome sequenced individuals of mostly European descent, including 23,288 individuals from Intermountain and 11,722 individuals from Denmark.

Samples were filtered on 98% variant yield and duplicates removed. The WGS protocol was the same as described above for the Icelandic and Danish data. Sequence variants were imputed into 138,006 chip-typed individuals. Over 245 million high-quality sequence variants and indels, sequenced to a mean depth of 20×, were identified using Graphtyper (v.2.7.1)^136^. Quality-controlled chip genotype data were phased using Shapeit 4^141^. A phased haplotype reference panel was prepared from the sequence variants using the long-range phased chip-genotyped samples^134,135^. In all, 21,316,504 variants (imputation info > 0.8 and MAF > 0.1%) were tested.

#### Nashville Biosciences

The Nashville Biosciences (NashBio) is a data and analytics provider owned by The Vanderbilt University Medical Center (VUMC, Tennessee, USA). NashBio utilizes VUMC’s biobank collection BioVU® that includes a collection of de-identified DNA samples linked to de-identified data from the VUMĆs electronic health records referred to as Synthetic Derivative (SD) database. All patients of the VUMC consented to their residual samples from routine clinical testing and data being contributed to BioVU®. BioVU® extracts and banks germline DNA samples that are de-identified and only linked to the SD through a randomly assigned unique identifier, not back to the patient or their underlying medical record. The use of BioVU® is classified as non-human subject research by VUMC’s Institutional Review Board (IRB), and NashBio is not required to seek study-specific consent for use of these datasets. The overall biobanking program is reviewed annually by the IRB to maintain this determination and make decisions about patient protections, privacy, and ethical issues. Each individual study seeking to use the SD database and BioVU® biobank is filed with VUMC’s IRB to validate its non-human subject classification and 441 appropriate use of data.

TheNashBio dataset included in this study is a subset of BioVU®, consisting of in total 80,965 whole genome sequenced individuals of genetically defined European descent and 31,025 individuals of genetically defined African descent. All samples were whole genome sequenced at deCODE Genetics/Amgen using Illumina NovaSeq in accordance with the above described deCODE/Amgen protocols. Using the same quality criteria as for other deCODE/Amgen analyzed datasets, in total 22,316,589 variants were tested in the European cohort and 33,108,534 in the African cohort.

#### Genetic ancestry analysis in deCODE datasets

For the non-Icelandic datasets analyzed at deCODE Genetics/Amgen (Copenhagen Hospital Biobank and the Danish Blood Donor Study, Intermountain Health and Nashville Biosciences), genetic ancestry analysis was performed to identify a subset of individuals with similar ancestry. For the Danish samples we used ADMIXTURE (v1.23)^144^ run in supervised mode using the 1000 Genomes populations^129^, CEU (Utah residents with Northern and Western European ancestry), CHB (Han Chinese in Beijing, China), ITU (Indian Telugu in the UK), PEL (Peruvian in Lima, Peru) and YRI (Yoruba in Ibadan, Nigeria), as training samples. These training samples had themselves been filtered for ancestry outliers using principal component analysis (PCA) and unsupervised ADMIXTURE. Samples assigned <0.93 CEU were excluded resulting in 358,483 samples included in the analysis. For the Intermountain samples we used the same methods as for the Danish samples to identify a subset of 108,149 individuals of European descent that were included in the analysis.

To identify ancestry-homogeneous subsets in the Nashville Biosciences cohort, sample ancestry was estimated with ADMIXTURE (v1.23)^144^ in supervised mode using the 1000 Genomes populations^129^, CEU (Utah residents with Northern and Western European ancestry), CHB (Han Chinese in Beijing, China), ITU (Indian Telugu in the UK), PEL (Peruvian in Lima, Peru) and YRI (Yoruba in Ibadan, Nigeria) as training samples. Samples assigned >0.93 CEU ancestry were placed in the EUR cohort. Samples with admixed African and European ancestry (defined as YRI + CEU > 0.9, YRI > 0.3, and CEU > 0.02) or predominantly African ancestry (YRI > 0.9) were placed in the AFR subcohort. Genotypes were prepared for principal components analysis (PCA) with PLINK v1.9^125^ using --maf 0.01 --thin-count 1000000 --indep-pairwise 60000 6000 0.3 to minimize effects of local linkage disequilibrium and very recent population structure. Pairwise relatedness was calculated using KING v2.3.0 --ibdseg --degree 3^145^ to identify relatives of third degree or closer, with one sample from each pair of relatives removed prior to PCA and projected onto the resulting PCs using a deCODE in-house script which adjusts for shrinkage. PCA was performed with PCAone v0.3.4^146^ using default parameters.

#### Association testing in deCODE datasets

The four fibromyalgia case/control GWASs performed at deCODE Genetics/Amgen (Iceland, Denmark, US Intermountain Health and US NashBio) all used the ICD-10 diagnosis M79.7 to define cases and no exclusions were applied to controls. Genome-wide associations were performed using software developed at deCODE Genetics, using logistic regression assuming an additive model^134^. For the Icelandic data, the model included sex, county of birth, current age or age at death (first-and second-order terms included), blood sample availability for the individual, sequencing status, and an indicator function for the overlap of the lifetime of the individual with the time span of phenotype collection. To include imputed but ungenotyped individuals in Iceland, we used county of birth as a proxy covariate for the first principal component (PC) because county of birth has been shown to be in concordance with the first PC in Iceland^147^. For the Danish data, 12 PCs were used, in addition to sex, year of birth and sequencing status as covariates, whereas for the Intermountain Health data, we included 4 PCs, year of birth, sex and sequencing status as covariates. For the NashBio data, associations were adjusted for the top 20 PCs in addition to sex, year of birth and sequencing batch. The number of PCs used to adjust for population stratification was determined by identifying the point at which further PCs appeared to capture local LD rather than population structure as reflected by sharp peaks in PC loadings and in plateauing of eigenvalues^148^. Software developed at deCODE Genetics was used for these analyses^134^. All statistical tests were two-sided unless otherwise indicated.

### Harmonization and meta-analysis

Each cohort’s summary statistics were harmonized via an in-house analysis pipeline. For each summary statistics file (i.e. for each cohort and ancestry), we removed variants with missing data in any column, non-ACGT alleles, p-values outside (0, 1], allele frequencies outside (0, 1), imputation INFO scores outside (0, 1], or non-positive odds ratios, standard errors, or sample sizes. We normalized indels to their minimal representation by iteratively removing common trailing then leading base pairs from the reference and alternate alleles – subject to the constraint that each allele retains at least one base pair – and advancing the genomic position by the number of leading base pairs removed. We inferred rs numbers based on chromosome and base-pair columns using dbSNP build 156, requiring an exact match to both the ref and the alt allele, allowing allele flips for single-nucleotide variants (but not indels), and converting the dbSNP variants to minimal representation as well before doing the matching. We removed variants without rs numbers in dbSNP, or that mapped to multiple genomic positions (this happens very rarely). To account for dbSNP’s tendency to merge rs numbers over time, if a variant mapped to multiple rs numbers, we took the lowest-numbered rs number. We calculated each cohort and ancestry’s effective sample size per-variant via the formula N_eff_ = ((4 / (2 × AAF × (1 -AAF) × INFO)) -BETA^2^) / SE^2^, skipping the “× INFO” if INFO was not available, and summed these N_eff_ across cohorts and ancestries.

To account for p-value inflation due to residual confounding like cryptic relatedness, we applied stratified LD score regression^149,150^ to each cohort and ancestry’s summary statistics prior to meta-analysis, using the effective sample size N_eff_ rather than the raw N to avoid bias^151^. We used the standard 52-annotation baseline model precomputed by the developers of stratified LD score regression. We manually calculated LD scores based on ancestry-specific reference panels from the corresponding 1000 Genomes Phase 3 superpopulation (EUR for Europeans, SAS for Central/South Asians, AFR for Africans, AMR for Admixed Americans). However, we subset to variants present in Europeans in HapMap 3^152^, since the standard 52 annotations are only available for those variants. For cohort-ancestry pairs where the LD score regression intercept was greater than 1, we divided each variant’s X^2^ statistic by the LD score regression intercept, then adjusted the standard errors and p-values accordingly. This has the effect of reducing the significance of every variant in summary statistics with evidence of inflation, while leaving non-inflated summary statistics alone.

After harmonization and LD score regression correction, we performed standard fixed-effects inverse-variance weighted meta-analyses via the “--meta-analysis” option from plink version 2.0.0. Variants were matched across cohorts based on the combination of rs number, reference allele, and alternate allele. In total, 54,629 cases and 2,509,126 controls went into the multi-ancestry meta-analysis, and 49,000 cases and 2,264,287 controls went into the European genetic ancestry-only analysis. We also performed sex-stratified meta-analyses restricted to males and females, and leave-one-cohort-out analyses for each of the 11 constituent cohorts, with both multi-ancestry (leaving out all ancestries from that cohort, if the cohort had multiple) and European-only versions. For females we performed both European-only and multi-ancestry meta-analyses, but for males we performed only the European-only meta-analysis due to the lack of male summary statistics in Genes and Health and the low number of male cases in non-European ancestries in the multi-ancestry cohorts.

After meta-analysis, we filtered to variants with an overall minor allele frequency of > 1% across all cohorts that went into that particular meta-analysis. Since two cohorts (the UK Biobank and Mass General Brigham Biobank) reported variants in GRCh37 coordinates, we avoided having to perform an (error-prone) remapping of variant positions from GRCh37 to GRCh38 by filtering to variants present in at least 5 cohorts in the multi-ancestry meta-analyses and at least 3 studies in the European meta-analyses, thereby ensuring that every variant would be present in at least one GRCh38 study by the pigeonhole principle.

### Causal gene prioritization

We prioritized causal genes for each risk locus in the multi-ancestry meta-analysis via a combination of approaches. First, we performed manual literature search for each lead variant, focusing heavily on the nearest gene to each lead variant since nearest genes are expected to be causal about two-thirds of the time^153^. We supplemented this search with two orthogonal bioinformatic approaches: the state-of-the-art machine learning method FLAMES^47^ and an in-house quantitative trait locus (QTL)-based pipeline developed by deCODE genetics.

#### FLAMES

FLAMES^47^ is a framework that nominates candidate causal genes at risk loci from a GWAS, via an ensemble of two complementary approaches.

First, a gradient boosting model scores genes based on locus-specific variant-to-gene evidence, aggregating features like QTLs, Variant Effect Predictor (VEP) annotations, chromatin interactions, and variant-to-gene distance. The model’s predictive weights were established by training it on external GWAS loci, where “silver-standard” causal genes had been identified via missense or predicted loss-of-function variants from exome studies. For this analysis, we supplied the model with 99% credible sets derived from SuSiE (Sum of Single Effects^154^ fine-mapping of variants within 500 kilobases of the lead variant at each locus from our primary meta-analysis.

Second, FLAMES incorporates gene-level scores from Polygenic Priority Score (PoPS)^155^, another tool for causal gene prioritization. PoPS prioritizes genes by identifying gene features that are enriched for genetic association across the entire GWAS – such as pathway memberships, protein-protein interaction networks, co-expression data, and expression in various tissues and cell types – then scoring each gene based on which of these enriched features it has. PoPS identifies these trait-relevant gene features by leveraging MAGMA (Multi-marker Analysis of GenoMic Annotation)^156^ to aggregate variant-level GWAS p-values into gene-level association scores, then using a regression model to identify which gene features are predictive of high MAGMA scores.

FLAMES integrates these two evidence streams by multiplying the gradient boosting scores with scaled PoPS scores, effectively upweighting genes supported by both locus-specific and GWAS-wide evidence. At the recommended confidence threshold (scaled FLAMES score > 0.248), FLAMES nominated a candidate causal gene at 19 of the 26 loci from the primary meta-analysis, while abstaining from prediction at the remaining 7 loci due to a lack of confidence in the causal gene.

#### deCODE pipeline

As a second strategy to nominate candidate causal genes, we performed functional annotation and QTL analyses on all 26 lead variants from the primary meta-analysis, as well as all variants in high LD (r^2^ ≥ 0.8 and within ±1 megabase) with these lead variants.

We used Variant Effect Predictor (VEP)^157^ to attribute to the studied variants the most severe predicted variant consequences in canonical and non-canonical transcripts. We classified as high-impact variants those predicted as start-lost, stop gained, stop-lost, splice-donor, splice-acceptor, or frameshift, collectively called loss-of-function (LOF) variants, whereas moderate impact are variants predicted to otherwise affect coding or splicing of a protein (missense).

For all lead and correlated variants, we studied their association with a) mRNA expression (top local expression quantitative trait locus [eQTL], splicing QTL [sQTL], or alternative polyadenylation QTL [apaQTL]) in multiple tissues analyzed at deCODE, in addition to data from GTEx^158^ and other public datasets, b) plasma protein levels (top cis-protein QTL [pQTL]) identified in large proteomic datasets from Iceland and the UK^159^. RNA sequencing was performed on whole blood from 17,848 Icelanders and on subcutaneous adipose tissue from 769 Icelanders, respectively. Gene expression was computed based on personalized transcript abundances using kallisto^160^. Association between sequence variants and gene expression (cis-eQTL) was tested via linear regression, assuming additive genetic effect and normal quantile gene expression estimates, adjusting for measurements of sequencing artifacts, demographic variables, blood composition and PCs^161^. The gene expression PCs were computed per chromosome using a leave-one-chromosome-out method. All variants within 1 megabase of each gene were tested.

The Icelandic proteomics data were analyzed using the SomaLogic SOMAscan v4 proteomics assay that scans 4,907 aptamers, measuring 4,719 proteins in samples from 35,892 Icelanders with genetic information available at deCODE Genetics^159,162^. Plasma protein levels were standardized and adjusted for year of birth, sex and year of sample collection (2000-2019)^159,162^. The UK proteomics dataset was analyzed using the Olink Explore 3072 proximity extension assay (PEA) platform with 2,941 immunoassays characterizing 2,925 proteins in 54,265 participants in the UK Biobank^159,162^.

### Phenome-wide associations of lead variants

For each of our 26 lead variants from the primary meta-analysis, we looked up genome-wide significant (p < 5 × 10^-8^) associations in the GWAS Catalog that overlapped either the variant itself, or any proxy variants in high linkage disequilibrium (r^2^ ≥ 0.8 and within ±1 megabase). To account for the GWAS Catalog containing large numbers of closely related phenotypes, we grouped related traits for visualization. Specifically, we removed GWAS results obtained through MTAG analysis or performed on multiple traits combined, identified by “and” or “or” in their trait names. We then standardized trait names by removing words in parentheses and laterality descriptors (“left” and “right”), and standardized the spelling of “neutrophill” to “neurophil”. Finally, we grouped trait names that became identical after this standardization.

We also looked up the p-values of our 26 lead variants in pre-existing GWAS summary statistics for each of 330 disease endpoints from the November 2024 release of MVP-Finngen-UKBB (https://mvp-ukbb.finngen.fi/about), a GWAS meta-analysis of Million Veteran Program, FinnGen and the UK Biobank. We performed Bonferroni correction across the 330 diseases and 26 variants tested, leading to a significance threshold of p = 0.05 / 8580 ≈ 5.83 × 10^-6^.

We applied the same approach to 124 GWAS of drug prescription endpoints from FinnGen’s DF12 release, where each GWAS was on whether an individual was ever prescribed any drug from a particular category (e.g. analgesics). We performed Bonferroni correction across the 124 drug categories and 26 variants tested, leading to a significance threshold of p = 0.05 / 3224 ≈ 1.55 × 10^-5^.

### Heritability

For the European-only meta-analysis, we estimated single-nucleotide polymorphism-based heritability – the proportion of phenotypic variance explained by the aggregated effect of single-nucleotide polymorphisms across the autosomal genome – using stratified LD score regression^150^ with the standard 52-annotation baseline model mentioned above. We used LD scores computed from the European-ancestry subsample of 1000 Genomes Phase 3 (precomputed by LDSC authors), and subset to variants present in Europeans in HapMap 3. The observed-scale heritability was calculated from the LD score regression slope. As with all LD score regression-based analyses in this study, we used the effective sample size N_eff_ rather than the raw N to avoid bias due to case-control imbalance.

### Tissue and cell-type enrichment

We used LD score regression applied to specifically expressed genes (LDSC-SEG)^113^ to infer tissue and cell-type enrichments for the European-only meta-analysis. As for the global heritability analysis, we used LD scores computed from the European-ancestry subsample of 1000 Genomes Phase 3 (precomputed by LDSC authors) and the 52-annotation baseline model, subsetting to variants present in Europeans in HapMap 3. LDSC-SEG is a variant of stratified LD score regression that analyzes each tissue or cell type in turn, adding a binary annotation to the 52-annotation baseline model for each one. This annotation flags variants located within or within 100 kilobases of the 10% of genes most specifically expressed in that tissue or cell type. The heritability enrichment for each tissue or cell type is computed by dividing its per-SNP heritability (derived from the LD score regression slope) by the average per-SNP heritability across all SNPs in the analysis.

To compute tissue enrichments, we used annotations provided by the LDSC-SEG authors that were derived for each of 53 tissues in the Genotype-Tissue Expression (GTEx) project, where the top 10% of specifically expressed genes were defined by ranking genes by their differential expression *t*-statistic for expression in that tissue versus all other tissues. For GTEx brain tissues, the comparison was instead between that tissue and all non-brain tissues.

To compute cell-type enrichments, we generated gene sets from a comprehensive single-cell transcriptomic atlas of the mouse, PanSci, which profiles over 20 million cells from 14 organs and tissues^163^. From this dataset, we included 119 cell types and 7 broad lineages that were represented by at least 1,000 cells after quality control (filtering to cells with ≤10% mitochondrial reads, ≥100 genes detected, and non-zero Malat1 expression). Our analysis was restricted to a universe of 16,404 protein-coding genes with identifiable human orthologs. For each cell type and lineage, we defined its specifically expressed genes as those with a detection rate > 10% within that cell type and a fold-change > 2 compared to all other cells.

### Genetic correlation

We performed genetic correlation between our leave-FinnGen-out European-only fibromyalgia meta-analysis and each of 1,284 clinical endpoints from FinnGen Release 13 with (https://www.finngen.fi/en/researchers/clinical-endpoints), using LD score regression^149,164^. We calculated SNP heritability with LDSC for all 2,691 endpoints and performed genetic correlation only on those endpoints with significant heritability, resulting in a list of 1,284 endpoints. Variants were aligned and filtered to HapMap release 3 SNPs, and the LD reference was obtained from the European population of the 1000 Genomes Project Phase 3 release. We used the 80^th^ percentile of per-variant effective sample sizes (N_eff_) to munge the summary statistics.

To avoid diluting the results with low-specificity phenotypes, we then excluded endpoints containing any of the (case-insensitive) keywords “other” (e.g. “Other disorders of ear”), “unspecified” (e.g. “Disorder of external ear, unspecified”), classified (e.g. “Other viral diseases, not elsewhere classified”), “any” (e.g. “Any mental disorder”), or “all” (e.g. “All kidney diseases”), as well as derived endpoints for which the listed category did not correspond to a chapter of the International Classification of Diseases (ICD). We also excluded fibromyalgia itself as an endpoint. These exclusions reduced the number of endpoints tested from 1,284 to 855, resulting in a Bonferroni significance threshold of P = 0.05 / 855 ≈ 5.85 × 10^-5^.

For visualization purposes, we grouped endpoints by their ICD chapter. We grouped four categories with few significant endpoints – “III Diseases of the blood and blood-forming organs and certain disorders involving the immune mechanism”, “XVIII Symptoms, signs and abnormal clinical and laboratory findings, not elsewhere classified”, “XXI Factors influencing health status and contact with health services” and “XXII Codes for special purposes” – into an “Other” category. Because fibromyalgia has been previously proposed to share etiology with autoimmune disorders, we also created a separate “Autoimmune” category, since these disorders would otherwise be scattered across ICD chapters based on the affected organ system.

We also estimated the genetic correlation between fibromyalgia and the subtypes of asthma and rheumatoid arthritis using summary statistics available at deCODE. In line with other LDSC-based analyses, we used per-variant effective sample sizes, filtered variants to HapMap release 3 SNPs, and used precomputed LD scores for European populations (downloaded from https://data.broadinstitute.org/alkesgroup/LDSCORE/eur_w_ld_chr.tar.bz2).

### Polygenic risk scores

We computed polygenic risk scores (PRSs) for the leave-UK Biobank-out European and multi-ancestry meta-analyses, using PRS-CS^165^. PRS-CS takes GWAS summary statistics and an LD reference panel as input, and applies Bayesian shrinkage to the variants’ effect sizes to infer posterior effect sizes, which are used as the weights of the polygenic risk score. These weights can then be scored on any cohort of interest, which should be non-overlapping with the cohorts used to create the PRS to avoid bias.

We used the European-ancestry subsample of 1000 Genomes Phase 3 as the reference panel, removing indels from the summary statistics prior to the analysis. We ran PRS-CS with the “--n_burnin 5000” and “--n_iter 10000” options to specify 5,000 burn-in iterations and 10,000 total iterations of the Markov Chain Monte Carlo sampling. We then scored the PRS weights on the UK Biobank cohort with the “--score” option from plink version 2.0.0. Since PRS-CS requires a single sample size across all variants, the sample size we provided to PRS-CS for was the 80^th^ percentile of per-variant effective sample sizes (N_eff_), as recommended by others for PRS analyses^166^.

To assess the accuracy of the PRS across ancestries, we scored the PRS weights from the multi-ancestry leave-UK Biobank-out meta-analysis on each of the 3 largest genetic ancestries from the UK Biobank (European, Central/South Asian, and African). We assessed PRS accuracy via the area under the receiver-operating curve, as well as by computing odds ratios for each quintile of polygenic risk relative to the middle quintile, and prevalence of fibromyalgia within each quintile.

## Notes

### Author Declarations

Ethics committee/IRB of All of Us, Estonian Biobank, FinnGen, Genes and Health, deCODE Iceland, UK Biobank, Nashville Biosciences, Copenhagen Hospital Biobank and Danish Blood Donor Study, Intermountain Health, Michigan genomics Initiative, and Mass General Brigham Biobank gave ethical approval for this work. This research was conducted under the auspices of Research Ethics Board application #23-0111-C from Mount Sinai Hospital, "Uncovering the causal genetic variants, genes and cell types underlying brain disorders".

