## Supplementary materials for "The genetic architecture of fibromyalgia across 2.5 million individuals"

### Supplementary Figures

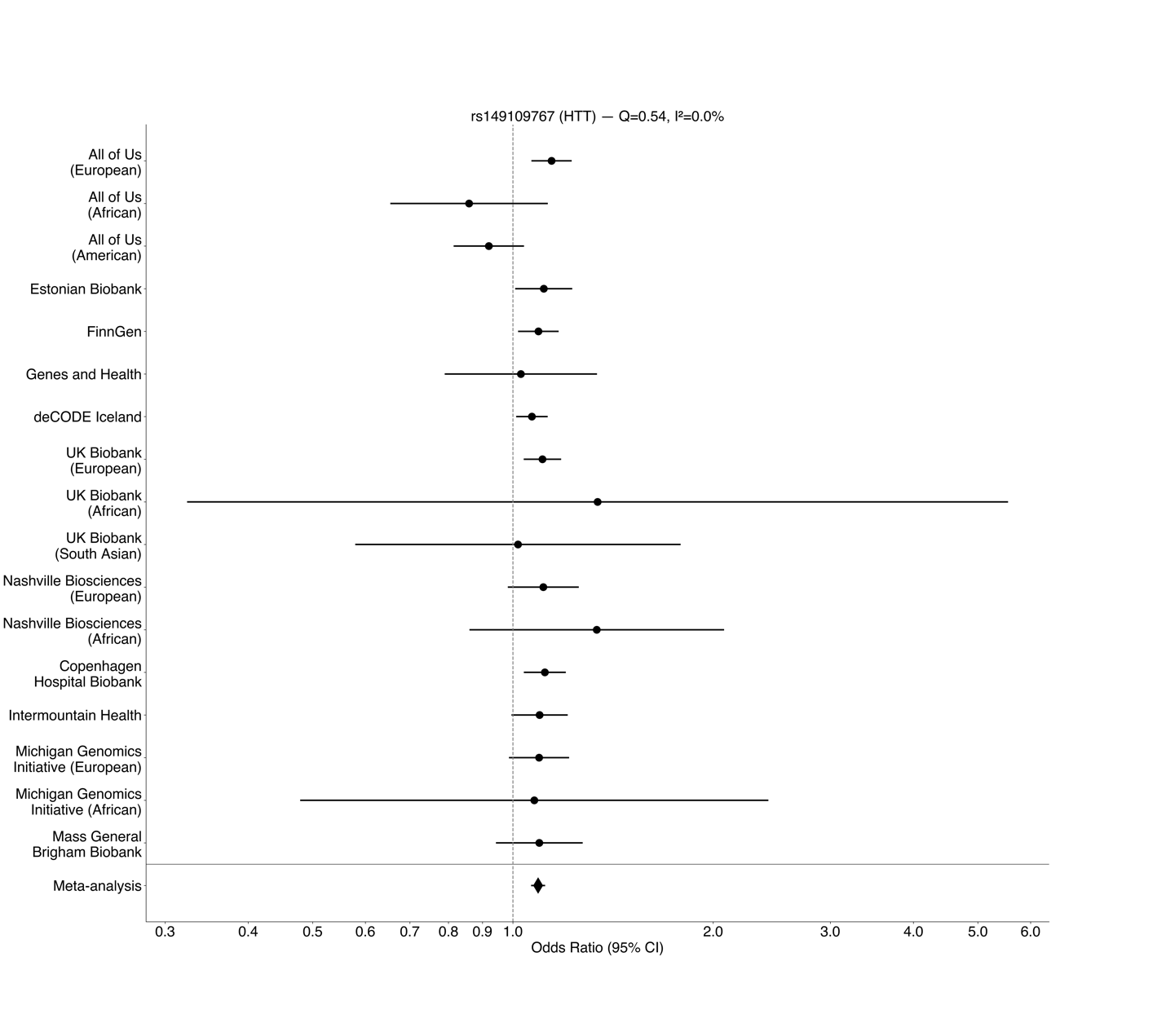

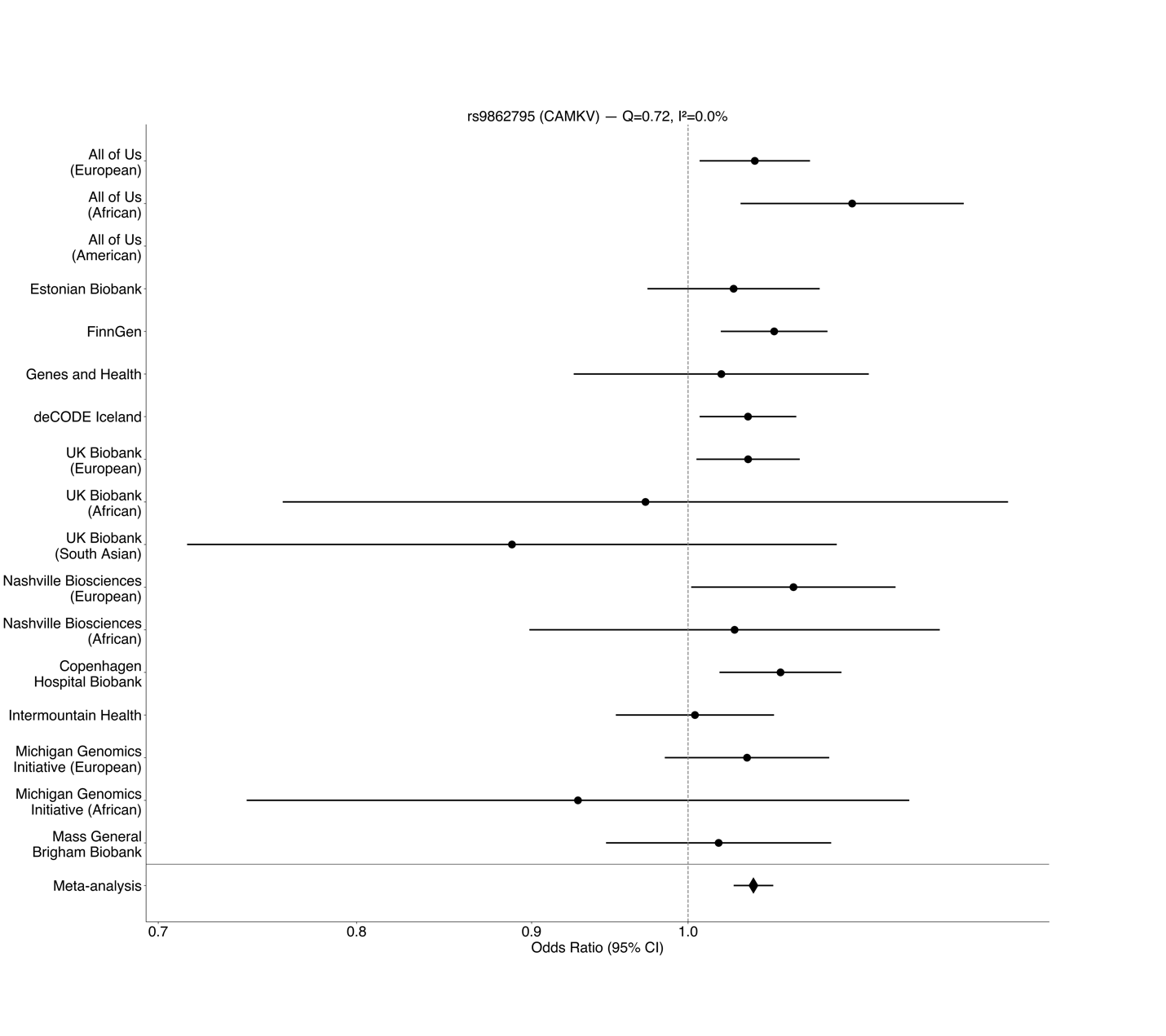

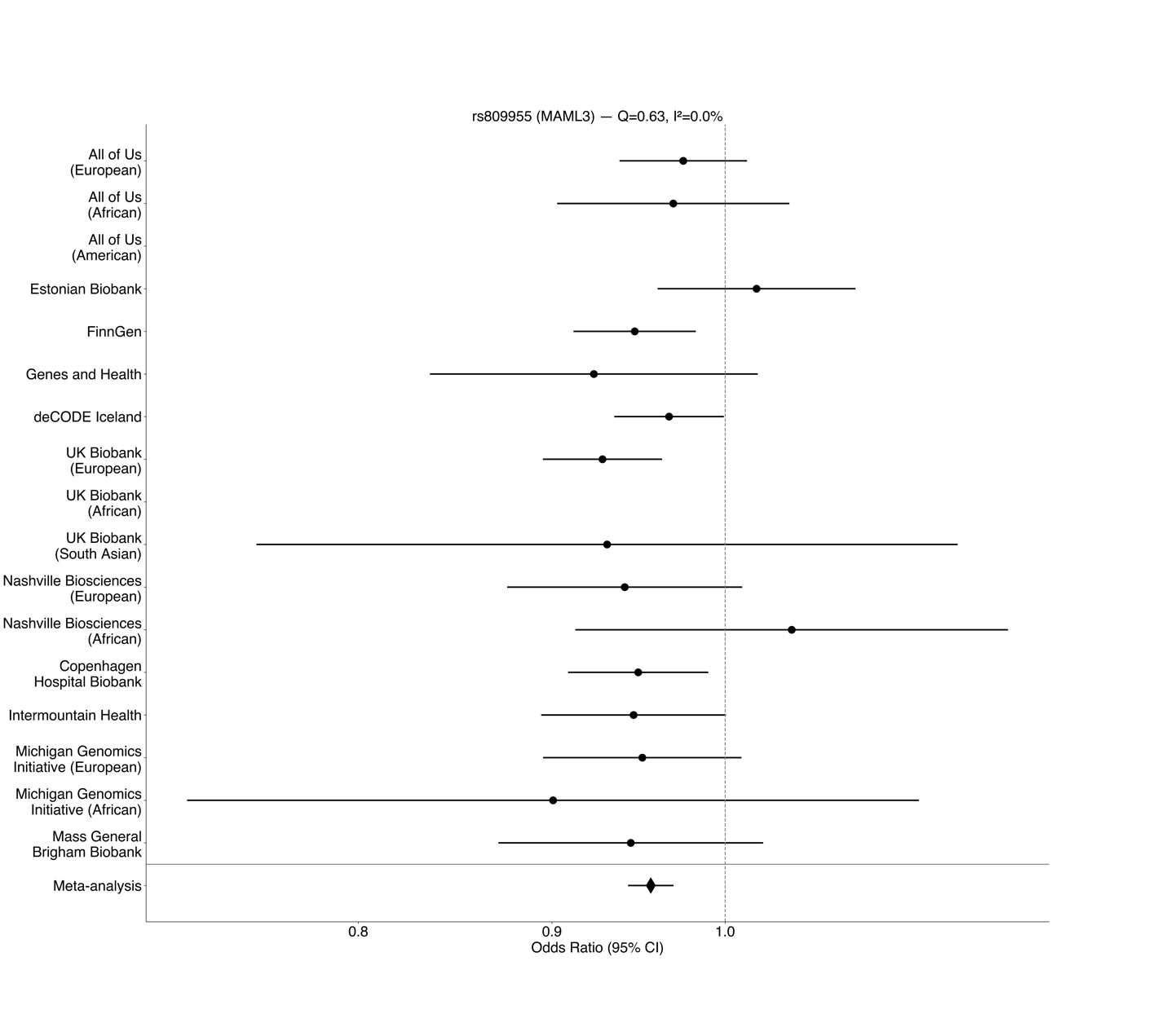

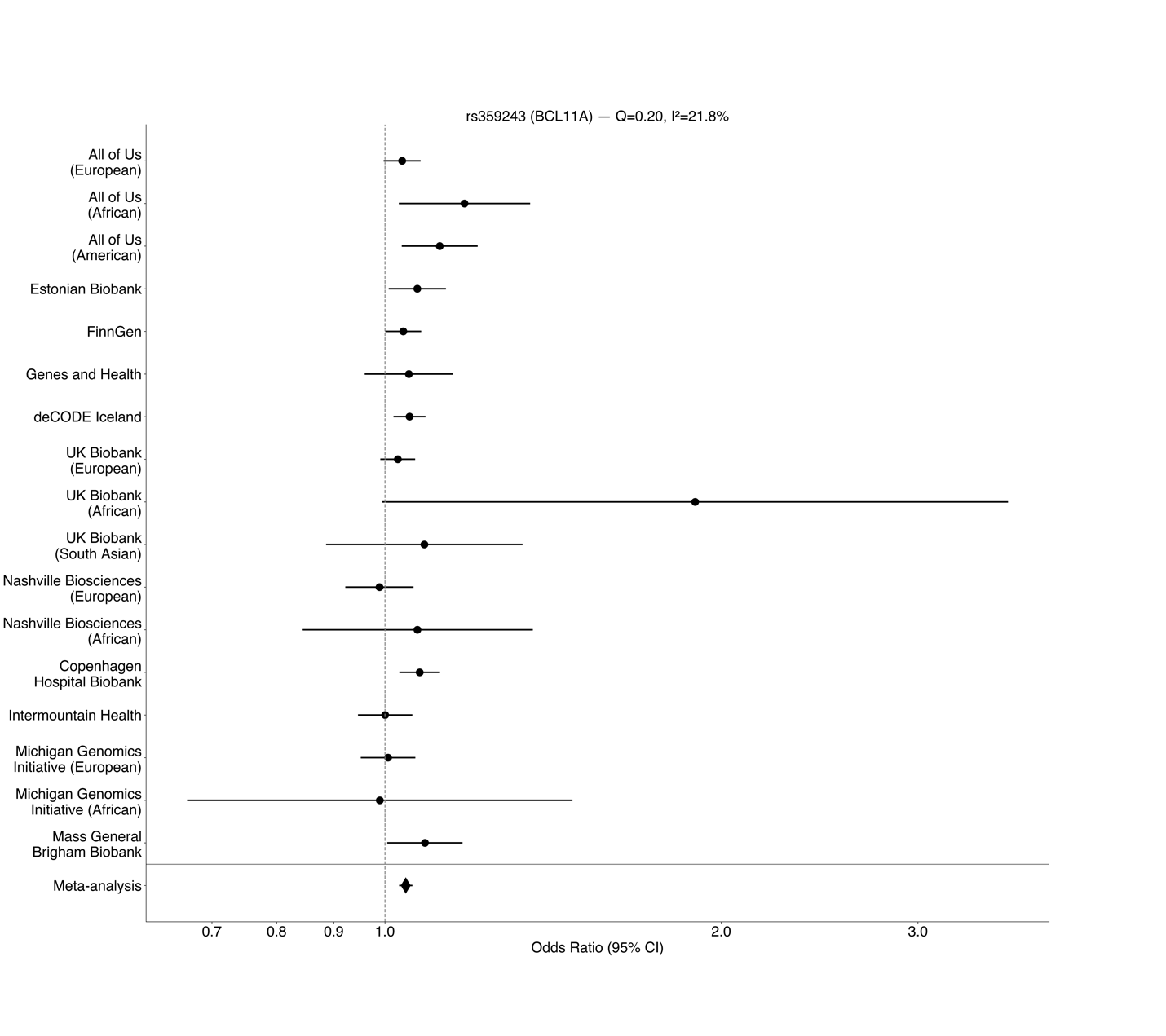

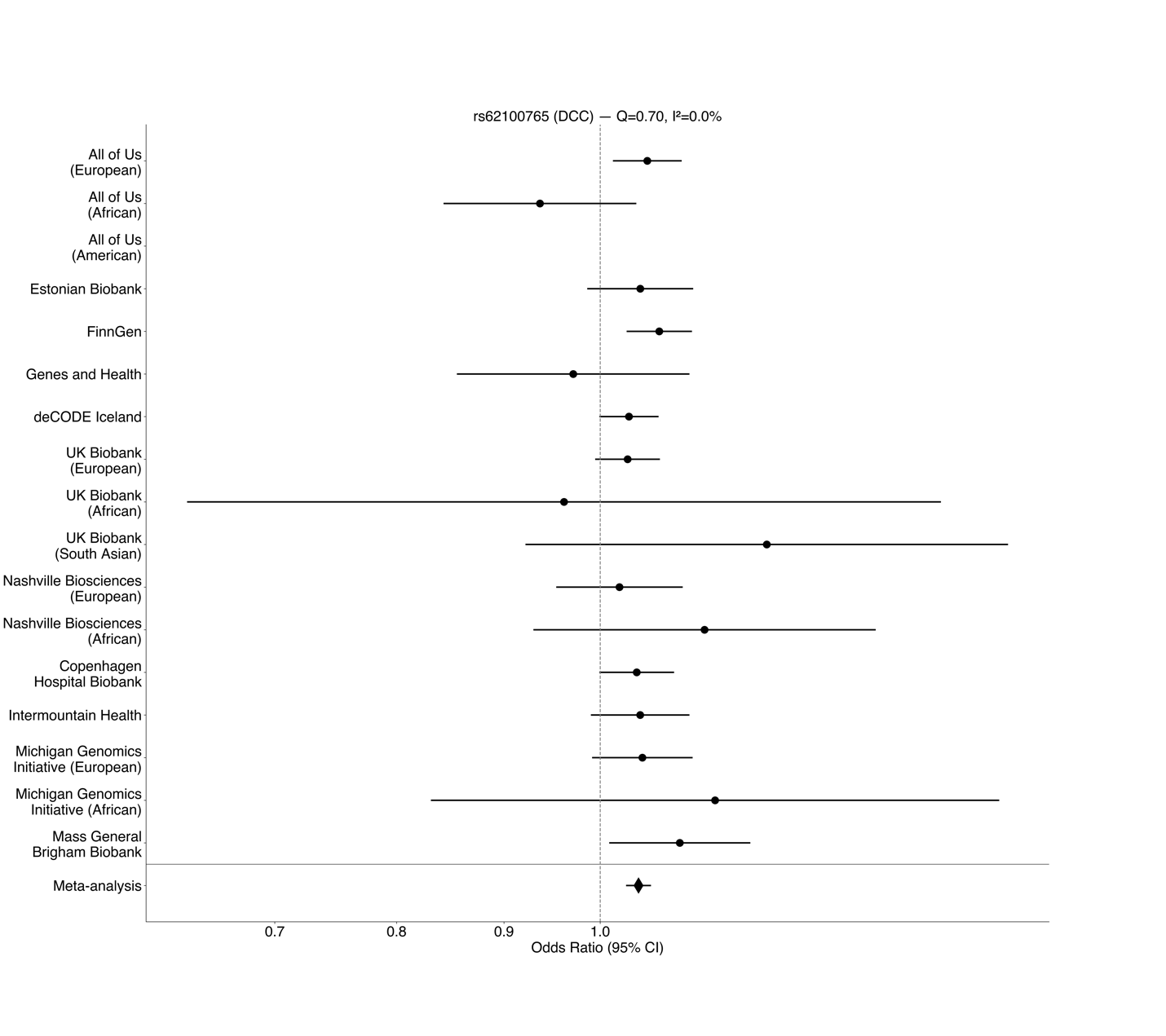

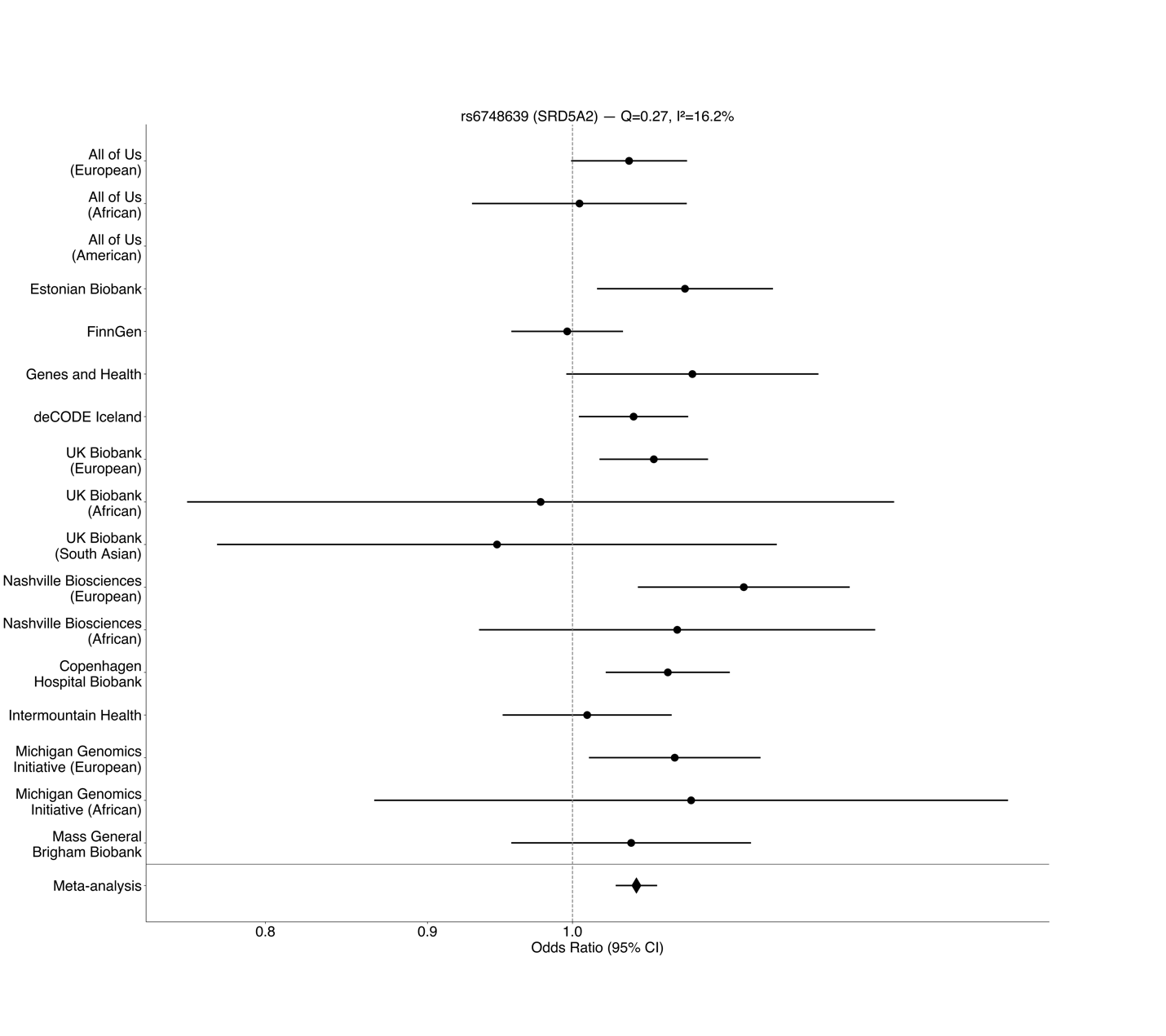

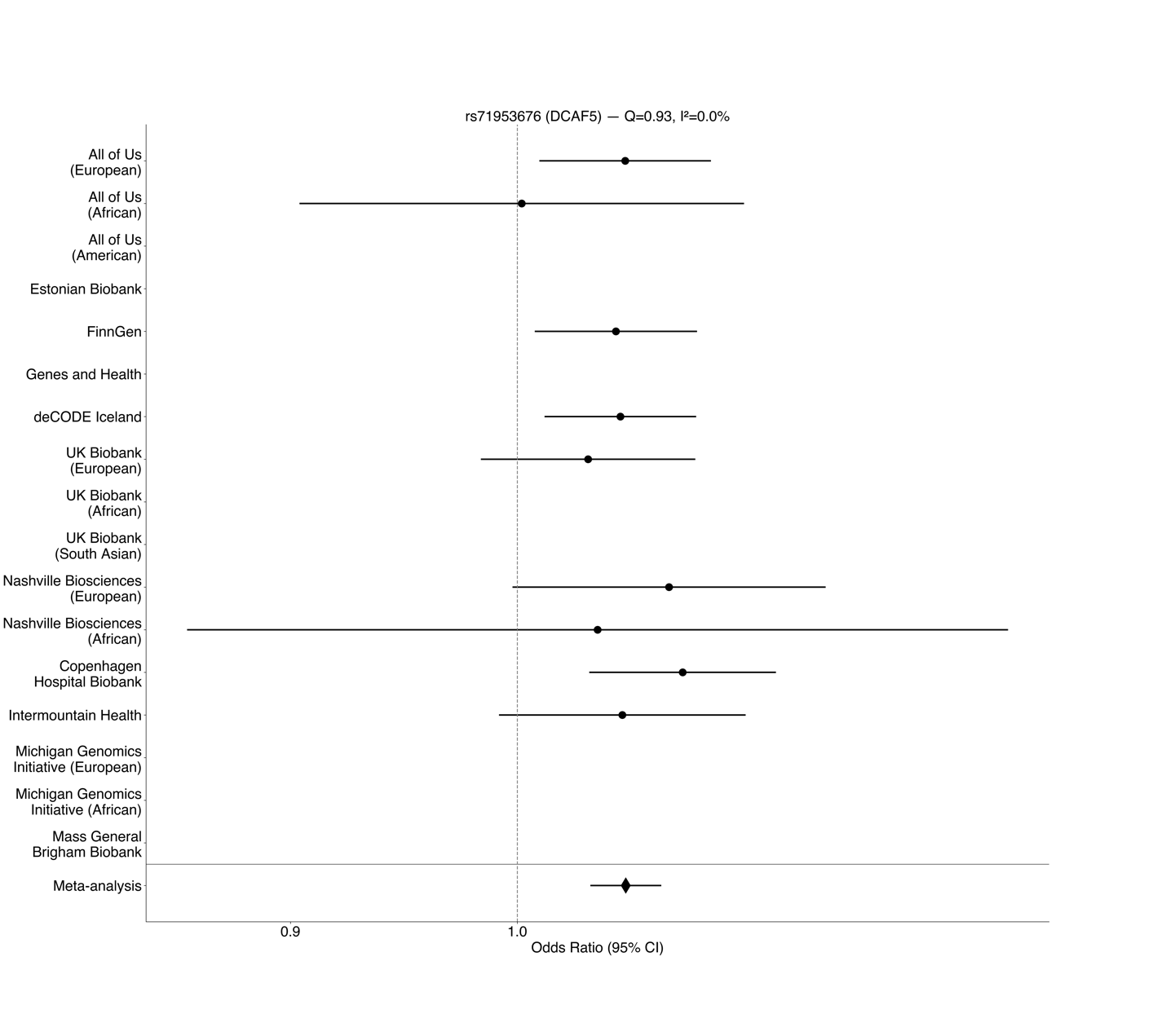

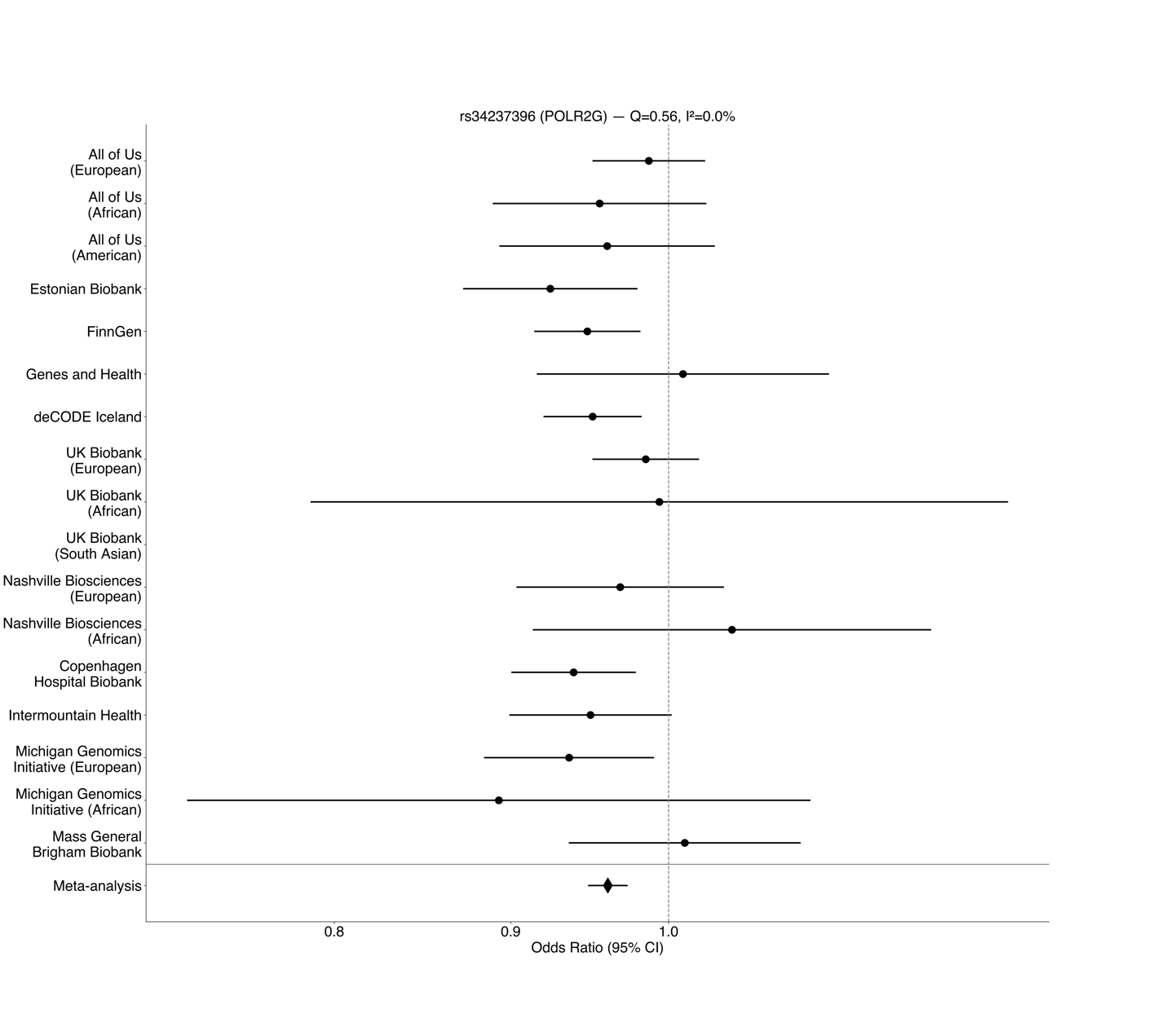

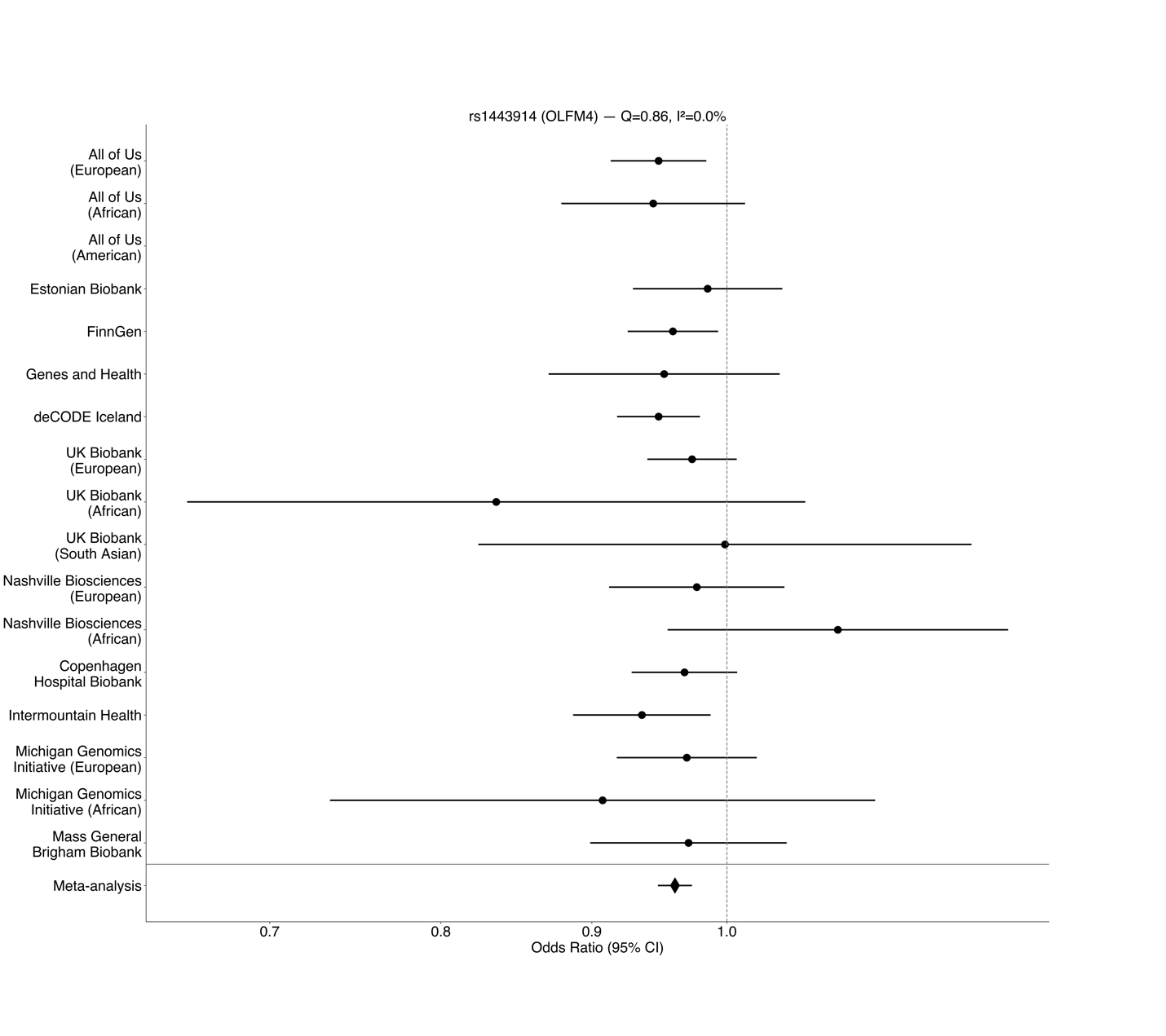

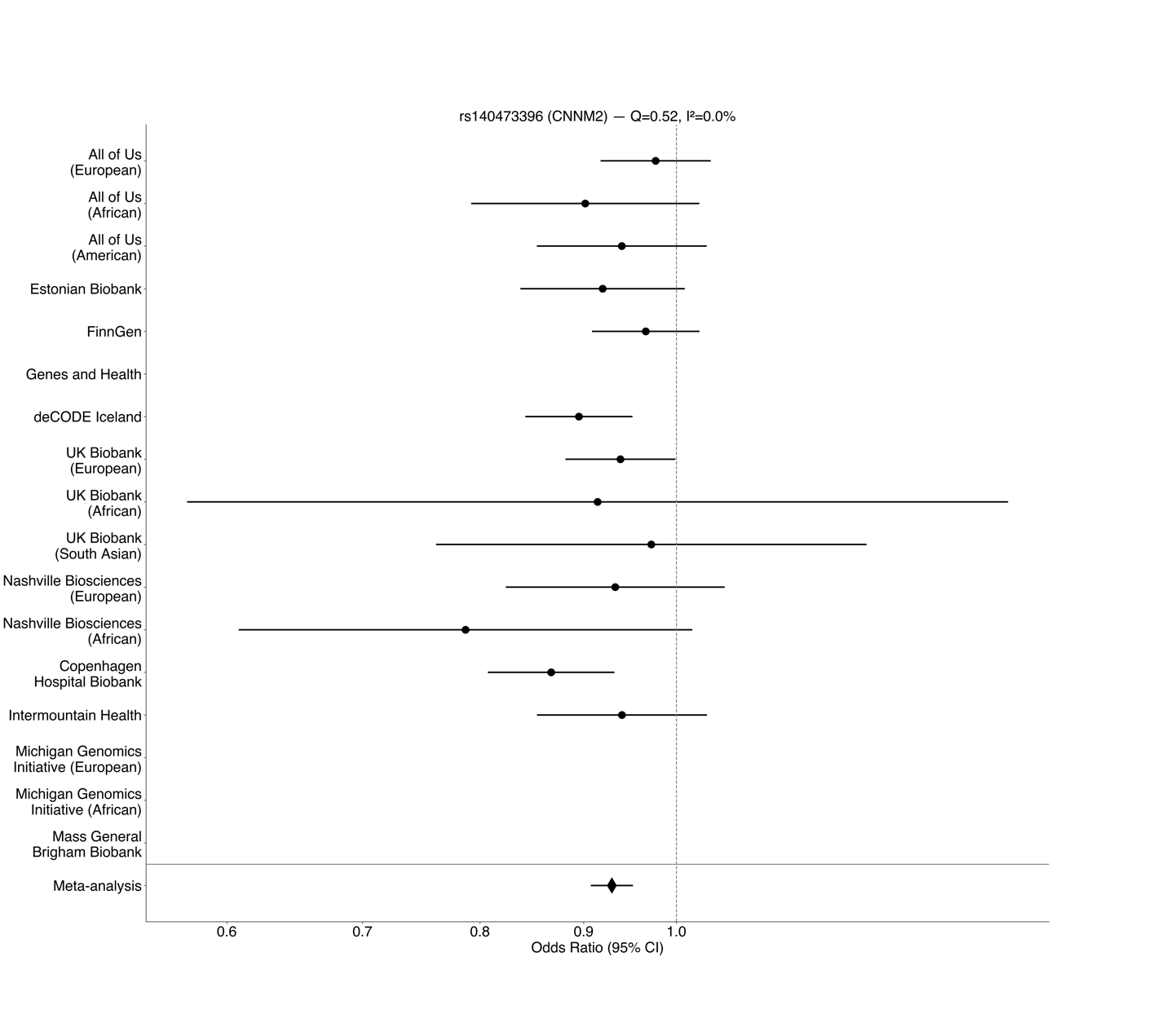

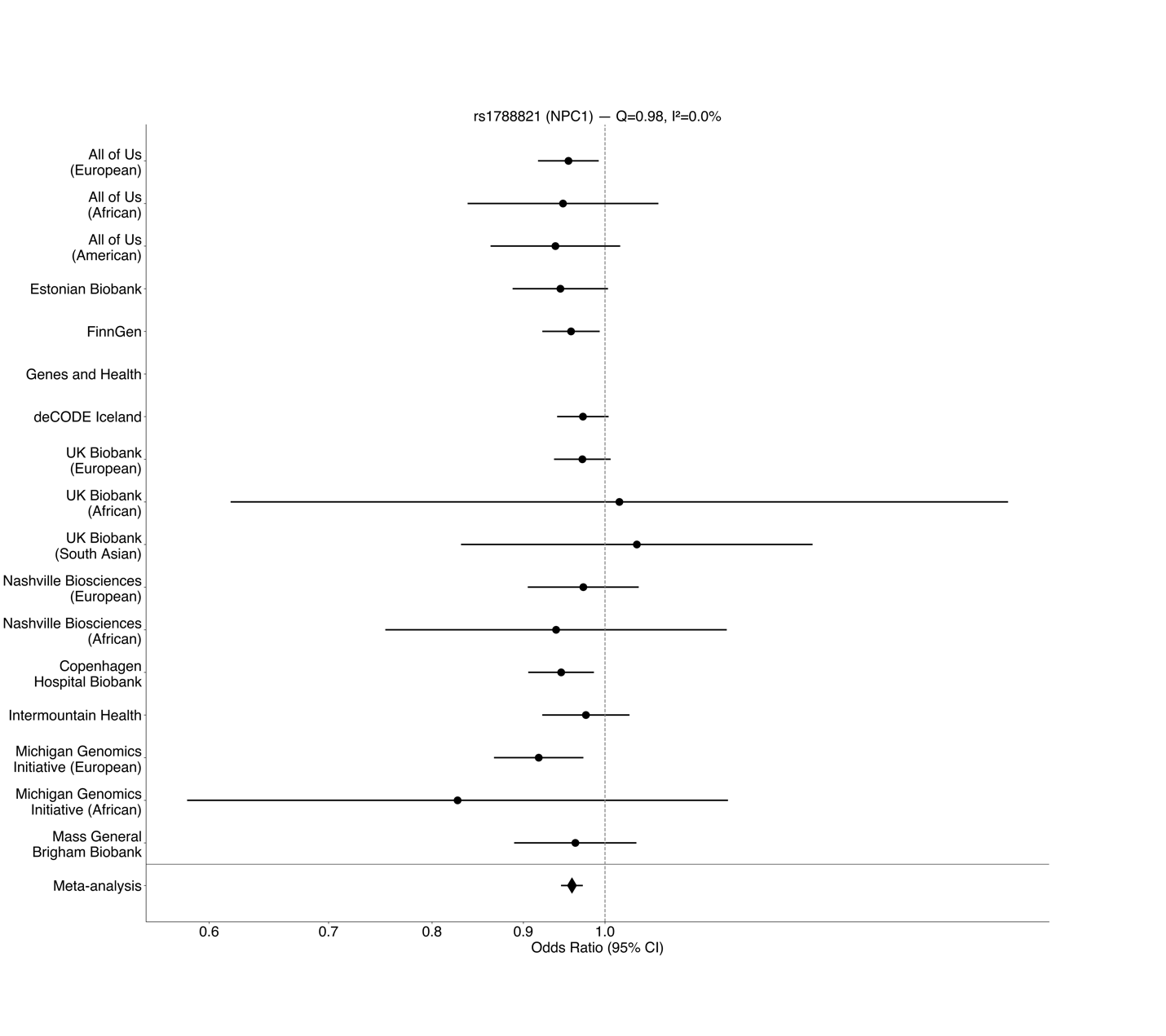

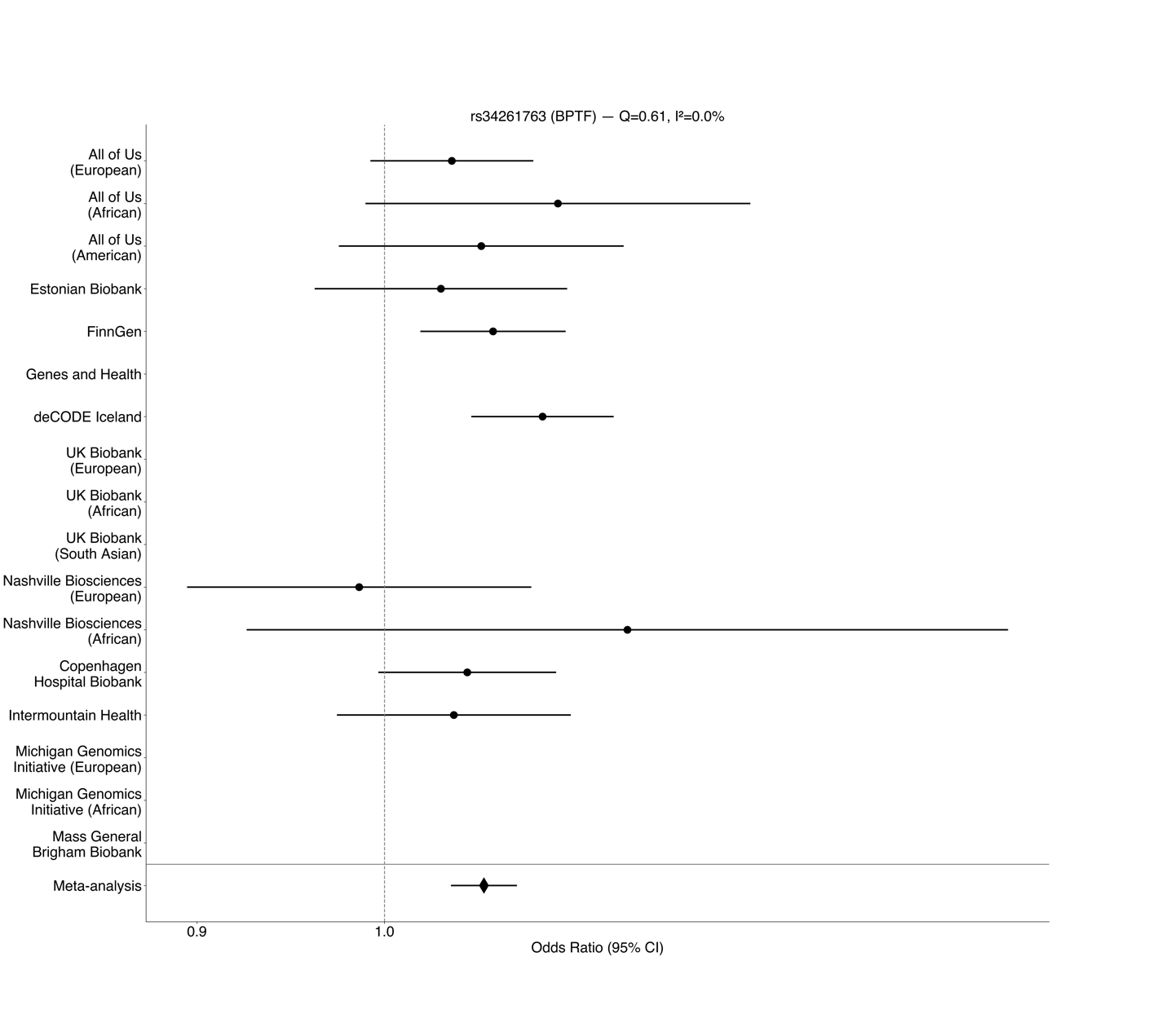

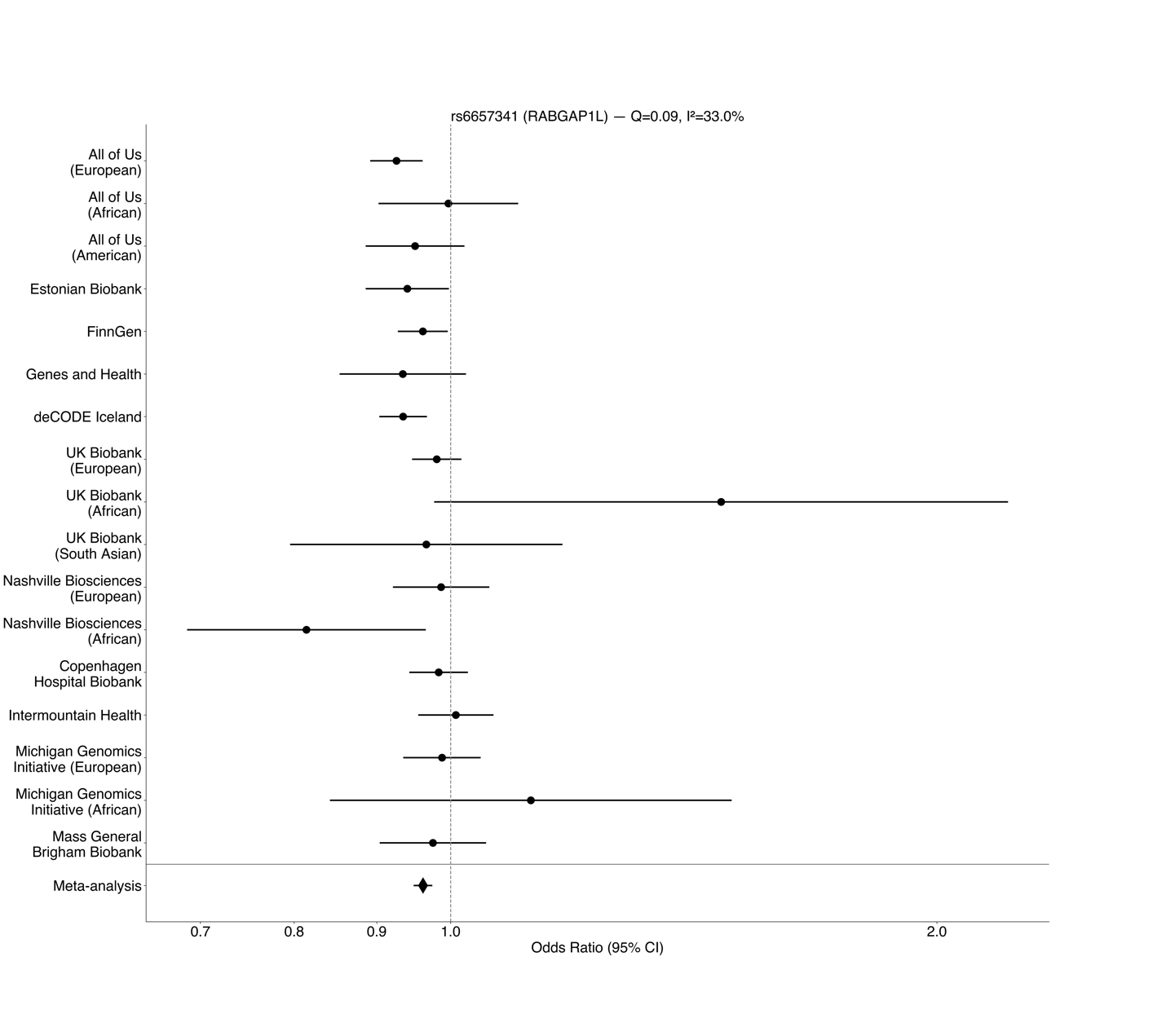

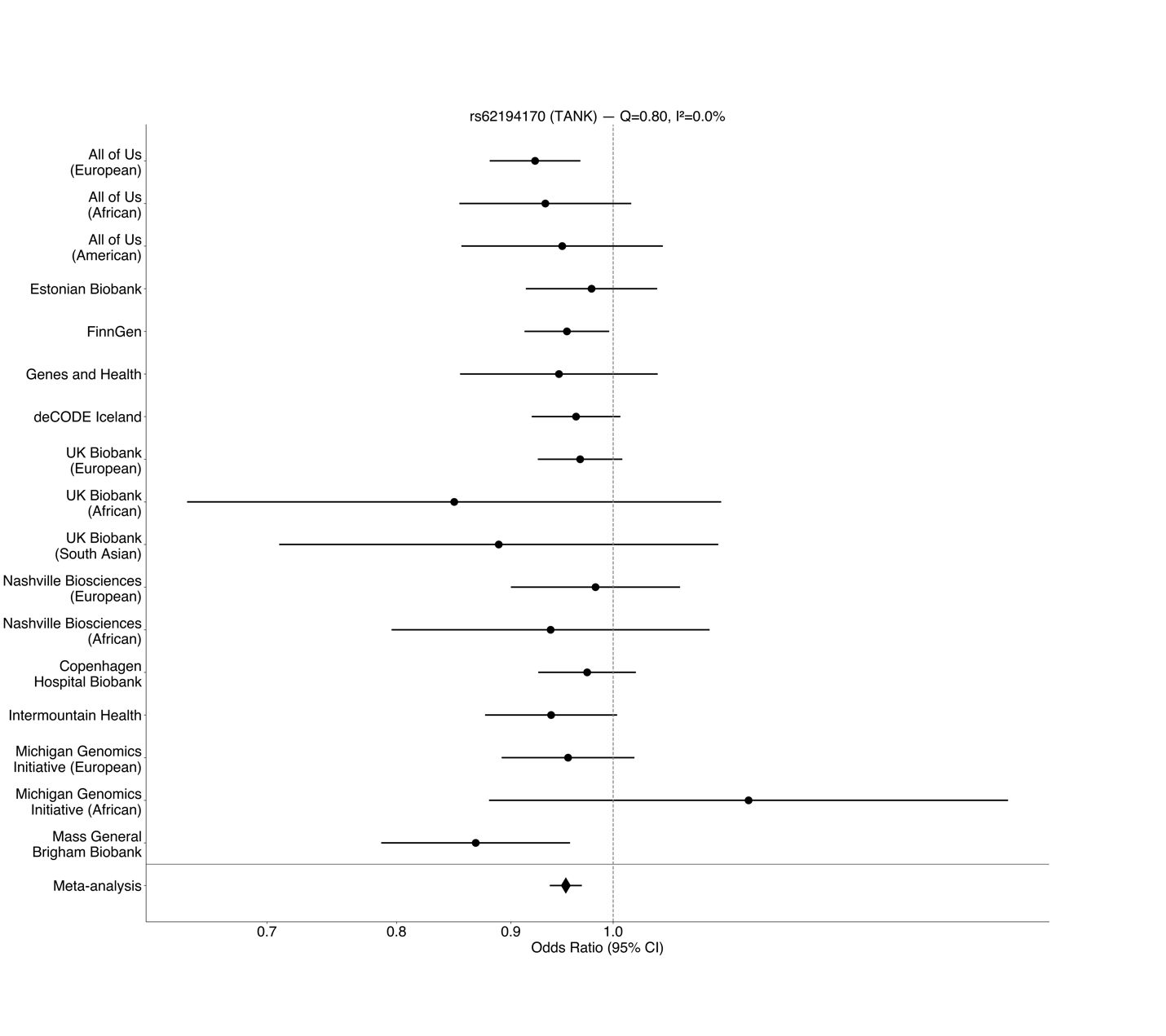

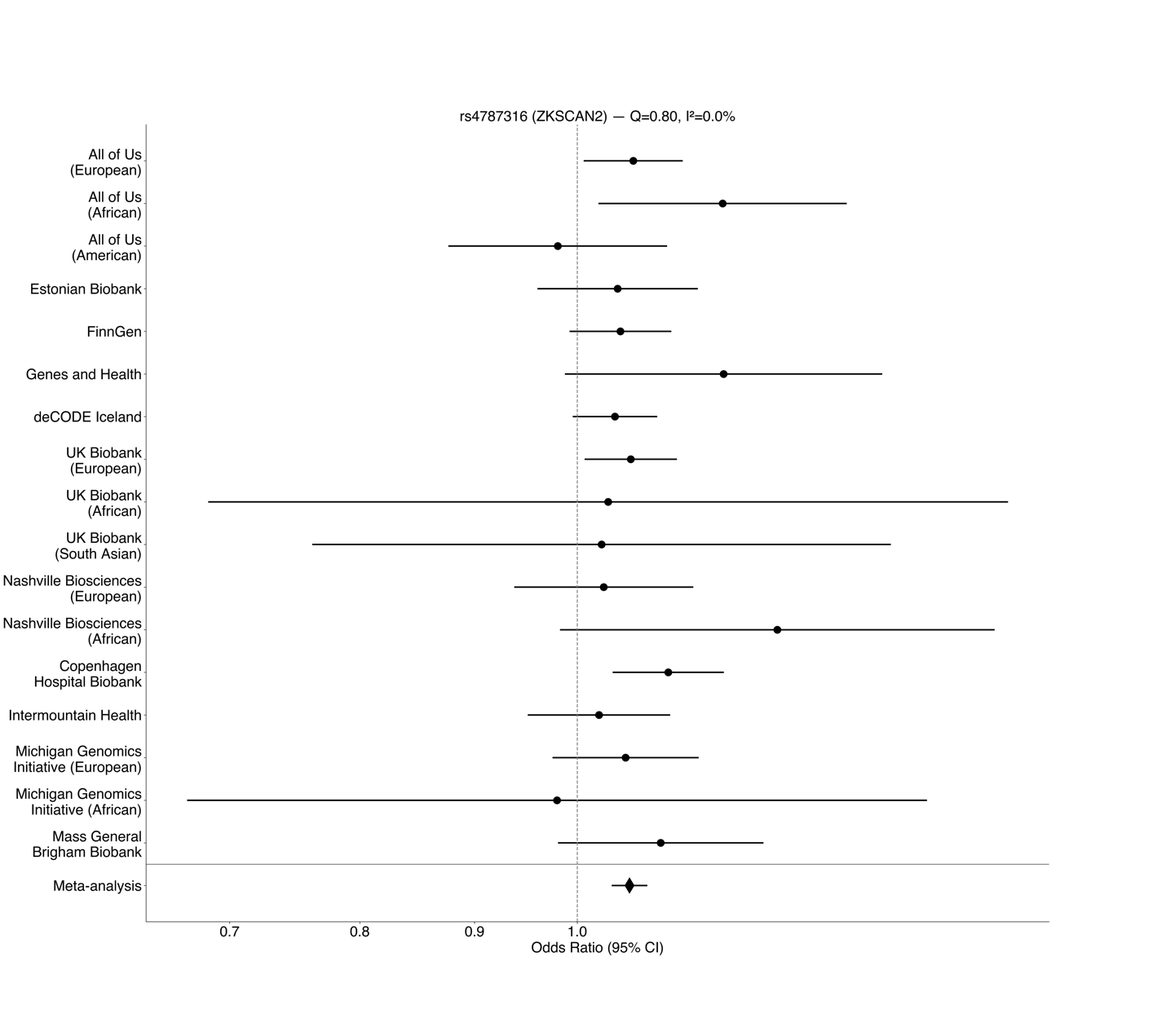

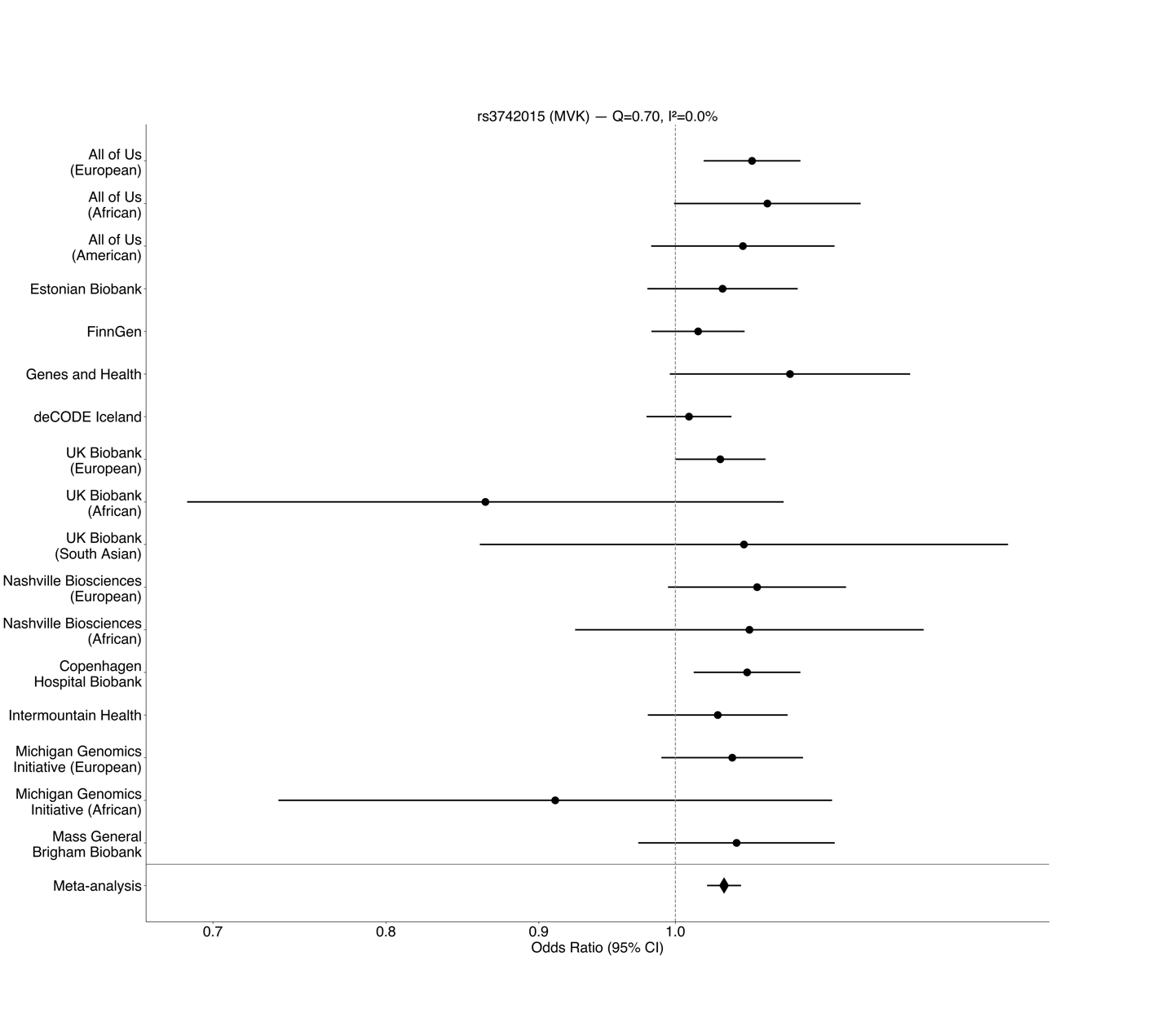

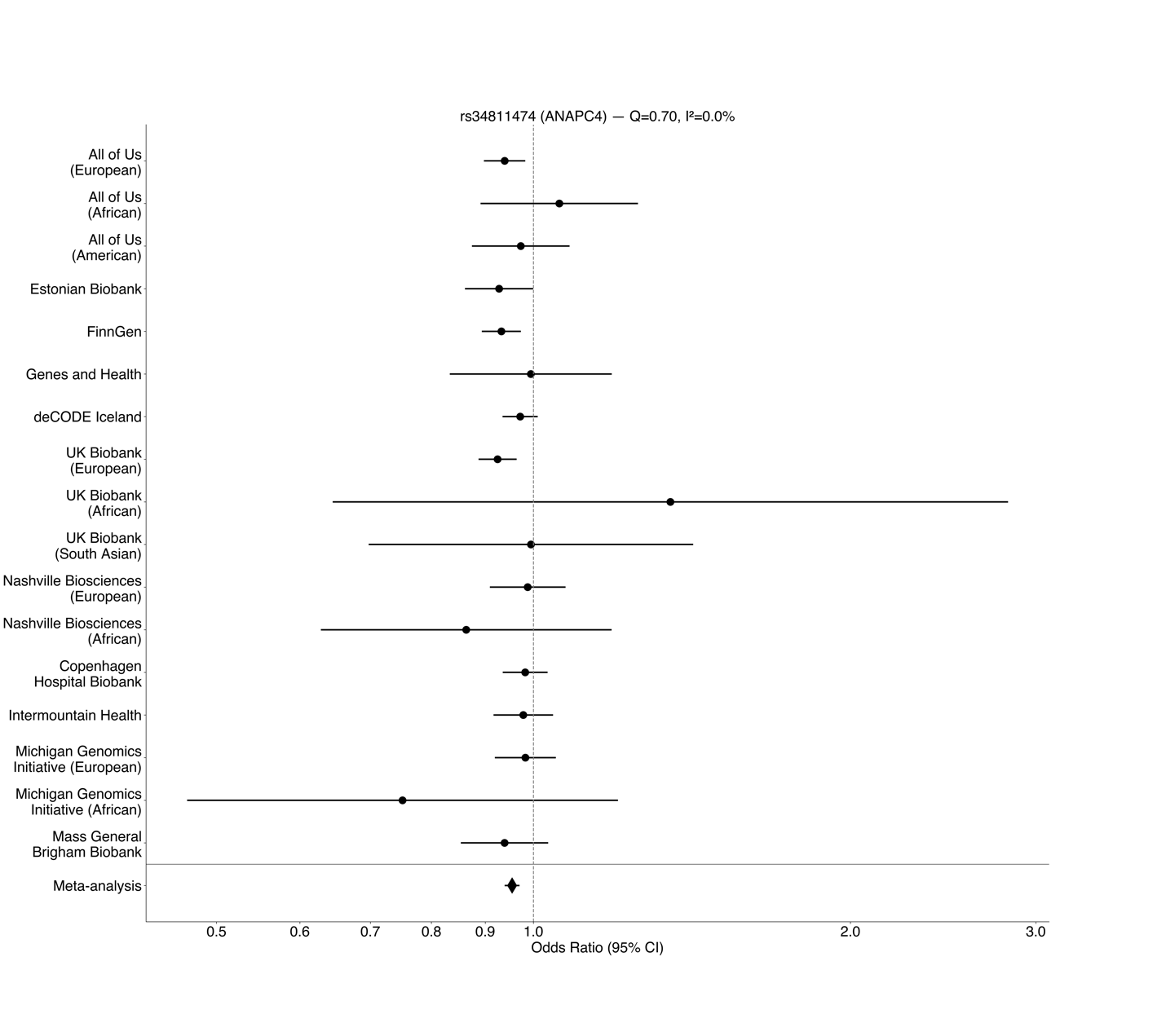

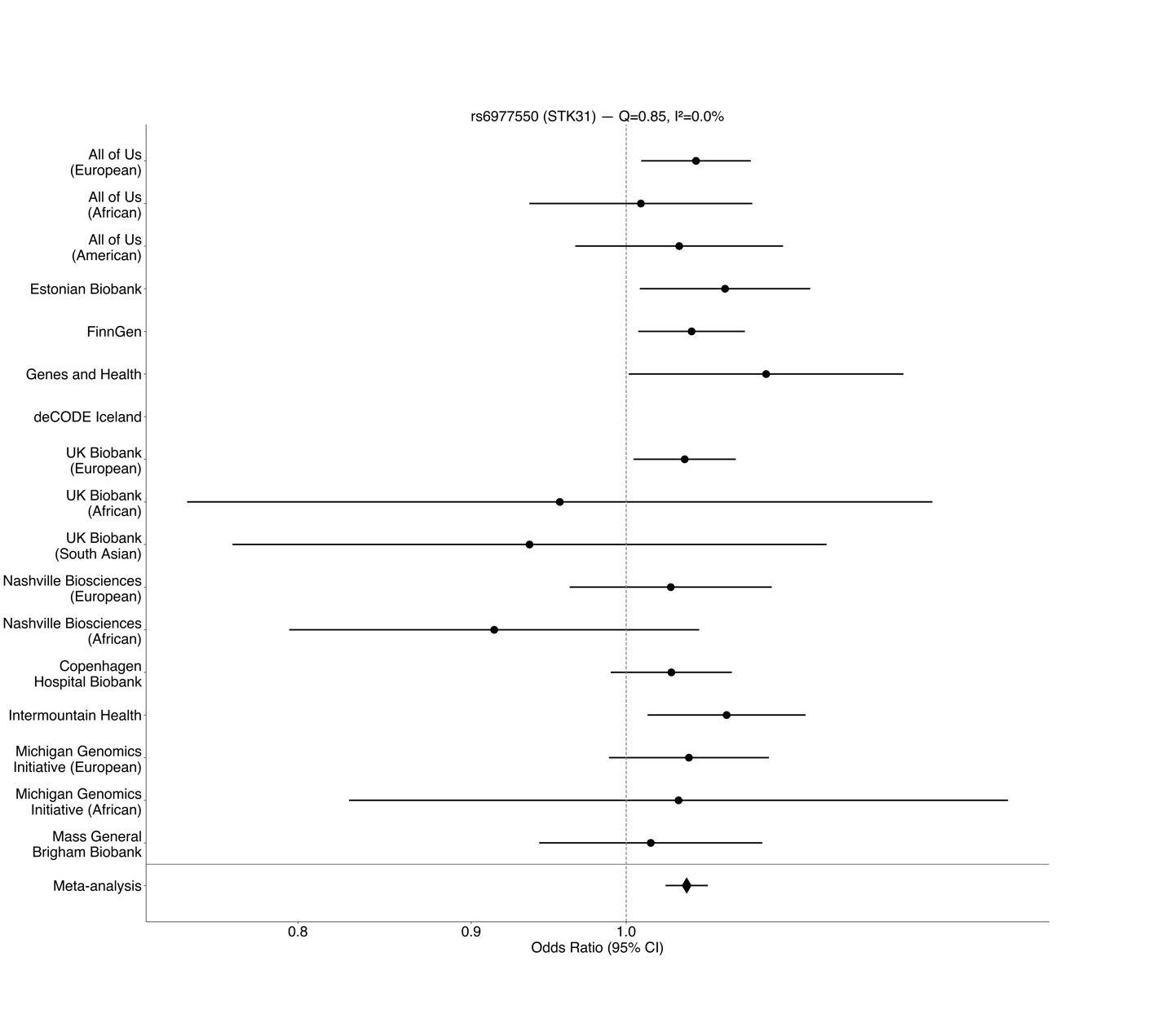

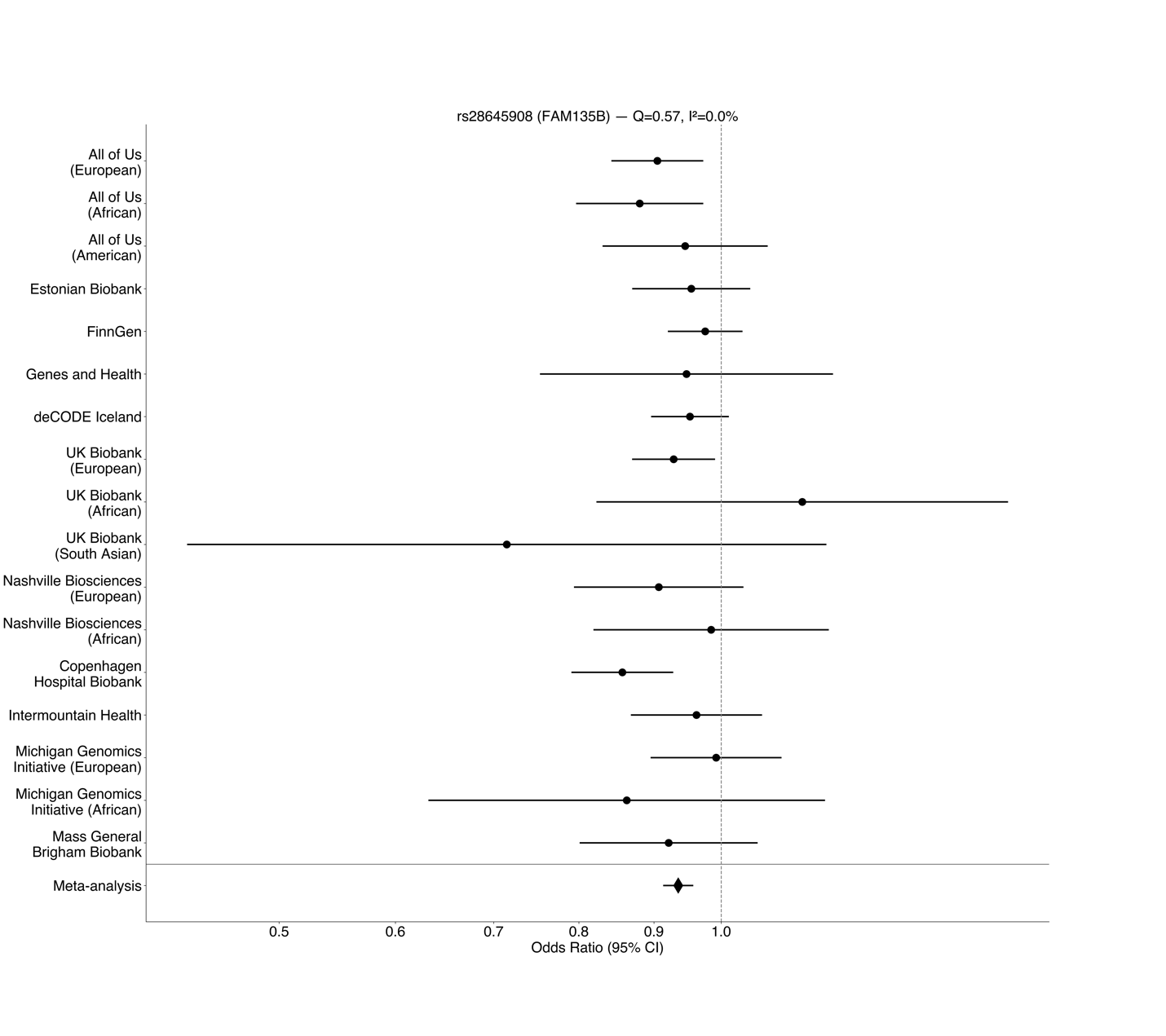

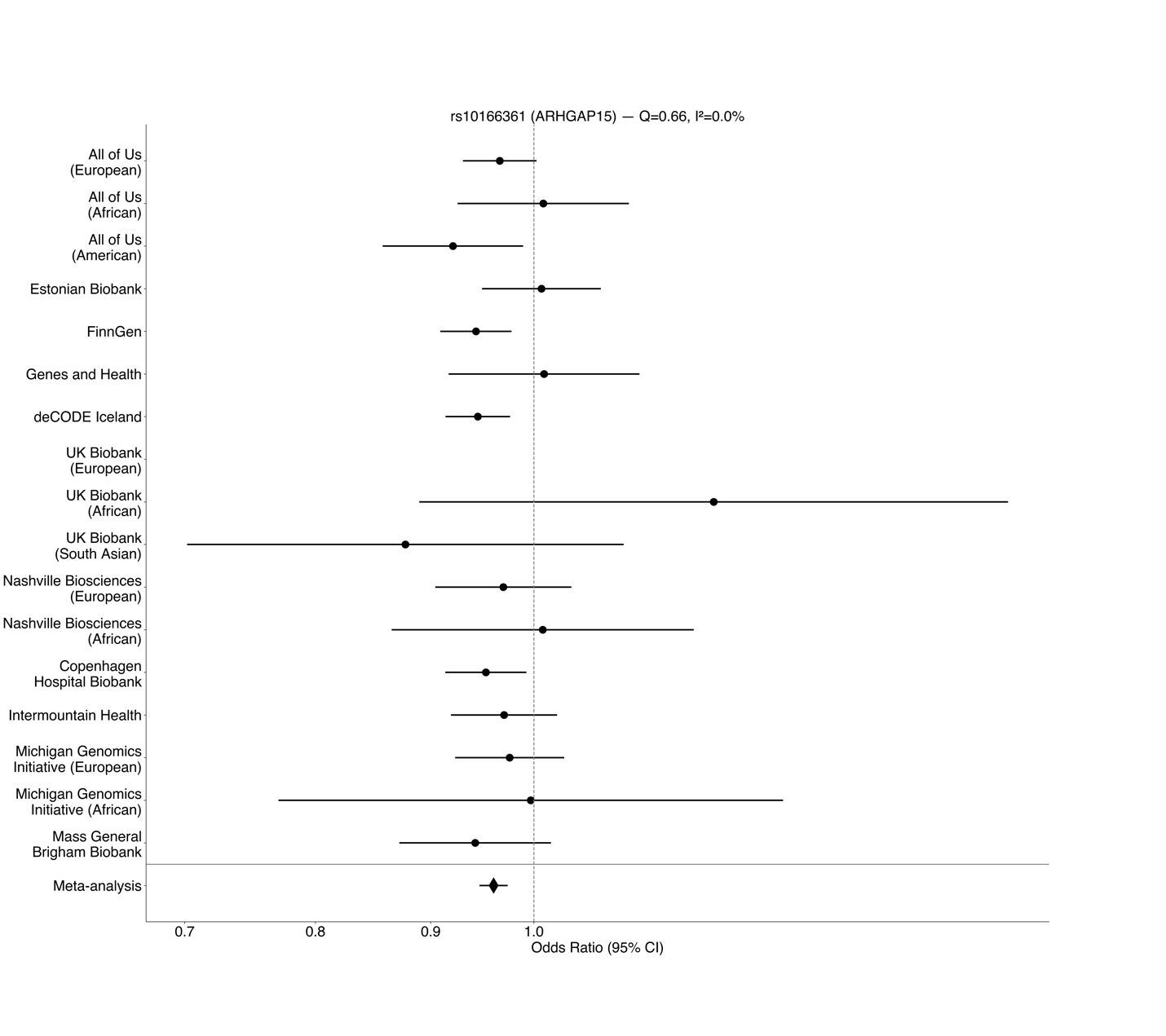

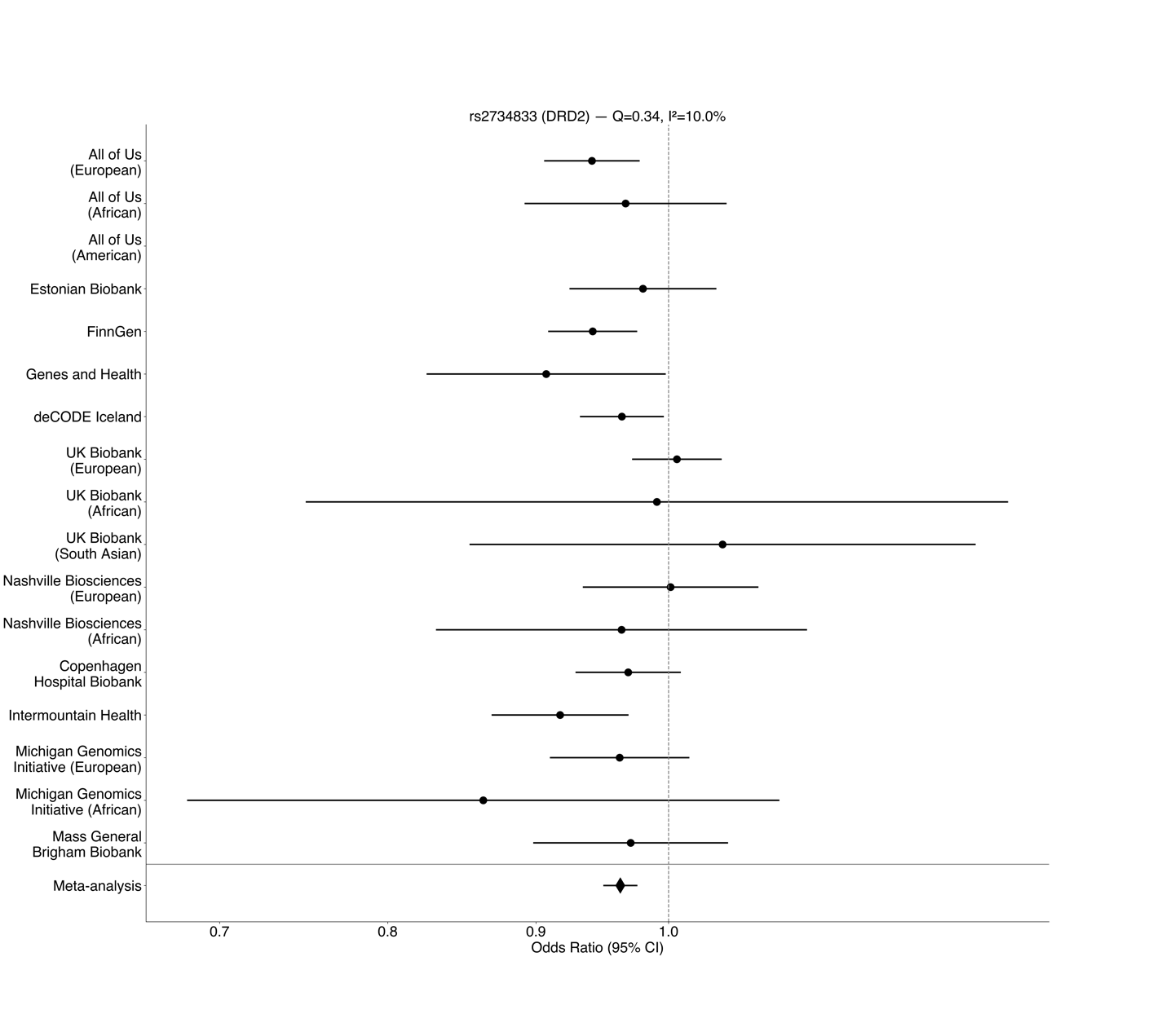

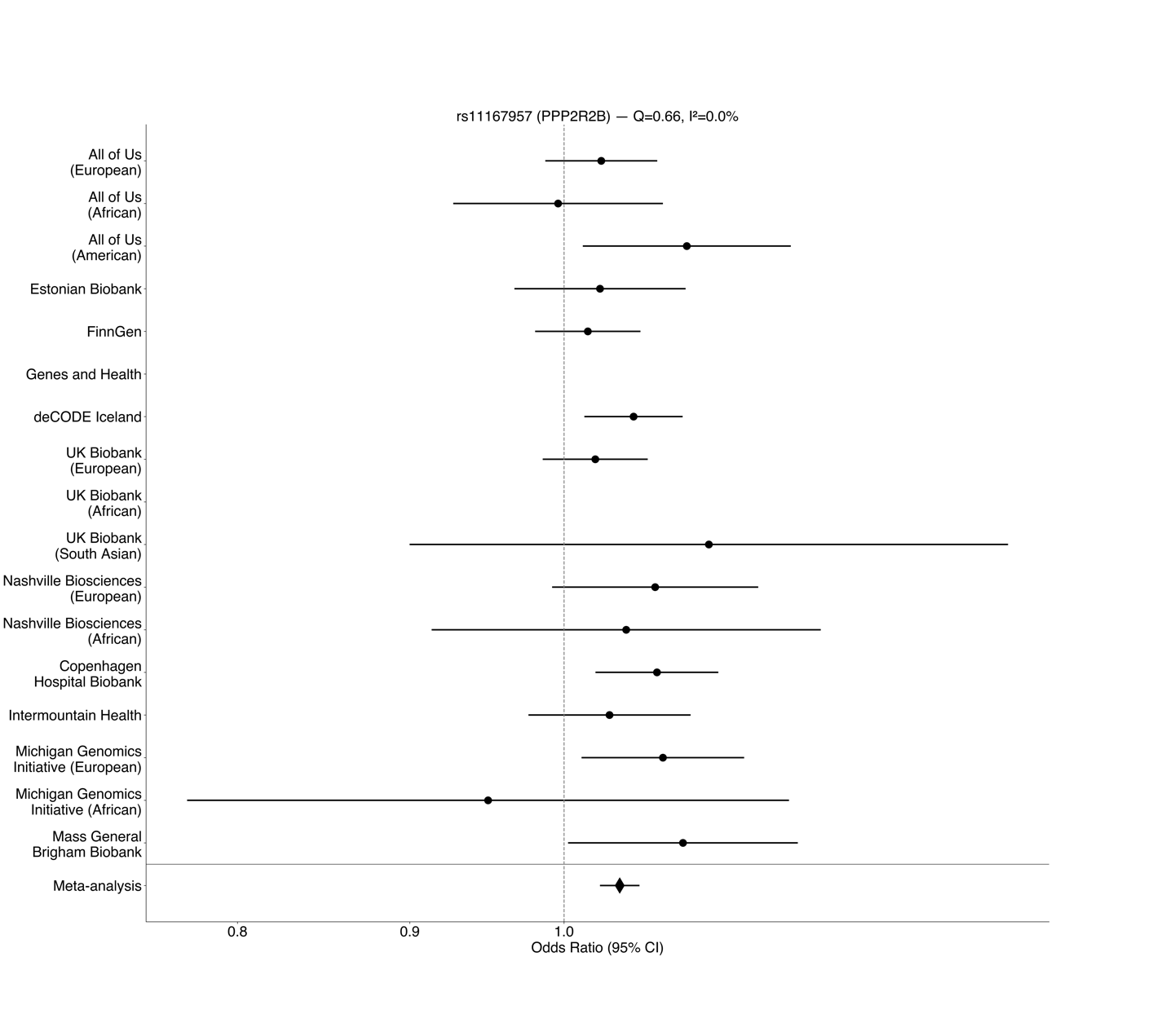

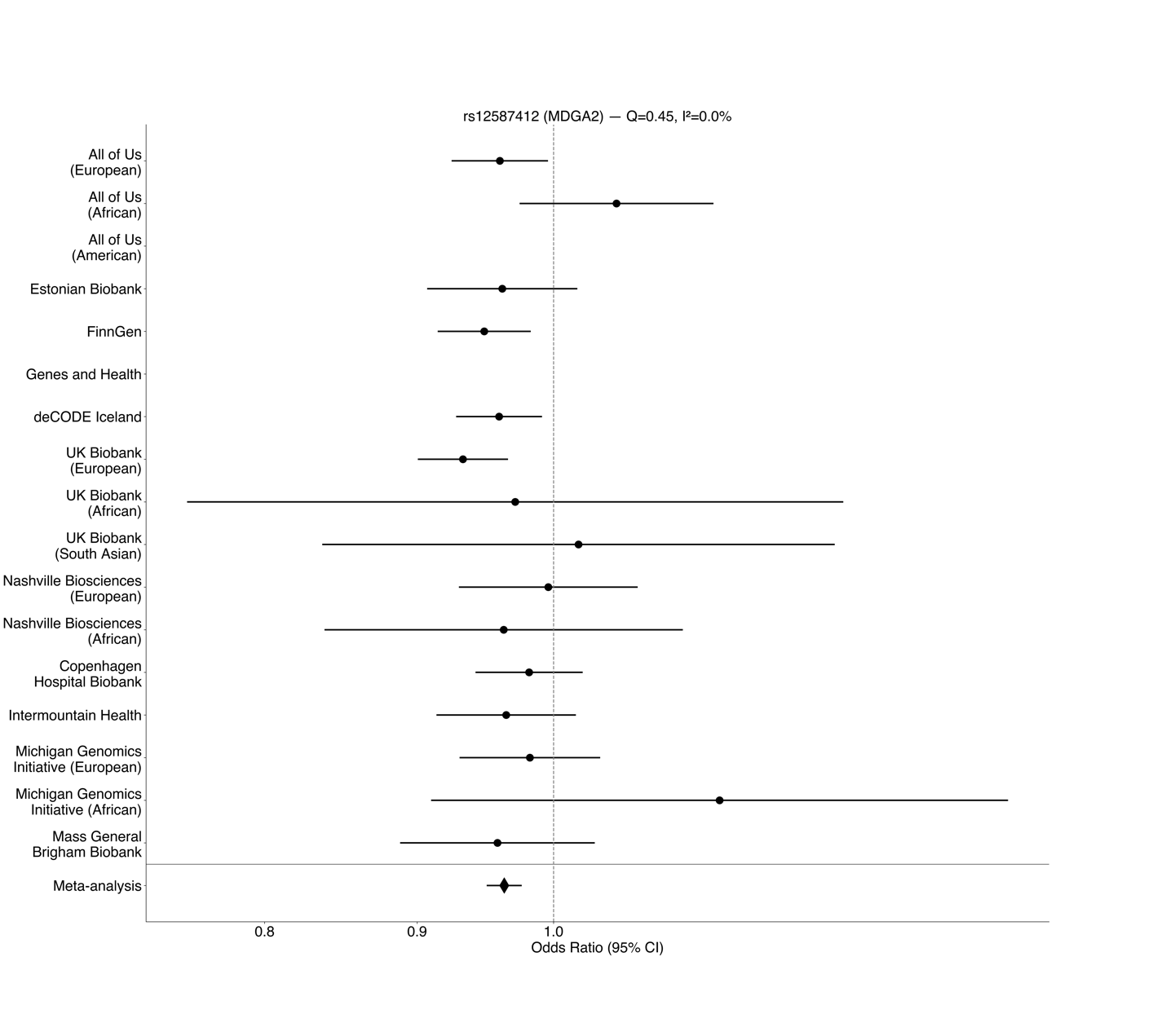

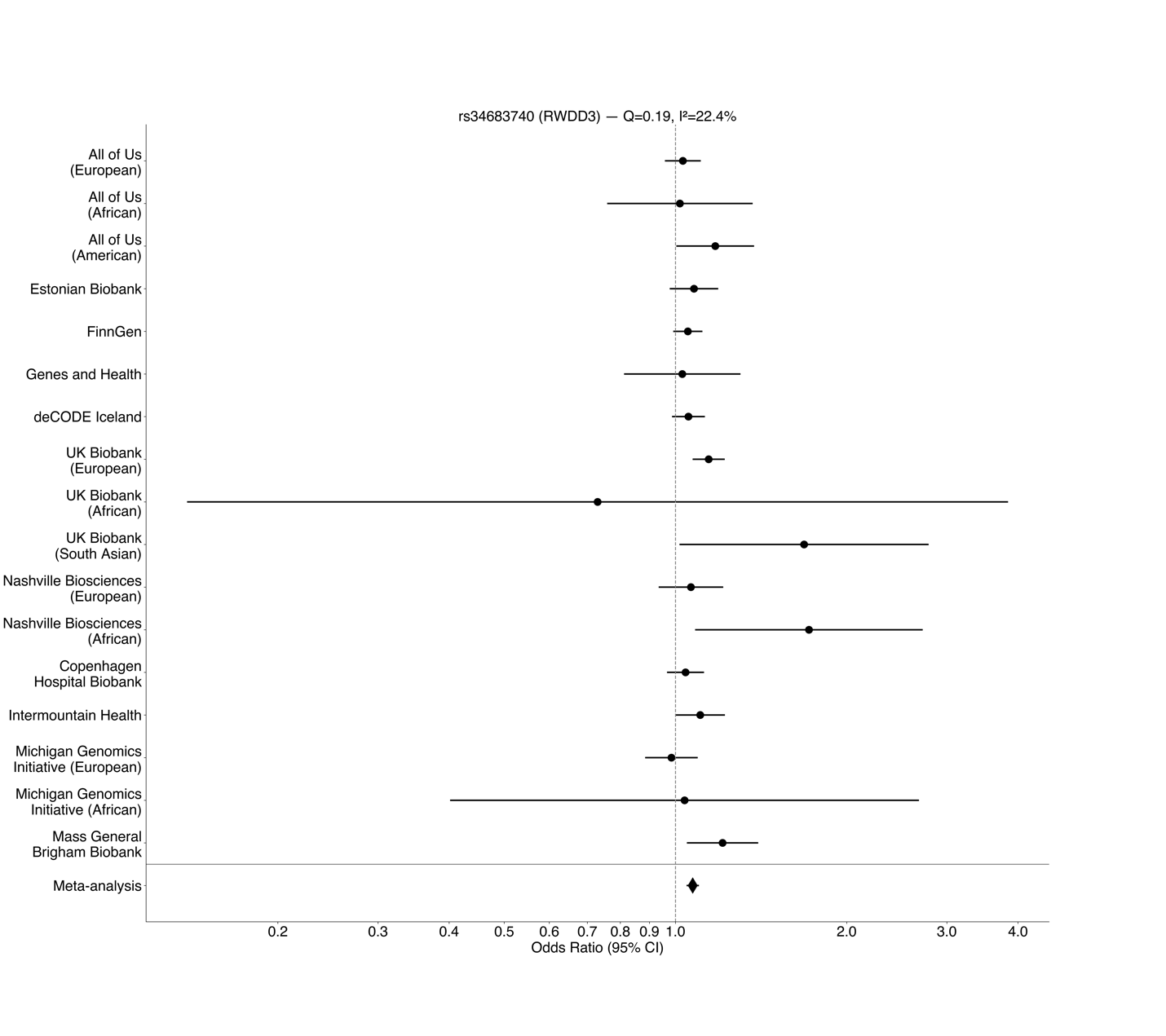

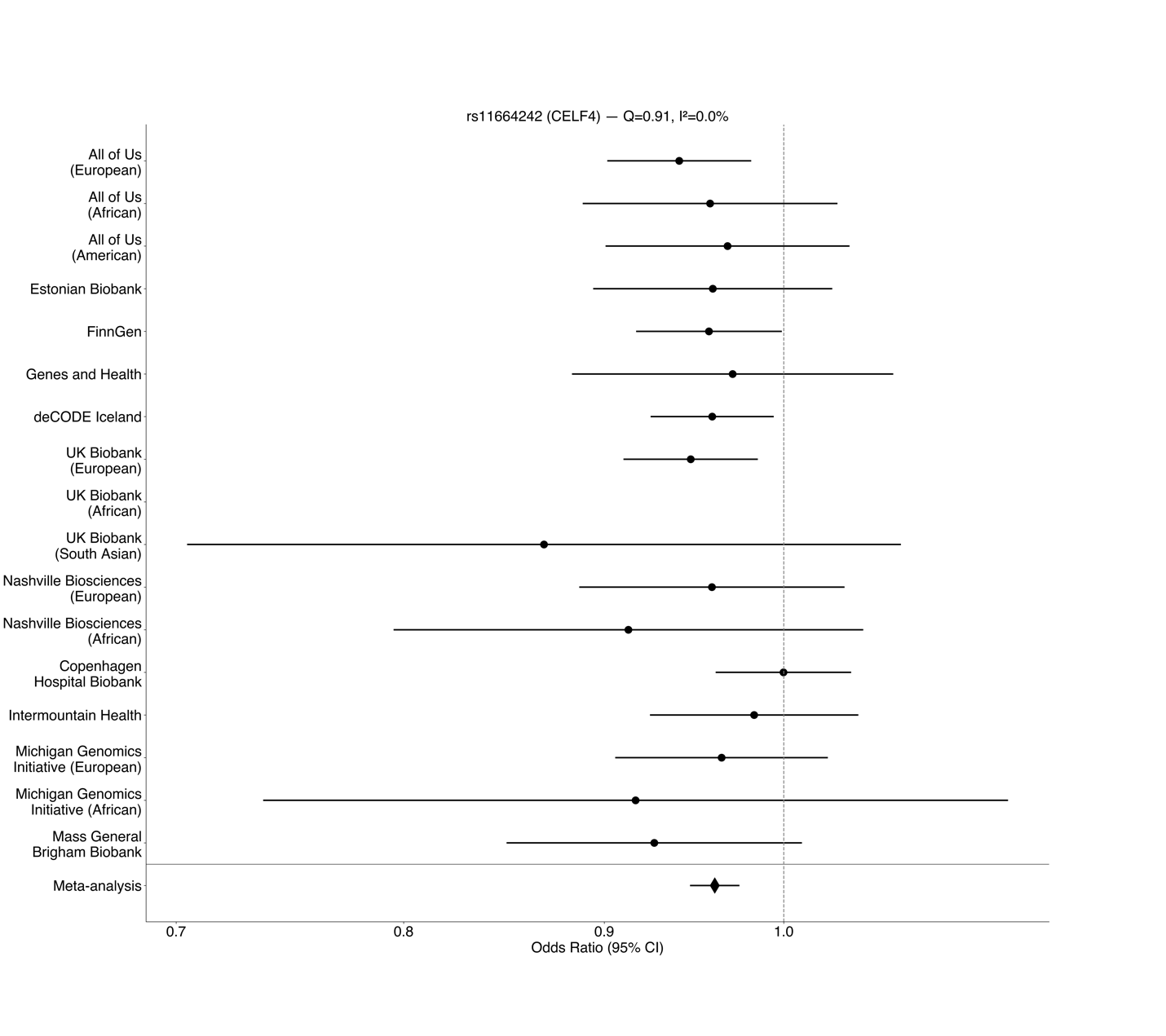

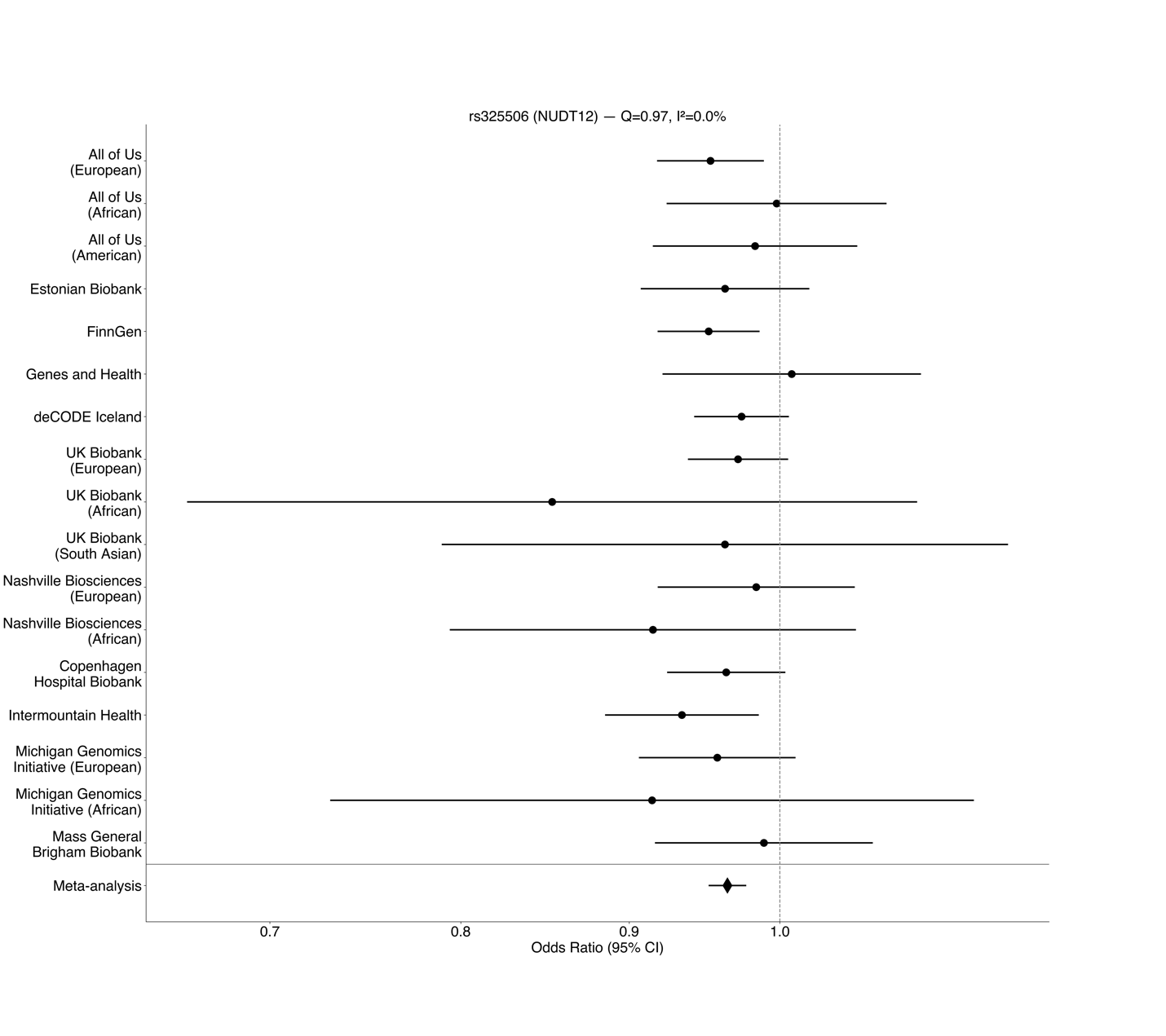
**Figure S1: Forest plots of lead variants from the multi-ancestry meta analysis.** “Copenhagen Hospital Biobank” is short for “Copenhagen Hospital Biobank and Danish Blood Donor Study”.

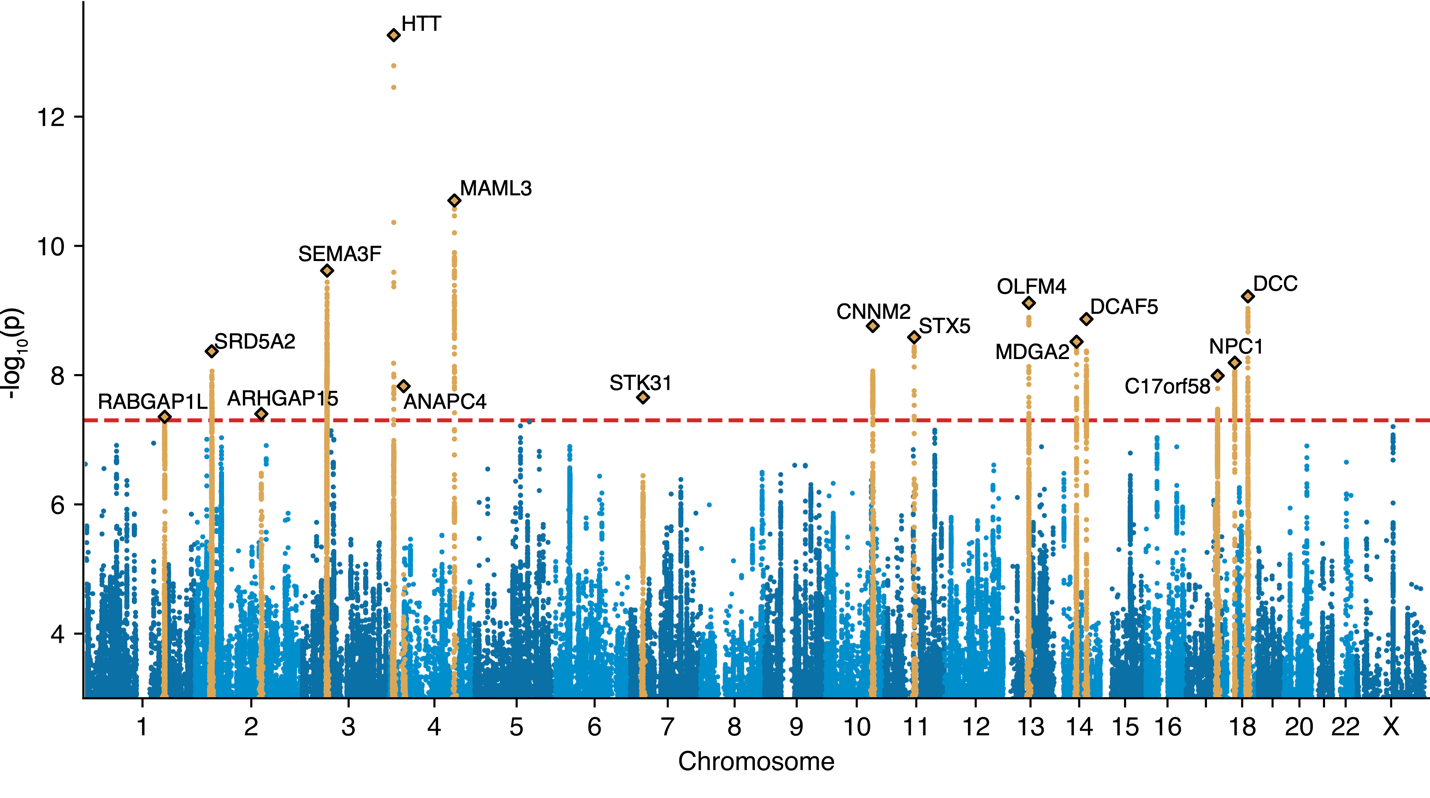

**Figure S2: Manhattan plot of the European-only meta-analysis.** Gold-highlighted variants have linkage disequilibrium r^2^ > 0.001 and are within 5 megabases of the lead variants (diamonds). The nearest gene to each lead variant is labeled.

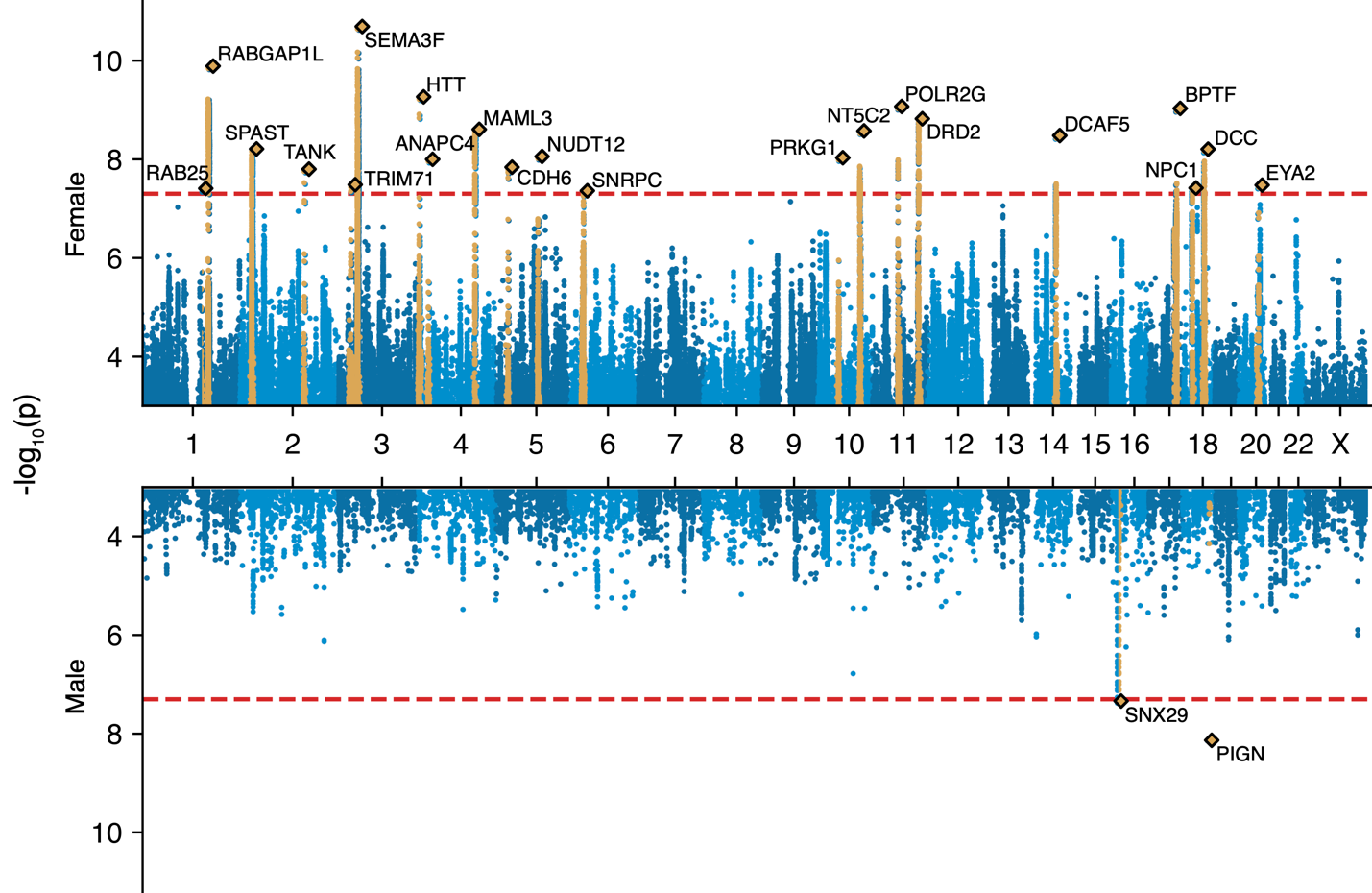

**Figure S3: Miami plot of the multi-ancestry female-only and male-only meta-analyses.** Gold-highlighted variants have linkage disequilibrium r^2^ > 0.001 and are within 5 megabases of the lead variants (diamonds). Note that males are European only, due to low male case counts in non-European ancestries (see Methods).

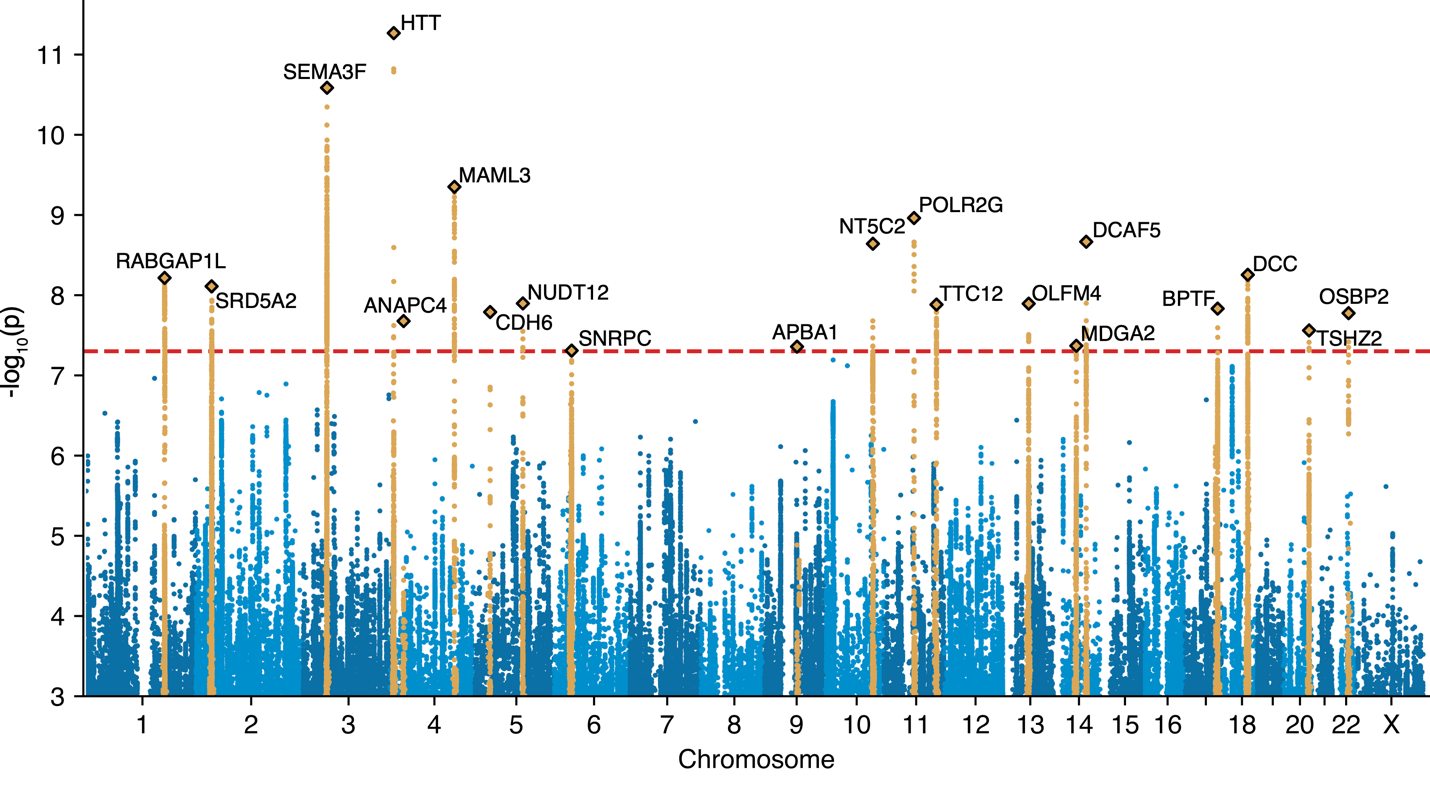

**Figure S4: Manhattan plot of the European-only, female-only meta-analysis.** Gold-highlighted variants have linkage disequilibrium r^2^ > 0.001 and are within 5 megabases of the lead variants (diamonds). The nearest gene to each lead variant is labeled. The male version of this analysis is not shown here because it would be the same as the bottom half of Figure S2: the male-only meta-analysis is restricted to European males due to low male case counts in non-European ancestries (see Methods).

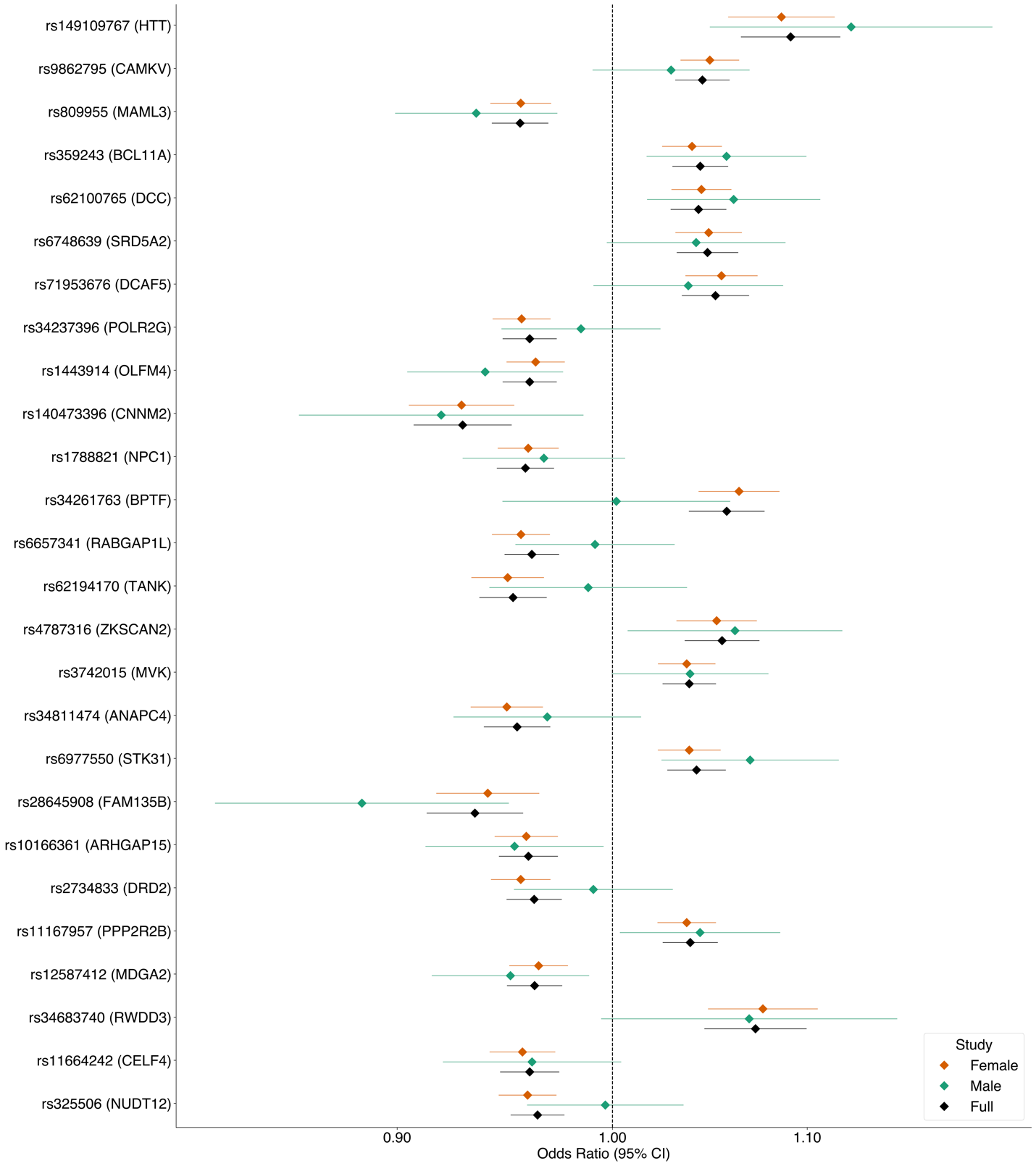

**Figure S5: Sex differences in fibromyalgia lead variant associations.** Male, female, and full-cohort odds ratios for the 26 lead variants from the primary meta-analysis.

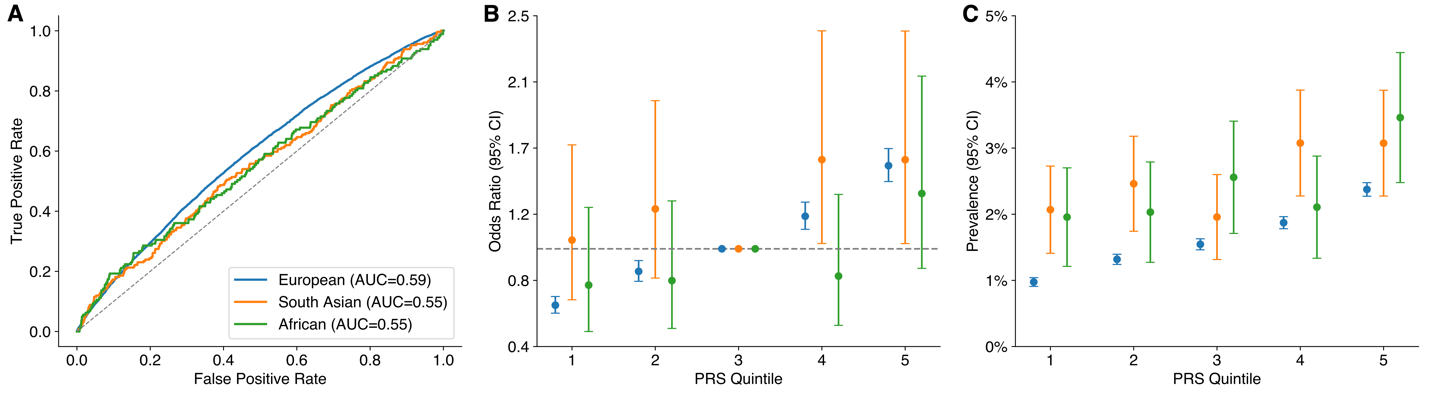

**Figure S6: Polygenic risk for fibromyalgia in the UK Biobank.** Genetic ancestry-stratified a) areas under the receiver-operating curve, b) odds ratios for each quintile of polygenic risk relative to the middle quintile, and c) prevalences of fibromyalgia within each quintile, using polygenic risk scores computed from the multi-ancestry leave-UK Biobank-out meta-analysis.

### Supplementary Tables

**Supplementary Table S1.** Case and control sample sizes in full-cohort analysis stratified by cohort, ancestry, and sex.

**Supplementary Table S2.** Lead variants and candidate causal genes from sex-specific meta-analyses.

**Supplementary Table S3.** Candidate causal genes prioritized through functional annotations and identification of coding variants in high linkage disequilibrium with lead variants.

**Supplementary Table S4.** GWAS Catalog associations for lead variants and variants in high linkage disequilibrium with lead variants.

**Supplementary Table S5.** Disease phenome-wide association study associations of lead variants queried from MVP + FinnGen + UKBB meta-analysis summary statistics.

**Supplementary Table S6.** Drug phenome-wide association study associations of lead variants queried from FinnGen summary statistics.

**Supplementary Table S7.** Tissue enrichment results from LDSC-SEG analysis.

**Supplementary Table S8.** Cell type enrichment results from LDSC-SEG analysis of main mouse cell types.

**Supplementary Table S9.** Cell type enrichment results from LDSC-SEG analysis of mouse cell lineages.

**Supplementary Table S10.** Genetic correlation results between fibromyalgia and FinnGen disease endpoints.
